# CD4^+^ T cell mitochondrial genotype in Multiple Sclerosis: a cross-sectional and longitudinal analysis

**DOI:** 10.1101/2023.03.22.23287580

**Authors:** Filipe Cortes-Figueiredo, Susanna Asseyer, Claudia Chien, Hanna G. Zimmermann, Klemens Ruprecht, Tanja Schmitz-Hübsch, Judith Bellmann-Strobl, Friedemann Paul, Vanessa A. Morais

## Abstract

Multiple Sclerosis (MS) is a chronic autoimmune demyelinating disease of the central nervous system (CNS), with a largely unknown etiology, where mitochondrial dysfunction significantly contributes to neuroaxonal loss and brain atrophy. Mirroring the CNS, peripheral immune cells from patients with MS, particularly CD4^+^ T cells, show inappropriate mitochondrial phenotypes and/or oxidative phosphorylation (OxPhos) insufficiency, with a still unknown contribution of mitochondrial DNA (mtDNA). We hypothesized that mitochondrial genotype in CD4^+^ T cells might influence MS disease activity and progression.

Thus, we performed a retrospective cross-sectional and longitudinal study on patients with a recent diagnosis of either Clinically Isolated Syndrome (CIS) or Relapsing-Remitting MS (RRMS) at two timepoints: six months (VIS1) and 36 months (VIS2) after disease onset. Our primary outcomes were the differences in mtDNA extracted from CD4^+^ T cells between: (**I**) patients with CIS/RRMS (PwMS) at VIS1 and age- and sex-matched healthy controls (HC), in the cross-sectional analysis, and (**II**) different diagnostic evolutions in PwMS from VIS1 to VIS2, in the longitudinal analysis.

We successfully performed mtDNA whole genome sequencing (WGS) (mean coverage: 2055.77 reads/base pair) in 183 samples (61 triplets). Nonetheless, mitochondrial genotype was not associated with a diagnostic of CIS/RRMS, nor with longitudinal diagnostic evolution.

## 4. Introduction

Multiple Sclerosis (MS) is a chronic neuroinflammatory and neurodegenerative disease with a largely unknown etiology, secondary to an autoimmune demyelination in the central nervous system (CNS). Histopathologically, it is characterized by gliosis, oligodendrocyte death, and neuroaxonal loss [1]. Worldwide, approximately 2.8 million patients with MS deal with significant levels of disability [2].

Several studies have shown that the autoimmune response seen in MS mostly derives from a dysfunctional autoreactive CD4^+^ T cell compartment [3–5], although other pro-inflammatory cells, such as B cells and myeloid cells, seem to be implicated as well [6]. Recently, mitochondria have also been shown to play a role in driving MS disease activity and progression. In the CNS, mitochondrial dysfunction has been found to be a critical trigger for neuroaxonal loss and brain atrophy [7,8]. In the peripheral immune compartment, CD4^+^ T cells from patients with MS show oxidative phosphorylation (OxPhos) insufficiency [9–11], which, in animal models, has been linked to an exacerbation of CNS autoimmune-mediated inflammation [12,13].

In parallel, mutations in mitochondrial DNA (mtDNA), which is responsible for encoding 22 transfer RNAs (tRNAs), two ribosomal RNAs (rRNAs), and 13 proteins of the OxPhos chain, as well as particular haplogroups, which are inherited mutational patterns that may be classified into phylogenetic clusters [14,15], have also been associated with an increased risk of MS, albeit not consistently [16–20].

Interestingly, mtDNA polymorphisms have been shown to modulate both metabolism and immunity [21,22], and mtDNA variants have significant tissue-specificity, including in T cells [23]. Nonetheless, whether the aforementioned CD4^+^ T cell OxPhos dysfunction in patients with MS is an inherent consequence of the particular mitochondrial genotype of this immune subset remains unknown.

We hypothesized that mitochondrial genotype in CD4^+^ T cells might influence MS disease activity and progression. Thus, we aimed to explore the differences in mtDNA extracted from CD4^+^ T cells between patients with a recent diagnosis of either Clinically Isolated Syndrome (CIS) or Relapsing-Remitting MS (RRMS) and healthy controls (HC). We also analyzed longitudinal mtDNA changes in patients with CIS/RRMS (PwMS).

## 5. Materials and Methods

### 5.1. Cohort description

We performed an observational retrospective evaluation of prospectively collected data within the Berlin CIS-Cohort (reference: NCT01371071) [24] at the Charité — Universitätsmedizin Berlin, Berlin, Germany. Berlin CIS-Cohort’s inclusion criteria are: (**I**) age ≥ 18 years and (**II**) diagnosis of either Clinically Isolated Syndrome (CIS) within six months from symptom onset or of Relapsing-Remitting Multiple Sclerosis (RRMS) within two years from symptom onset, according to the 2017 revisions of the McDonald criteria [25].

To address the disease activity and progression of patients with CIS/RRMS (PwMS), the following variables were assessed on each clinical visit: number of relapses and time to last relapse; expanded disability status scale (EDSS) and Multiple Sclerosis functional composite (MSFC) scores; brain MRI: T2 hyperintense lesions and T1 gadolinium-enhancing lesions; and optical coherence tomography (OCT): peripapillary retinal nerve fiber layer (RNFL) thickness and ganglion cell-inner plexiform layer (GCIPL) volume. Additionally, we assessed if patients fulfilled the criteria for no evidence of disease activity (NEDA)-3, namely, absence of new relapses, on MRI: no T1 gadolinium-enhancing lesions and no new or enlarging T2 hyperintense lesions and the absence of EDSS worsening. Details of MRI, OCT, and NEDA-3 were described earlier [26]. Finally, blood samples were collected and, subsequently, peripheral blood mononuclear cells (PBMCs) were isolated. Further details on the methodology used for collection and analysis of clinical data may be found in Section 1 of the Supplementary Information.

PwMS were included in this study if biological samples and a clinical assessment were available for two clinical visits: six months (VIS1) and 36 months (VIS2) after disease onset. Recruitment of HC without a family history of MS was finalized in May 2019; HC were matched to PwMS in a 1:1 ratio according to sex and a maximum age difference of five years.

Our primary outcomes were a mitochondrial genotype comparison between: (**I**) PwMS and HC in the cross-sectional analysis and (**II**) different diagnostic evolutions from VIS1 to VIS2 in the longitudinal analysis, namely, CIS in both clinical visits, a conversion from CIS to RRMS, and RRMS in both clinical visits.

Since mtDNA whole genome sequencing (WGS) in CD4^+^ T cells from PwMS was unreported in the literature, a pilot study with 20 triplets (20 PwMS at VIS1&VIS2 and 20 age- and sex-matched HC) was performed to determine the appropriateness of the sample size through the Dupont method [27].

#### 5.1.1. Ethical approval

The institutional review board (IRB)’s approval was obtained by the Ethics Committee of the Charité — Universitätsmedizin Berlin (application number: EA1/182/10), informed consents were given by every subject, and the study followed the standards of the Declaration of Helsinki [28].

### 5.2. Sample processing

#### 5.2.1. CD4^+^ T cell enrichment and flow cytometry analysis

PBMC sample processing was performed at the same time for each triplet (PwMS at VIS1&VIS2 and HC), to minimize differences within processing. The MojoSort™ Human CD4 T Cell Isolation Kit (#480010, BioLegend, San Diego, CA, USA) was used for CD4^+^ T cell enrichment, in accordance with the manufacturer’s instructions.

To assess whether CD4^+^ T cell enrichment was achieved, flow cytometry was performed in a BD LSRFortessa™ X-20 Cell Analyzer (BD Biosciences, Franklin Lakes, NJ, USA) on a subset of samples (before and after magnetic enrichment). The following antibodies were used: CD14-eFluor450 (#48014941, clone 61D3, eBioscience™, Thermo Fisher Scientific, Waltham, MA, USA); CD19-Alexa Fluor 647 (#302222, clone HIB19, BioLegend, San Diego, CA, USA); CD3-PerCP-Cy5.5 (#45003741, clone OKT3, eBioscience™, Thermo Fisher Scientific, Waltham, MA, USA); CD4-BV711 (#317439, clone OKT4, BioLegend, San Diego, CA, USA); and CD56-PE-Cy7 (#25056741, clone CMSSB/NCAM, eBioscience™, Thermo Fisher Scientific, Waltham, MA, USA). Regarding the gating strategy, CD4^+^ T cells were defined as CD3^+^CD4^+^CD19^−^CD56^−^ single cells (Figure S5 and Figure S6). A detailed protocol and additional details regarding the methodology used for CD4^+^ T cell enrichment and flow cytometry may be found in Section 2 of the Supplementary Information.

#### 5.2.2. DNA extraction and mtDNA sequencing, including bioinformatic processing and data analysis

Similarly, DNA extraction was performed at the same time for each triplet with the QIAamp^®^ DNA Blood Midi Kit (QIAGEN GmbH, Hilden, Germany), according to the manufacturer’s instructions. Samples were sequenced with the Applied Biosystems™ Precision ID mtDNA Whole Genome Panel (Thermo Fisher Scientific, Waltham, MA, USA) at IPATIMUP — Instituto de Patologia e Imunologia Molecular da Universidade do Porto (Porto, Portugal). Libraries were prepared using the Ion Chef™ automated protocol and samples were run on 530™ chips with the Ion Torrent™ Ion S5™ (Thermo Fisher Scientific, Waltham, MA, USA). Samples with a coverage uniformity < 85% and/or a mean coverage < 1500 reads were resequenced.

Regarding bioinformatic processing, the PrecisionCallerPipeline (PCP) pipeline [29] was used, with mutserve v2 [30,31] for variant calling, HaploGrep v2.4.0 [15] for haplogroup calling, and Haplocheck v1.3.3 [31] for a contamination check. To account for possible false positives, variants with a variant level (VL) ≥ 10% were only accepted if they were found with both the PCP pipeline and the Ion Torrent Suite™ Software (TSS), while variants below TSS’s limit of detection (10%) and solely present with PCP where filtered in accordance with: (**I**) sequencing indicators for variant reliability (normalized coverage, coverage ratio, mean value of reported nuclear insertions of mitochondrial DNA [NUMTs], and the distance to the amplicon’s edge); and (**II**) previous reports of the same variants in curated mtDNA databases. Variants only present in TSS were excluded [29]. Data analysis was performed with R version 4.1.1 [32] and Excel 2016 (Microsoft Corporation, Redmond, WA, USA).

The level of statistical significance was set at a two-sided *p*-value < 0.05 for all tests and multiple comparison testing was adjusted with false discovery rate (FDR). Whenever data was missing, it was censored from the analysis. Reporting followed the standards from STROBE [33] and its extension STREGA [34].

A detailed description of the methodology regarding DNA extraction, bioinformatic processing, and data analysis may be found in Section 3 of the Supplementary Information.

## 6. Results

### 6.1. Cohort description

Overall, 61 PwMS were included in this study cohort (Table 1). Most patients presented with RRMS at both clinical visits, mean follow-up time between VIS1 and VIS2 was 30.50 ± 1.27 (mean ± SD) months. Approximately one third of the PwMS (*N*=20) suffered a relapse between VIS1 and VIS2, while PwMS under MS immunomodulatory treatment increased from 26.23% to 47.54%. The average age difference between PwMS at VIS1 and HC was 2.06 ± 1.31 (mean ± SD) years.

**Table 1.** Cohort characteristics.

| Variables | HC | PwMS |  | N<br>(existing data at<br>both VIS1 and<br>VIS2) |
| --- | --- | --- | --- | --- |
|  |  | V1 | V2 |  |
| Age at cohort entry (years) | 32.78 [19–55] | 32.52 [21–56] | - | 61 |
| Females |  | N=40 (66%) |  |  |
| Date of PBMC collection | 2015/06 –<br>2019/05 | 2011/05 –<br>2015/05 | 2014/01 –<br>2017/11 | 61 |
| Diagnostic evolution | - | CIS - CIS, N=14 (23%)<br>CIS - RRMS, N=8 (13%)<br>RRMS - RRMS, N=39 (64%) |  | 61 |
| Immunomodulatory<br>treatment | - | N=16 (26%) | N=29 (48%) | 61 |
| EDSS | - | 1.50 [0.00–3.50],<br>N=61 | 1.50 [0.00–4.00],<br>N=60 | 60 |
| N. relapses | - | 0.00 [0.00–1.00],<br>N=61 | 0.00 [0.00–5.00],<br>N=61 | 61 |
| Time to last relapse(days) | - | 164.85 [29.00–<br>234.00], N=60 | 898.10 [10.00–<br>1187.00], N=61 | 60 |
| NEDA-3 | - | N=9 (15%) |  | 59 |
| MSFC | - | -0.01 [-0.96–<br>0.76], N=59 | 0.17 [-1.16–1.01],<br>N=58 | 57 |
| N. T2 les. | - | 6.00 [0.00–<br>98.00], N=61 | 8.50 [0.00–<br>91.00], N=60 | 60 |
| V. T2 les. (mL) | - | 2.04 [0.00–<br>17.97], N=61 | 2.52 [0.00–<br>19.16], N=60 | 60 |
| N. T1 gd les. | - | 0.00 [0.00–3.00],<br>N=49 | 0.00 [0.00–4.00],<br>N=22 | 15 |
| V. T1 gd les. (mL) | - | 0.01 [0.00–0.28],<br>N=49 | 0.03 [0.00–0.26],<br>N=22 | 15 |
| GCIPL (mm<sup>3</sup>) | - | 1.98 [1.46–2.32],<br>N=51 | 1.96 [1.44–2.30],<br>N=58 | 50 |
| RNFL (μm) | - | 97.24 [68.00–<br>126.00], N=54 | 97.56 [64.50–<br>132.50], N=58 | 52 |

Numbers show the mean for each value followed by the minimum and maximum values between brackets, except for EDSS, number of relapses, number of T2 lesions, and number of T1 lesions, where the median value is shown instead of the mean. Abbreviations: EDSS — expanded disability status scale; GCIPL — ganglion cell-inner plexiform layer; gd — gadolinium; HC — healthy controls; les. — lesions; MSFC — Multiple Sclerosis functional composite; N. — number; NEDA — no evidence of disease activity; PBMC — peripheral blood mononuclear cell; PwMS — patients with Clinically Isolated Syndrome/Relapsing-Remitting Multiple Sclerosis; RNFL — retinal nerve fiber layer; V. — volume; V1 — patients with Clinically Isolated Syndrome/Relapsing-Remitting Multiple Sclerosis at visit 1; V2 — patients with Clinically Isolated Syndrome/Relapsing-Remitting Multiple Sclerosis at visit 2; VIS1 — visit 1; VIS2 — visit 2.

As mentioned previously, a pilot study with 20 triplets (20 PwMS at VIS1&VIS2 and 20 age- and sex-matched HC) was performed to determine the appropriateness of the sample size through the Dupont method [27]. Following WGS data analysis of this subset, a 35% discordance in prevalence was detected for deleterious variants in Complex I — 55% for PwMS at VIS1 (discordant prevalence: 45%) *vs*. 20% for HC (discordant prevalence: 10%). Hence, the sample size was adequate according to this endpoint.

### 6.2. CD4^+^ T cell enrichment, DNA extraction, and WGS quality

Following the magnetic enrichment in CD4^+^ T cells, we obtained a wide range in the number of cells and cell mortality, for all subject types (Table 2 and Table S5). In comparison with HC, PwMS at VIS1 showed a lower number of cells (mean difference of 0.65 million, 95% confidence interval [CI] 0.16– 1.14; paired t-test adjusted with FDR). In comparison with VIS2, cells from PwMS at VIS1 had lower mortality (mean difference of 2.77%, 95% CI 0.56–4.97%; paired t-test adjusted with FDR). Nonetheless, a significant increase in CD4^+^ T cells (CD3^+^ CD4^+^ CD56^−^ CD19^−^) was achieved (Figure 1A, and Table S3: mean fold change in paired samples of 2.59, 95% CI 2.31–2.87; paired t-test, *N*=49), regardless of subject type (Table S4).

**Table 2.** Magnetic enrichment and DNA extraction: Summary.

| Subject type | Number of cells after enrichment (millions) | Mortality | DNA (Synergy HTX) (ng/μL) | 260/280 ratio | DNA (Qubit®) (ng/μL) |
| --- | --- | --- | --- | --- | --- |
| HC | 3.03<br>[0.37–8.92] | 15.15%<br>[4.34–70.86%] | 20.04<br>[1.45–65.24] | 1.87<br>[1.19–2.62] | 13.78<br>[1.91–56.00] |
| V1 | 2.38<br>[0.57–6.40] | 14.97%<br>[5.57–33.41%] | 15.59<br>[2.93–45.33] | 1.89<br>[1.19–3.37] | 10.43<br>[2.54–37.00] |
| V2 | 2.48<br>[0.10–8.10] | 17.73%<br>[7.43–56.25%] | 15.94<br>[3.48–53.46] | 1.93<br>[1.15–5.20] | 10.54<br>[0.70–27.20] |
| **All** | **2.63**<br>**[0.10–8.92]** | **15.95%**<br>**[4.34–70.86%]** | **17.19**<br>**[1.45–65.24]** | **1.90**<br>**[1.15–5.20]** | **11.58**<br>**[0.70–56.00]** |

Numbers show the mean for each value followed by the minimum and maximum values between brackets. Abbreviations: HC — healthy controls; V1 — patients with Clinically Isolated Syndrome/Relapsing-Remitting Multiple Sclerosis at visit 1; V2 — patients with Clinically Isolated Syndrome/Relapsing-Remitting Multiple Sclerosis at visit 2.

**Figure 1.**
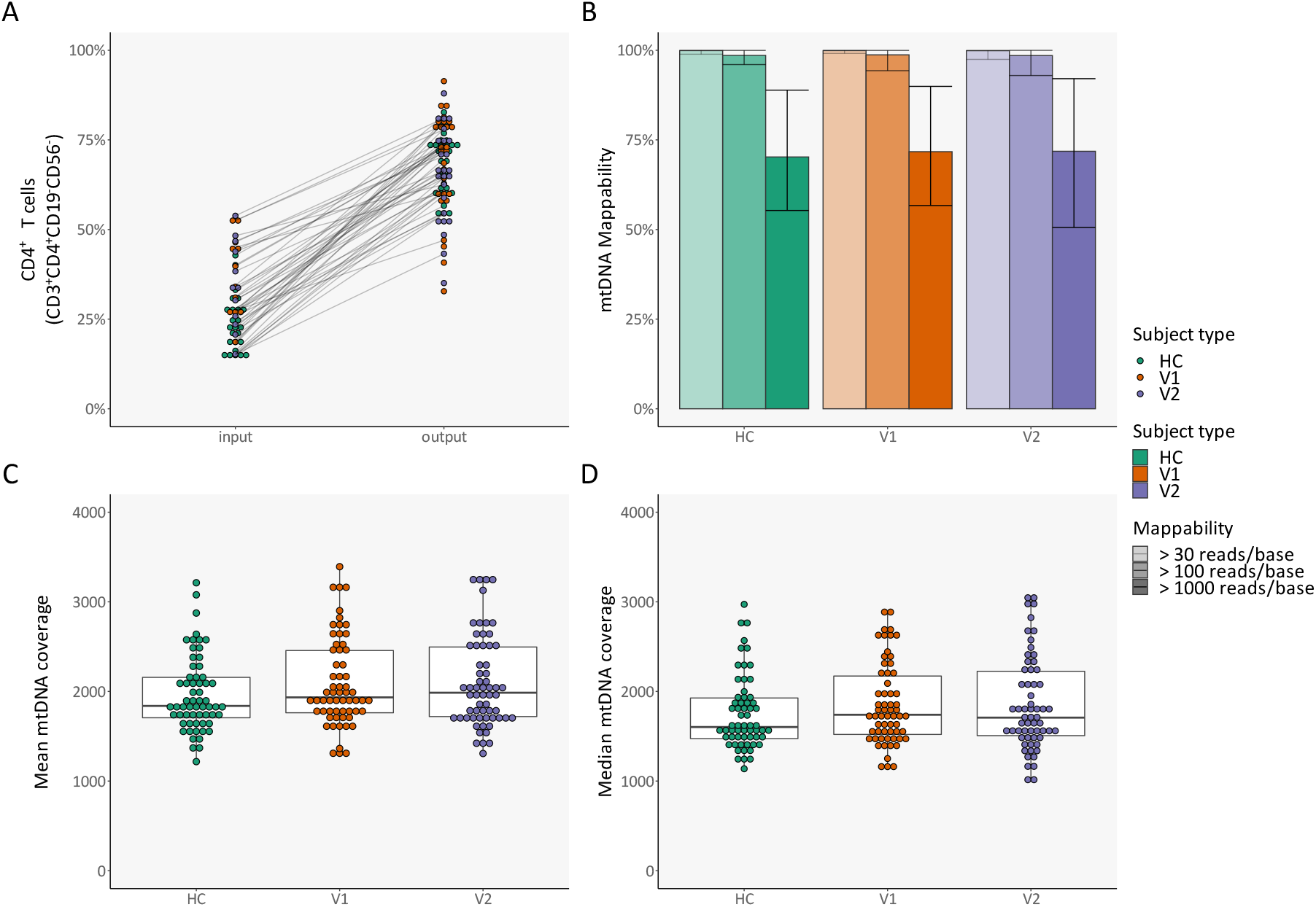
CD4^+^ T cell enrichment and mitochondrial DNA whole genome sequencing. (**A**) Percentage of CD4^+^ T cells (CD3^+^ CD4^+^ CD56^−^ CD19^−^) before and after magnetic enrichment, as assessed with flow cytometry — lines connect the same sample; (**B**) Mean mappability, per subject type and per definition — error bars denote minimum and maximum values; (**C**) Mean mtDNA coverage, per subject type; (**D**) Median mtDNA coverage, per subject type. Abbreviations: HC — healthy control; mtDNA — mitochondrial DNA; V1 — patient with Clinically Isolated Syndrome/Relapsing-Remitting Multiple Sclerosis at visit 1; V2 — patient with Clinically Isolated Syndrome/Relapsing-Remitting Multiple Sclerosis at visit 2.

Subsequently, after the extraction of DNA, variable amounts of DNA were achieved (Table 2 and Table S5). In comparison with PwMS at VIS1, HC had higher yields of extracted DNA (mean difference of 4.45 ng/μL for the measurement with Synergy HTX, 95% CI 1.06–7.84, and mean difference of 3.35 ng/μL for the measurement with Qubit®, 95% CI 0.80–5.90; paired t-tests adjusted with FDR). However, while, as expected, the two DNA measurements showed a correlation between each other (Figure S9A: adjusted R^2^ of 0.88 and a *p*-value < 2.2 × 10^−16^, linear regression model) and the number of cells used for DNA extraction correlated with DNA concentration (Figure S9B: adjusted R^2^ of 0.77 and a *p*-value < 2.2 × 10^−16^, linear regression model), DNA concentration had no effect on mtDNA coverage (Figure S9C–D), with most variability appearing to arise from each sequencing run (Figure S9E–F). Correspondingly, no differences in mtDNA coverage and mappability were found between different subject types (Figure 1B–D and Figure S9E–F).

Taking into account all samples analyzed through WGS (Table S7), no contamination was detected and all haplogroups corresponded to European lineages. PwMS had the same haplogroup with two minor exceptions (Triplets #5 and #48) at both visits, albeit without changing their simplified haplogroup (Table S7).

### 6.3. Cross-sectional comparison

The total number of variants was similar between different subject types, i.e., between HC and PwMS at VIS1 and between VIS1 and VIS2 for PwMS (Figure 2 and Table S7: paired t-tests adjusted with FDR). Variant distribution was also bimodal in all subject types (Figure 2A), likely arising from each sample’s haplogroup (Figure 2B–C), since the mean number of variants in HC and PwMS at VIS1 was significantly different in haplogroups with at least three samples (*p*-value 3.34 × 10^−16^; Kruskal-Wallis test). Haplogroups H and HV had significantly fewer variants (Table S8), which is consistent with the revised Cambridge Reference Sequence (rCRS)’s own H haplogroup [35]. Nonetheless, haplogroup distribution was independent from subject type (HC and PwMS at VIS1; Fisher’s exact test with Monte Carlo simulations).

**Figure 2.**
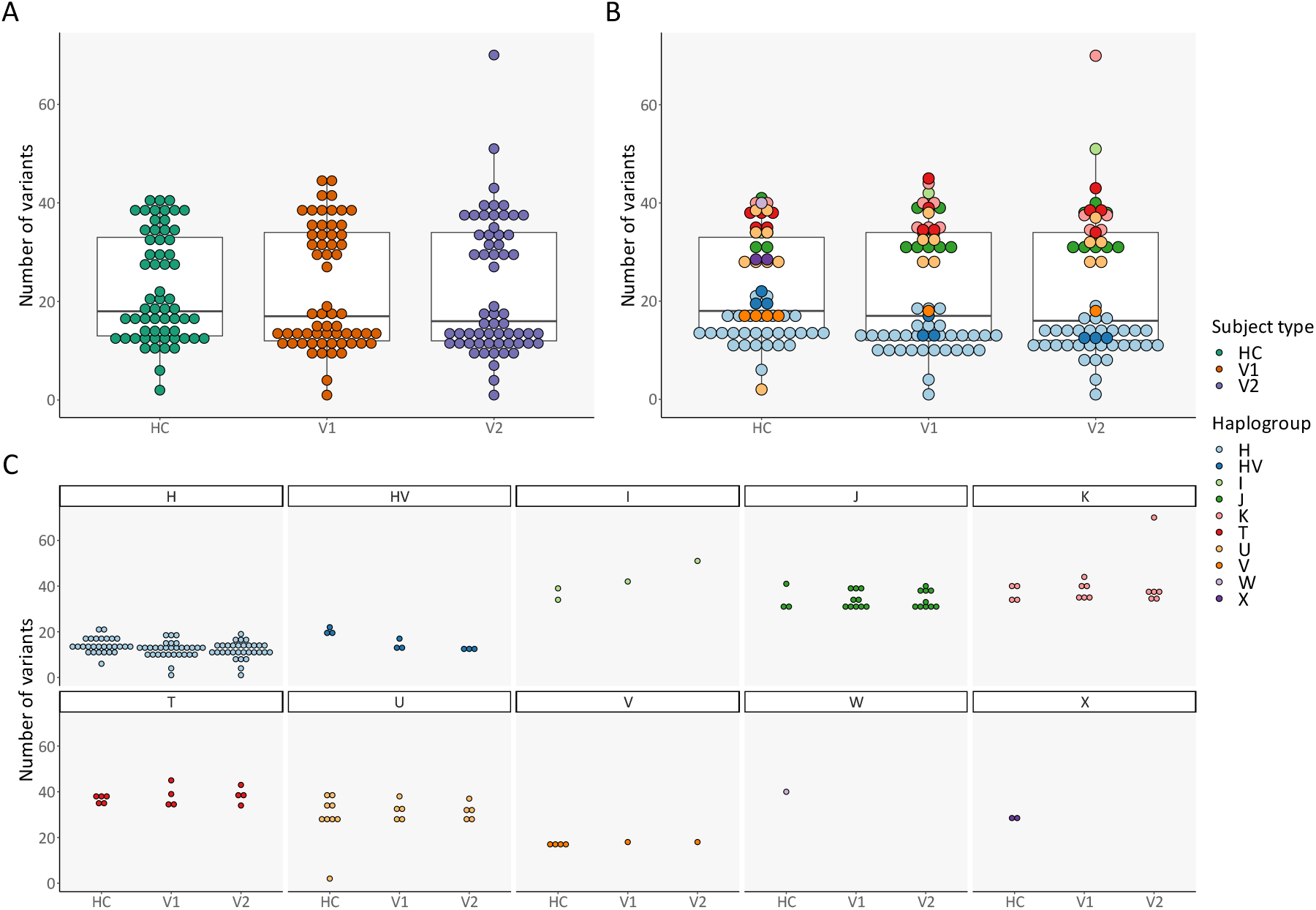
Total number of variants: Cross-sectional comparison (subject types and haplogroups) (**A**) Number of variants, per subject type; (**B**) Number of variants, per subject type and per haplogroup; (**C**) Expansion of **Figure 2B** for better visualization. Abbreviations: HC — healthy control; V1 — patient with Clinically Isolated Syndrome/Relapsing-Remitting Multiple Sclerosis at visit 1; V2 — patient with Clinically Isolated Syndrome/Relapsing-Remitting Multiple Sclerosis at visit 2.

When comparing HC and PwMS at VIS1, we observed that the total number of variants did not vary according to age (Figure S10A: linear regression model). The difference in the number of mutations between HC and PwMS at VIS1 was also not influenced by sex (Figure S10B–C: two sample t-test [Figure S10B], χ^2^ test of independence [Figure S10C], and two-proportions z-test [Figure S10C]).

When looking at the differences between PwMS at VIS1 and HC in various mtDNA regions, the highest discordancy in prevalence was in *MT-ND3* for PwMS at VIS1 and in *MT-RNR2* for HC (Figure S11A), while the highest discordant adjusted mutational rate, which is the sum of all VLs in a specific region divided by its region size, was in *MT-TA* for PwMS at VIS1 and in *MT-TT* for HC (Figure S11B). However, there were no significant differences for discordant prevalence (Figure S11A: McNemar’s tests adjusted with FDR); nor for other discordant mutational rates (Figure S11B: paired t-tests adjusted with FDR).

Regarding macro regions in mtDNA, the most commonly discordant affected region for PwMS at VIS1 was *Other*, which refers to positions in rCRS with an overlap between the two strands or left unannotated, and, for HC, transfer RNA (tRNA) (Figure S11C), while the region with the highest discordant adjusted mutational rate was Complex III for PwMS at VIS1 and *Other* for HC (Figure S11D). Nonetheless, there were no significant differences for discordant prevalence (Figure S11C: McNemar’s tests adjusted with FDR); nor for discordant mutational rates (Figure S11D: paired t-tests adjusted with FDR).

We further took all variants into account (Table S9), to see if individual variants differed between HC and PwMS at VIS1. However, no significant differences were found (Table S9: McNemar’s tests adjusted with FDR). Interestingly, a single PwMS had a pathogenic variant, namely, m.11778G>A in the *MT-ND4* gene (Table S9), which is associated with Leber’s hereditary optic neuropathy (LHON) and neuropathy, ataxia, and retinitis pigmentosa (NARP), albeit at low VLs: 5.8% and 7.3% for VIS1 and VIS2, respectively; well below the VLs usually required for pathogenesis [36]. The patient in question initially presented with pyramidal and sensory complaints, maintaining the latter throughout their disease course, without any visual changes.

Subsequently, we took into account variants predicted *in silico* to be likely deleterious for proteins, defined by a mean value > 0.5 from two independent scores: (**I**) MutPred [37,38]; and (**II**) APOGEE [39] (Figure S12 and Figure 3). Nevertheless, the number of deleterious variants (Figure S12A) and the cumulative deleterious burden (Figure 3A), which corresponds to the total sum of a variant’s VL multiplied by its deleterious score per sample, did not differ significantly between the different subject types (paired t-tests adjusted with FDR).

**Figure 3.**
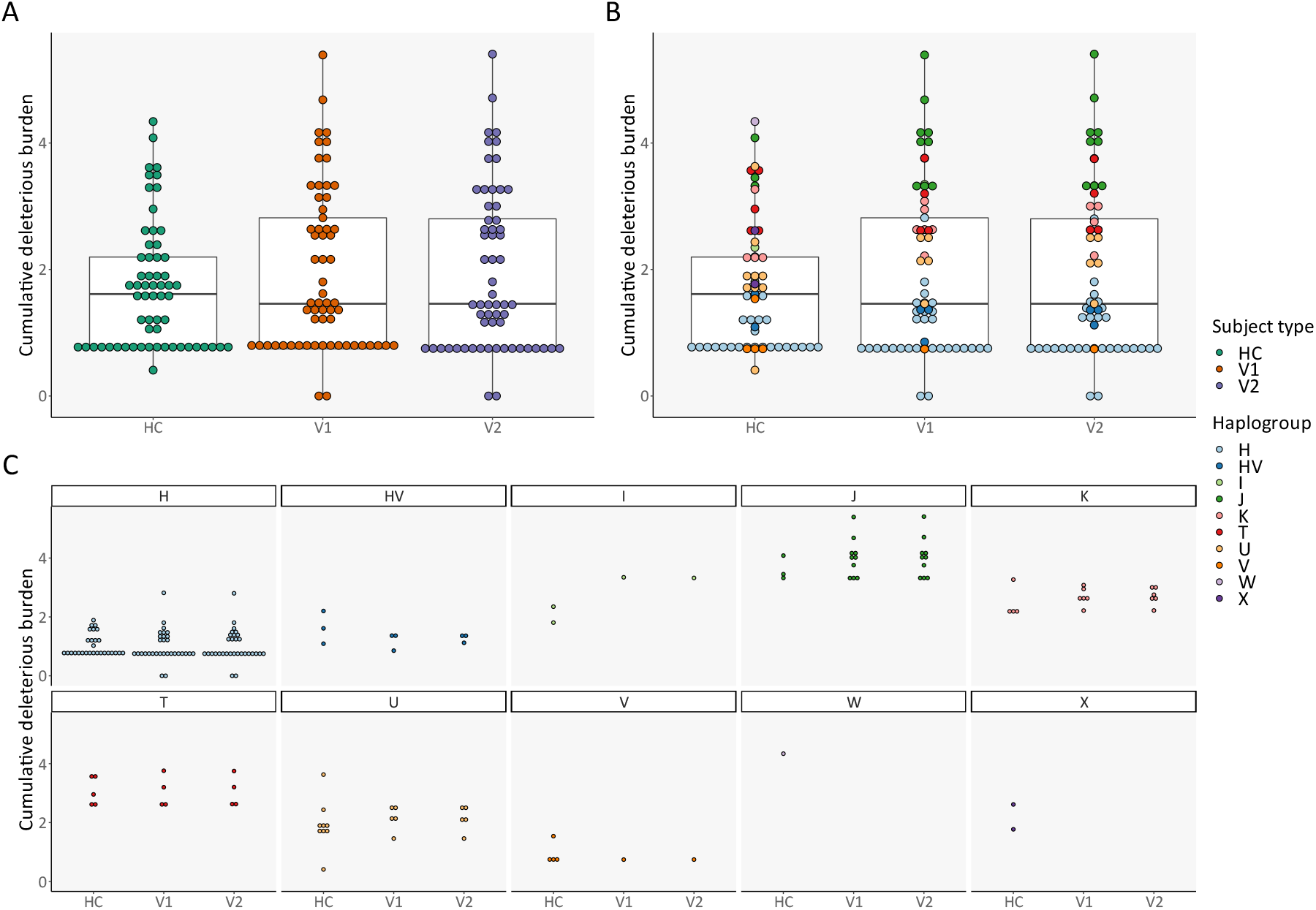
Cumulative deleterious burden: Cross-sectional comparison (subject types and haplogroups) (**A**) Cumulative deleterious burden, per subject type; (**B**) Cumulative deleterious burden, per subject type and haplogroup; (**C**) Expansion of **Figure 3B** for better visualization. Abbreviations: HC — healthy control; V1 — patient with Clinically Isolated Syndrome/Relapsing-Remitting Multiple Sclerosis at visit 1; V2 — patient with Clinically Isolated Syndrome/Relapsing-Remitting Multiple Sclerosis at visit 2.

We observed a similar haplogroup-specific effect for both the number of deleterious mutations and cumulative deleterious burden (Figure S12B–C and Figure 3B–C: *p*-values 4.61 × 10^−13^ and 1.38 × 10^− 15^, respectively; Kruskal-Wallis tests). Haplogroups J and T had significantly more deleterious variants (Table S10) and cumulative deleterious burden (Table S11).

When comparing HC and PwMS at VIS1, the number of deleterious variants did not vary according to age (Figure S13A: Kendall rank correlation test). The difference in the number of deleterious mutations between HC and PwMS at VIS1 was also not influenced by sex (Figure S13B–C: two sample t-test [Figure S13B], χ^2^ test of independence [Figure S13C], and two-proportions z-test [Figure S13C]). The same was true for cumulative deleterious burden (linear regression model for age [Figure S13D] and two sample t-test for sex [Figure S13E]).

Regarding differences between PwMS at VIS1 and HC in various mtDNA regions, the highest discordancy in prevalence was in *MT-ND5* for PwMS at VIS1 and in *MT-ATP6* for HC (Figure 4A). In parallel, the highest discordant adjusted cumulative deleterious rate, which is the cumulative deleterious rate for a specific region relative to its region size, was in *MT-ND3* for PwMS at VIS1 and in *MT-ATP6* for HC (Figure 4B). Regardless, no significant differences were found for discordant prevalence (Figure 4A: McNemar’s tests adjusted with FDR); nor for discordant cumulative deleterious rates (Figure 4B: paired t-tests adjusted with FDR).

**Figure 4.**
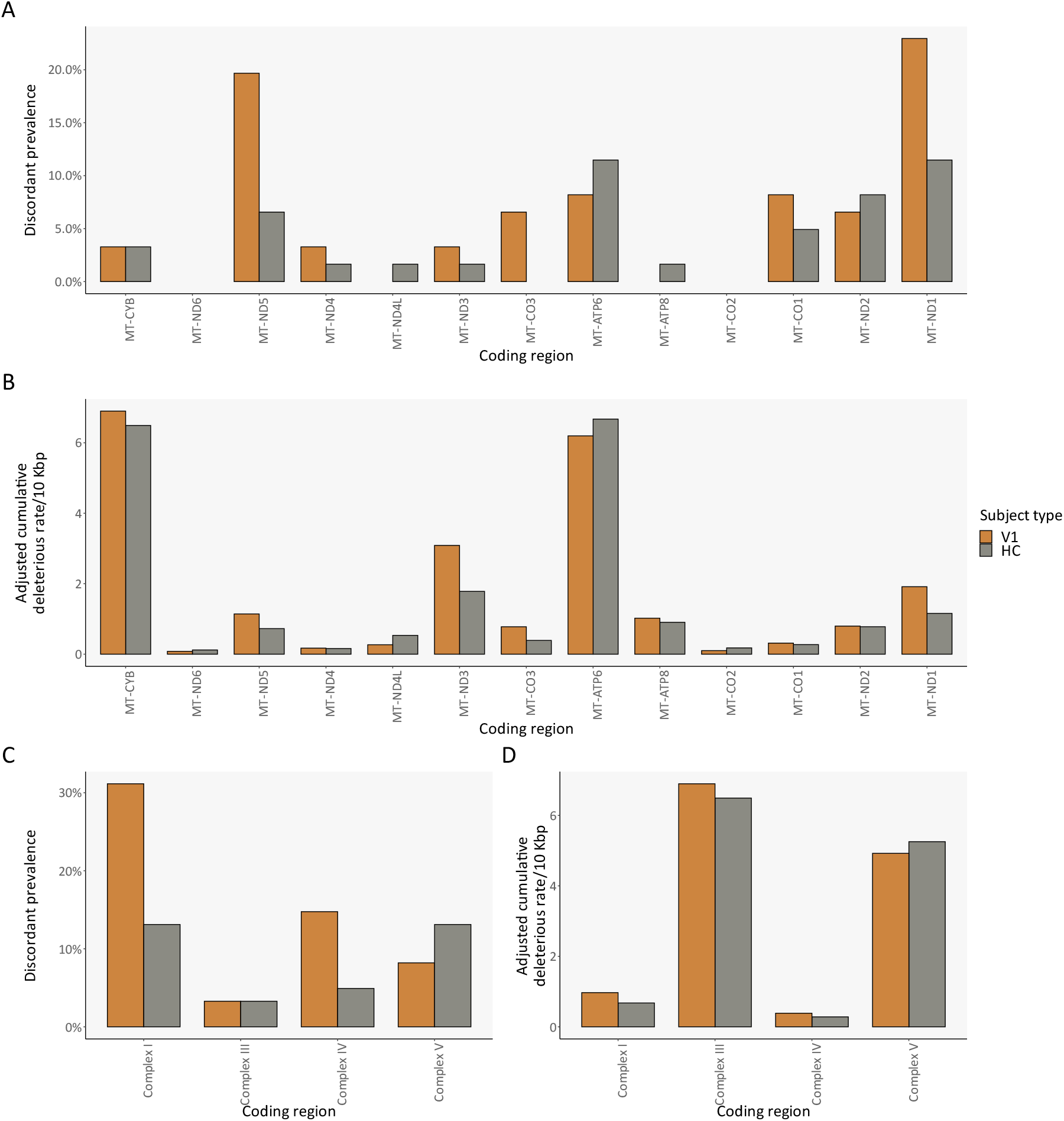
Deleterious variants: Cross-sectional comparison (regions) (**A**,**C**) Discordant prevalence of deleterious variants for each mtDNA coding region/locus and for each macro mtDNA coding region, per subject type, respectively; (**B**,**D**) Relative cumulative deleterious burden for each mtDNA coding region/locus and for each macro mtDNA coding region, per subject type, respectively. Abbreviations: bp — base pair; HC — healthy control; V1 — patient with Clinically Isolated Syndrome/Relapsing-Remitting Multiple Sclerosis at visit 1.

Regarding macro regions in mtDNA, the most commonly discordant affected region was Complex I for PwMS at VIS1 and Complex V for HC (Figure 4C), while the region with the highest adjusted cumulative deleterious rate was Complex III for PwMS at VIS1 and Complex V for HC (Figure 4D). However, there were no significant differences for discordant prevalence (Figure 4C: McNemar’s tests adjusted with FDR); nor for cumulative deleterious rate (Figure 4D: paired t-tests adjusted with FDR).

Finally, we looked into tRNA mutations predicted to be pathogenic according to the MitoTIP score [40]: no variants were found (Figure S14A). The cumulative MitoTIP score, which corresponds to the total sum of a variant’s VL multiplied by its MitoTIP score per sample, was similar in all subject types (Figure S14B–C: paired t-tests adjusted with FDR).

Nonetheless, as observed previously, haplogroup had a significant influence in the cumulative MitoTIP score (Figure S14B–C: *p*-value 3.59 × 10^−13^; Kruskal-Wallis test), with haplogroups K and U having higher cumulative MitoTIP scores, whereas haplogroups H and HV had lower scores (Table S12). No influence was found regarding age (Figure S14D: linear regression model); or sex (Figure S14E: two sample t-test). The region with the highest discordant adjusted cumulative MitoTIP rate, which is the cumulative MitoTIP rate for a specific region relative to its region size, was *MT-TA* for PwMS at VIS1 and *MT-TT* for HC (Figure S14F). However, no significant differences were found (paired t-tests adjusted with FDR).

### 6.4. Longitudinal comparison

As mentioned previously, there were no significant differences between PwMS at VIS1 and at VIS2 (paired t-tests adjusted with FDR) concerning total number of variants (Figure 2A), number of deleterious variants (Figure S12A), cumulative deleterious burden (Figure 3A), and cumulative MitoTIP score (Figure S14B).

The mean proportion of mutations present in a single visit — novel variants — were 4.01% (range: 0.00–53.33%) for VIS1 and 3.38% (range: 0.00–37.14%) for VIS2 (Figure 5A), with mean VLs of 8.39% (range: 2.50–100.00%) for VIS1 and 4.66% (range: 2.50–19.60%) for VIS2 (Figure 5B); no significant differences were found between different visits (paired t-tests).

**Figure 5.**
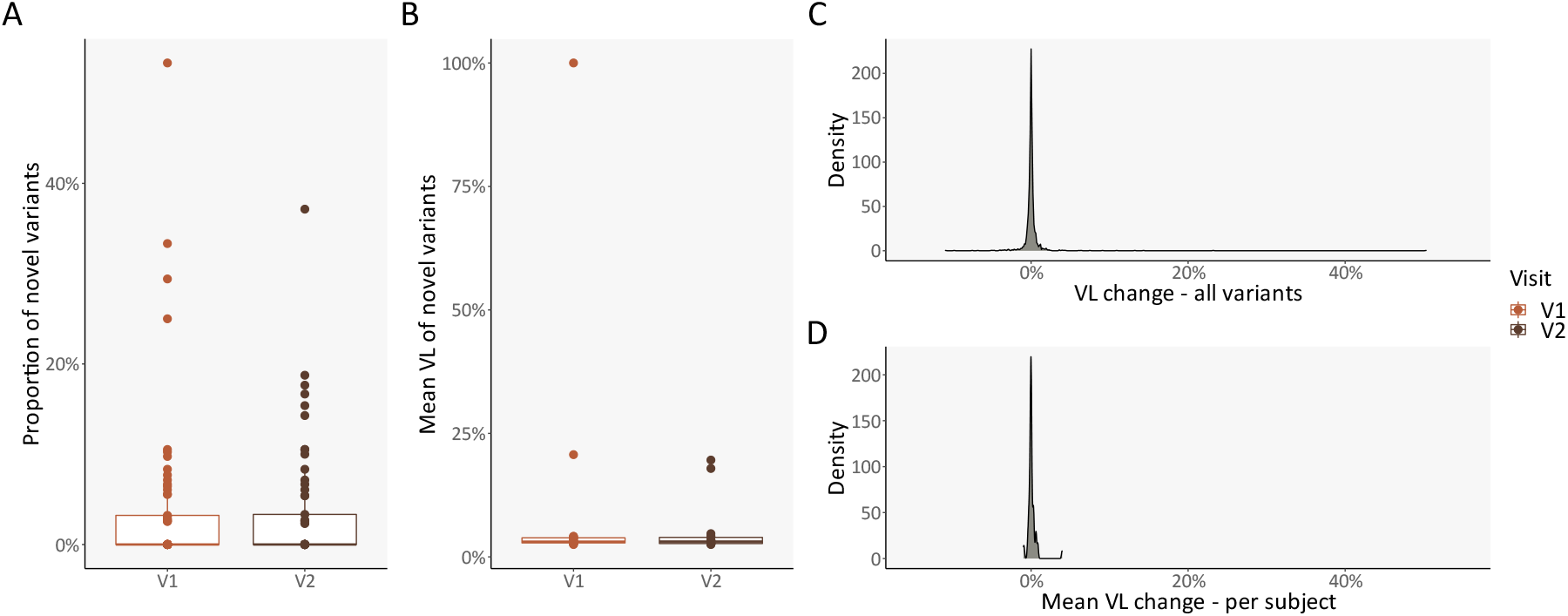
Longitudinal changes in PwMS: Intrapersonal changes. (**A**) Proportion of novel variants, per subject and clinical visit; (**B**) Mean variant level of novel variants, per subject and clinical visit; (**C–D**) Density plots of variant level change in old variants, for all mutations and per subject, respectively. Abbreviations: PwMS — patients with Clinically Isolated Syndrome/Relapsing-Remitting Multiple Sclerosis; V1 — patient with Clinically Isolated Syndrome/Relapsing-Remitting Multiple Sclerosis at visit 1; V2 — patient with Clinically Isolated Syndrome/Relapsing-Remitting Multiple Sclerosis at visit 2; VL — variant level.

Subsequently, we considered the difference in VLs for the previously found variants between the second and the first visits — old variants. The mean VL change was 0.06% in all variants (Figure 5C) and 0.09% per subject (Figure 5D) (range: -10.90–50.30%). Both VL changes were statistically similar to zero (one-sample t-test).

When we compared novel and old variants, novel variants had a significantly lower VL than old variants (Figure S15A: adjusted *p*-value 1.80 × 10^−150^; two-sample t-test adjusted with FDR) and a lower coverage ratio (Figure S15C: adjusted *p*-value 1.09 × 10^−6^; two-sample t-test adjusted with FDR). There were no significant differences for other quality control variables [29] (Figure S15B and Figure S15D–E; two-sample t-tests adjusted with FDR), nor for protein deleterious score and MitoTIP score (Figure S15F–G; two-sample t-tests adjusted with FDR).

Since each PwMS had a very similar mutational pattern between the two visits, we then moved on from an intrapersonal comparison to an interpersonal comparison, with a focus on exploring the origin of these novel variants and the VL change in old variants in the full PwMS cohort. For that purpose, we considered a PwMS as a single data point, where we performed the mean of the results from both sequencing runs.

Afterwards, we considered the various possible causes or cofactors behind differences in the proportion of novel variants, VLs of novel variants or VL change in old variants. Haplogroup (Figures S16–18A; Kruskal-Wallis test), age (Figures S16–18B; linear regression model), and sex (Figures S16– 18C; two sample t-test) did not influence the aforementioned WGS variables. Interestingly, different regions had a significant effect on the proportion of novel variants, excluding patients without novel variants and regions with mutations from less than three paired samples (Figure S16D: *p*-value 5.90 × 10^−6^; Kruskal-Wallis test), where *MT-TI* had the highest prevalence and D-loop the lowest (Table S13). This effect was absent for VLs of novel variants or VL change in old variants (Figures S17–18D; Kruskal-Wallis test).

Regarding the clinical variables in the cohort, change in MS medication status, i.e., Yes *vs*. No (Table 1 and Table S14), was independent from diagnostic evolution (Figure S19: Fisher’s exact test with Monte Carlo simulations).

To identify associations of clinical disability with diagnostic evolution, we performed batch Kruskal-Wallis tests with diagnostic evolution as a grouping variable for each available clinical variable (Figure S20 and Figure S21). Two different approaches were tested: (**I**) the mean of each clinical variable between VIS1 and VIS2 (Figure S20); and (**II**) the change of each clinical variable from VIS1 to VIS2, by subtracting VIS2 and VIS1 (Figure S21). After multiple comparison correction with FDR, only the mean values of the number and volume of T2 hyperintense lesions were associated with diagnostic change (Table S15).

Similarly, we performed batch Kruskal-Wallis tests with diagnostic evolution as a grouping variable for each WGS variable explained previously (Figure S22); no significant differences were found (Table S16). Additionally, haplogroup was independent from diagnostic evolution (Figure S23: Fisher’s exact test with Monte Carlo simulations). Similarly, as a post hoc analysis, dividing patients according to NEDA-3 status (Table 1 and Figure S24) or cumulative deleterious burden (Figures S25–26) yielded null results.

## 7. Discussion

The present study investigated whether mitochondrial genotype in CD4^+^ T cells influenced MS disease activity and progression. Accordingly, we performed WGS from CD4^+^ T cells in a matched cohort of PwMS. Overall, however, there were no significant differences regarding number of variants, number of deleterious variants, cumulative protein deleterious burden, or cumulative MitoTIP score between PwMS and HC, nor after a mean of 30.50 months of follow-up between VIS1 and VIS2 for PwMS.

Importantly, the CIS/RRMS cohort herein described is representative of the overall population of PwMS in Germany, regarding mean age at onset, female predominance, and distribution of the diagnoses, with a vast majority of RRMS [41].

According to the flow cytometry analysis performed in a subset of magnetically enriched samples, CD4^+^ T cell enrichment was successful with the MojoSort™ Human CD4 T Cell Isolation Kit, which was expected [42]. The Precision ID mtDNA Whole Genome Panel, in combination with the PCP pipeline [29], proved particularly valuable, as it was able to obtain mtDNA WGS from all samples without signs of contamination, despite a wide range of the number of cells after processing the samples, and, consequently, of DNA yield, which is consistent with previous reports [43].

Curiously, amongst the 21 most prevalent mutations in PwMS, five had already been associated with MS, ranging from case reports to large epidemiological studies [16,18,44–46]. Furthermore, remarkably, the high number of deleterious variants in complex I and IV matches the pattern of decreased activity in these complexes in CD4^+^ T cells of PwMS in previous studies [9,47]. Moreover, the higher number of deleterious variants and cumulative deleterious burden from haplogroups J and T is consistent with previous reports of increased MS risk for these haplogroups [16,17].

Nevertheless, in our cohort, the mitochondrial genotype did not differ significantly between PwMS and HC, nor between VIS1 and VIS2 within PwMS, which is consistent with previous mtDNA WGS studies in PwMS [19,20].

In contrast to the 35% discordance in prevalence for deleterious variants in Complex I registered in the pilot study, the final difference was 18%, due to an increase in prevalence of 8% for HC, while in PwMS it decreased by 9%. Thus, our sample size was incompatible with the previously-set endpoint. A further limitation might have also been the mtDNA WGS of the very heterogenous CD4^+^ T cell compartment, as well as its magnetic enrichment, as opposed to other methods aimed at achieving higher cell purity. Nonetheless, previous studies focused on CD4^+^ T cell OxPhos dysfunction in patients with MS employing magnetic enrichment and fewer subjects than the present cohort were still successful in defining a clear mitochondrial phenotype [9,47].

Despite these limitations, this study constitutes a thorough cross-sectional and longitudinal analysis of mtDNA WGS in PwMS, in addition to surveying mitochondrial genotype specifically in CD4^+^ T cells. Moreover, we were able to assess the longitudinal dynamics of mtDNA, which have only very recently started to be unveiled [48,49].

Overall, CD4^+^ T cell mitochondrial genotype was not associated with a diagnostic of CIS/RRMS, nor with longitudinal diagnostic evolution. We further postulate that mitochondrial dysfunction in CD4^+^ T cells is unlikely to derive from mitochondrial genotype.

## Data Availability

All data produced in the present study are available upon reasonable request to the authors.

## 9. Acknowledgements

We would like to thank Ana Mafalda Rocha, Bettina Vogelreuter, Bibiane Seeger-Schwinge, Filipa S. Carvalho, Jana Hermann, René Gieß, and Rosalie Schmidt for their support in the conduction of this study. Additionally, we would like to thank Susan Pikol and Cynthia Kraut for their MRI lesion segmentations and collection of MRI data related to this study. We would also like to thank the Flow Cytometry Facility of the Instituto de Medicina Molecular João Lobo Antunes for their technical support, particularly Diana Macedo. Finally, we would like to express our most sincere gratitude to all the subjects who have participated in this research project.

## 10. Author contributions

F.C.F. was responsible for: Conceptualization of the research study, execution of the experimental tasks, data analysis, and manuscript draft preparation. S.A. was responsible for: Clinical data curation and collection, subject recruitment, critical review of the study’s conceptualization, and manuscript revision. C.C. was responsible for: MRI data collection and management, MRI data analysis, and manuscript revision. H.G.Z. was responsible for: OCT data analysis and management, and manuscript revision. K.R. was responsible for: Clinical data collection, subject recruitment, and manuscript revision. T.S.-H. was responsible for: Cohort management, clinical data collection, subject recruitment, project administration, and manuscript revision. J.B.-S. was responsible for: Cohort management, clinical data collection, subject recruitment, project administration, critical review of the study’s conceptualization, and manuscript revision. F.P. and V.A.M. were responsible for: Conceptualization of the research study, funding acquisition, provision of resources, supervision of data interpretation, and manuscript revision (co-senior authors). All authors read and approved the final manuscript.

## 11. Data availability statement

Participant consent did not include sharing of individual study data. Data may be made available for the purpose of replication of results upon reasonable request from the corresponding authors.

## 12.3. List of abbreviations

bp(s): base pair(s)
CI: confidence interval
CIS: Clinically Isolated Syndrome
EDSS: expanded disability status scale
FDR: false discovery rate
GCIPL: ganglion cell-inner plexiform layer
gd: gadolinium
HC: healthy control(s)
IRB: institutional review board
les: lesions
LHON: Leber’s hereditary optic neuropathy
MS: Multiple Sclerosis
MSFC: Multiple Sclerosis functional composite
mtDNA: mitochondrial DNA
N: number
NARP: neuropathy, ataxia, and retinitis pigmentosa
NEDA: no evidence of disease activity
NUMT(s): nuclear insertion(s) of mitochondrial DNA
OCT: optical coherence tomography
OxPhos: oxidative phosphorylation
PBMC: peripheral blood mononuclear cell
PCP: PrecisionCallerPipeline
PwMS: patient(s) with Clinically Isolated Syndrome/Relapsing-Remitting Multiple Sclerosis
rCRS: revised Cambridge Reference Sequence
RRMS: Relapsing-Remitting Multiple Sclerosis
rRNA: ribosomal RNA
tRNA: transfer RNA
TSS: Ion Torrent Suite™ Software
V: volume
V1: patient(s) with Clinically Isolated Syndrome/Relapsing-Remitting Multiple Sclerosis at visit 1, six months after disease onset
V2: patient(s) with Clinically Isolated Syndrome/Relapsing-Remitting Multiple Sclerosis at visit 2, 36 months after disease onset
VIS1: visit 1
VIS2: visit 2
VL(s): variant level(s)
WGS: whole genome sequencing

