## Supplemental Information for "CD4^+^ T cell mitochondrial genotype in Multiple Sclerosis: a cross-sectional and longitudinal analysis"

### Supplementary Information

### Section 1 — Clinical data collection and analysis

#### 1.1. Clinical data collection and analysis

As mentioned in the main manuscript, we only included patients within the Berlin CIS-Cohort [1], who had a diagnosis of either Clinically Isolated Syndrome (CIS) within six months from symptom onset or of Relapsing-Remitting Multiple Sclerosis (RRMS) within two years from symptom onset, according to the 2017 revisions of the McDonald criteria [2]. Study data on patients with CIS/RRMS (PwMS) and their matched healthy controls (HC) were collected and managed using Research Electronic Data Capture (REDCap) tools hosted at the Charité — Universitätsmedizin Berlin, Berlin, Germany [3,4].

To address the disease activity and progression of PwMS, we collected data on the number of relapses and time to last relapse, as well as expanded disability status scale (EDSS) [5] and Multiple Sclerosis functional composite (MSFC) [6] scores. For MSFC Z-scores, we used the data collected at the baseline visit (VIS1) in this study cohort as reference.

Regarding MRI data collection and analysis, we used the same methodology as the one employed in a previous study [7]. All MRI data were acquired using two 3 T machines (Tim Trio; Siemens, Erlangen, Germany). The scanning protocol included a 3-dimensional (3D) T1-weighted magnetization prepared rapid acquisition gradient echo (MPRAGE) sequence (repetition time [TR] = 1,900 milliseconds, echo time [TE] = 2.55 milliseconds, inversion time [TI] = 900 milliseconds) for pre-contrast and post-contrast administration and 3D T2-weighted (T2w) fluid-attenuated inversion recovery (FLAIR) sequence (TR/TE/TI = 6,000/388/2,100 milliseconds; $1 \times 1 \times 1$ $mm^3$ resolution).

For brain lesion segmentation, pre-contrast and post-contrast MPRAGE images were co-registered to MNI-152 standard space for all patients using FLIRT from FSL with linear-rigid registration [8,9]. Subsequently, FLAIR images were co-registered with pre-contrast MPRAGE images. Brain T2-hyperintense lesions were manually segmented using FLAIR to create binary masks by 2 expert raters under the supervision of a board-certified radiologist using ITK-SNAP (itksnap.org) [10,11]. Lesion mask measures were corrected by thresholding each lesion to be at least 15 voxels to reduce chances of human error or non-specific white matter lesion detection. Gd+ lesions were segmented using post-contrast MPRAGEs by the same raters and ITK-SNAP.

Similarly, for optical coherence tomography (OCT) data collection and analysis, we used the same methodology as the one employed in a previous study [7]. All OCT images were obtained under normal room light conditions by experienced operators through spectral domain OCT and Heidelberg Eye Explorer (HeyEx) version 1.9.10.0 (Heidelberg Engineering, Heidelberg, Germany) with automatic real-time (ART) function for image averaging and an activated eye tracker. All individual scans were quality controlled according to the OSCAR-IB criteria [12,13] and reported based on the APOSTEL recommendations [14]. A peripapillary circular scan within a 12° (~3.4 mm) diameter ring (1,536 A-scans; $9 \leq ART \leq 99$) around the optic nerve head was used to measure the peripapillary retinal nerve fiber layer (RNFL) thickness. The RNFL on ring scans was segmented with HeyEx viewing module version 6.0.14.0. Macular volume scans (25° × 30°, 61 vertical B-scans, 12 ≤ ART ≤ 18) were segmented with the SAMIRIX pipeline [15]. The ganglion cell and inner plexiform layer (GCIPL) and inner nuclear layer (INL) thickness were calculated as a 6-mm diameter cylinder around the fovea [16]. Segmentation errors were manually corrected if necessary. We calculated mean RNFL thickness and GCIPL volume for both eyes per clinical visit; single-eye assessments were excluded.

### Section 2 — CD4+ T cell enrichment and flow cytometry analysis

#### 2.1. Overall CD4+ T cell enrichment protocol

As mentioned in the main manuscript, sample processing was performed at the same time for each triplet (PwMS at VIS1 & visit 2 [VIS2] and HC), to minimize differences within processing. Briefly, samples with peripheral blood mononuclear cells (PBMCs) were stored at -80° C prior to processing. They were then thawed in a pre-warmed water bath at 37° C and washed with pre-warmed (at 37° C) PBS$^{-/-}$ containing FBS at 2% (PBS+FBS). Following a centrifugation at 300 x *g* for 5 min at 4° C and supernatant removal, cells were passed through a Falcon® 70 µm cell strainer (#352350, Corning Inc., NY, USA) and counted manually with 0.4% Trypan blue (#T8154, Sigma-Aldrich®, Merck KGaA, Darmstadt, Germany), in a 1:1 ratio, with the aid of a Neubauer chamber.

An aliquot was taken for flow cytometry analysis if the total number of cells was > 10.5 million cells. If feasible, a particular attempt was made to take an aliquot per subject type on each day, where the sample with the highest number of cells was selected. This aliquot would then be stored at 4° C until further processing.

After cell counting and removal of an aliquot for flow cytometry analysis, samples were further centrifuged at 300 x *g* for 5 min at 4° C, and the supernatant was discarded. Afterwards, CD4+ T cell enrichment with the MojoSort™ Human CD4 T Cell Isolation Kit (#480010, BioLegend, San Diego, CA, USA) ensued, according to the manufacturer's instructions. Following the magnetic enrichment, cell counting was performed once more, while samples were kept at 4° C.

If the number of cells was > 2 million, particularly if an aliquot for flow cytometry analysis was taken before the magnetic enrichment, a further aliquot was taken and kept at 4° C until further processing.

Subsequently, for storage, samples were centrifuged at 300 x *g* for 5 min at 4° C, followed by supernatant removal and the addition of freezing solution at 4° C (80% PBS+FBS, 10% FBS, 10% DMSO). Finally, samples were placed in a Nalgene® Mr. Frosty (Sigma-Aldrich®, Merck KGaA, Darmstadt, Germany) for a minimum of 4 h at -80° C until they were transported to long-term storage in an ultrafreezer, kept at the same temperature.

#### 2.2. CD4+ T cell enrichment optimization

For CD4+ T cell enrichment, we tested two different protocols: (**I**) a positive selection, by first depleting monocytes with the MojoSort™ Human CD14 Selection Kit (#480025, BioLegend, San Diego, CA, USA), followed by a capture of labelled cells with the MojoSort™ Human CD4 Nanobeads (#480014, BioLegend, San Diego, CA, USA), as suggested by the manufacturer [17]; and (**II**) a negative selection with the MojoSort™ Human CD4 T Cell Isolation Kit (#480010, BioLegend, San Diego, CA, USA).

In addition to the antibodies listed in the main manuscript, we initially used:

- CD123-FITC (#11123942, clone 6H6, eBioscience™, Thermo Fisher Scientific, Waltham, MA, USA);
- Fixable Viability Dye eFluor™ 506 (#650866, eBioscience™, Thermo Fisher Scientific, Waltham, MA, USA).

After comparing the two options from the same parent sample (Table S1), according to the temporary gating strategy (Figure S1), as well as considering the main contaminants in the enrichment protocols (Figure S2), the MojoSort™ Human CD4 T Cell Isolation Kit yielded more events in flow cytometry and a larger percentage of viable CD4$^+$ T cells. Thus, we decided to: (**I**) use the MojoSort™ Human CD4 T Cell Isolation Kit for CD4$^+$ T cell enrichment; and (**II**) eliminate CD123 from the staining protocol, as it was the least relevant contaminant.

#### 2.3. Flow cytometry optimization and protocol

To infer the exact number of cells needed for the flow cytometry analysis and the optimal strategy of how cells should be fixed (before or after surface staining), we tried eight different cell amounts in duplicate (Figure S3): (**I**) with staining before fixation; and (**II**) with staining after fixation. After analyzing the data (Table S2), according to a temporary gating strategy (Figure S4), fixing the cells prior to staining and a minimum of 300,000 cells was determined as optimal. Consequently, in order to account for cell loss in sample processing, 375,000 cells were used for flow cytometry analysis.

Initially, cells were frozen prior to fixation and stored in the same freezing solution described previously. On the day of flow cytometry analysis, cells were quickly thawed and washed with PBS+FBS, as in *2.1. Overall CD4+ T cell enrichment protocol*. Afterwards, fixation was performed with 2% paraformaldehyde (PFA). Samples would be then washed with PBS$^{-/-}$, followed by a centrifugation at 1000 x *g* for 5 min at 4° C, and supernatant removal. Then, the antibody mixture was added, followed by an incubation protected from light for 30 min at 4° C. After the incubation, samples were again washed with PBS$^{-/-}$ and centrifuged at 1000 x *g* for 5 min at 4° C. The supernatant would then be removed, and cells were resuspended in PBS$^{-/-}$ for flow cytometry analysis.

However, since cell loss was high and the number of recorded events in the flow cytometer was highly variable (Table S3), fixation was performed immediately after finishing the magnetic enrichment and kept at 4° C. The aforementioned staining protocol was then performed on the day of the flow cytometry analysis. Samples were left unanalyzed for a maximum of five days for freshly fixed samples and 11 days for previously frozen samples. Out of a total of 132 suitable aliquots analyzed through flow cytometry, only 23 samples were previously frozen (Table S3).

Flow cytometry acquisition was performed until 30,000 events or 10 min had passed (whichever occurred first). Rate was kept at "low" unless < 10,500 events were acquired at 3.5 min; if so, the rate was increased to "medium".

Regarding the gating strategy (Figure S5 and Figure S6), FSC-A *vs.* FSC-H followed by SSC-A *vs.* SSC-H, were used to select single cells, followed by a gating on $CD3^+$ cells, then $CD4^+$ cells, and, finally, on $CD19^-$ and $CD56^-$ cells. CD14 was excluded from the gating strategy, as $CD3^+CD4^+CD14^+$ cells likely represent doublets of $CD4^+$ T cells associated with monocytes and not a distinct cell lineage [18]. Also, we did not take into account cell viability in the flow cytometry analysis, since mitochondrial DNA (mtDNA) would be extracted from the samples without previous isolation of live cells and no functional assays would take place.

Four different batch gating strategies with FlowJo™ v10 (BD Biosciences, Franklin Lakes, NJ, USA) were adopted: (**I**) cells frozen prior to fixation, processed in 2018 (Figure S5A–B); (**II**) cells freshly fixed, processed in 2018 (Figure S5C–D); (**III**) cells freshly fixed, processed in 2019, before a scheduled maintenance on the BD LSRFortessa™ X-20 Cell Analyzer on August 12, 2019, where a blue laser was substituted and the flow cell was optimized (Figure S6A–B); and (**IV**) cells freshly fixed, processed in 2019, after the scheduled maintenance (Figure S6C–D).

Nonetheless, there were a few exceptions to samples processed in 2019, which are highlighted in Table S3, namely:

- On May 23, 2019, the MojoSort™ Human CD14 Selection Kit was used by mistake instead of the MojoSort™ Human CD4 T Cell Isolation Kit. Cells were kept overnight at 4° C and underwent the proper $CD4^+$ T cell enrichment protocol the next day. Even though $CD4^+$ T cells increased afterwards, CD4 signal was significantly reduced in these samples. Thus, these samples were excluded from the flow cytometry analysis;
- On July 25, 2019, one of the seven analyzed samples had an inexplicably low CD4 signal. This sample was excluded from the flow cytometry analysis;
- On August 23, 2019, the samples fit better with the batch gating strategy aimed at cells processed prior to the scheduled maintenance on August 12, 2019. We used the previous gating strategy and these samples were included in the flow cytometry analysis.

#### 2.4. Figures and Tables

#### Table S1 — $CD4^+$ T cell enrichment trial: Summary

| | Manual cell count with Neubauer chamber | | | Flow cytometry analysis | | | | |
|---|---|---|---|---|---|---|---|---|
| **Source** | Number of cells (millions) | Viable cells (%) | From input (%) | Number of events | Single cells (%) | Viable cells (%) | Viable T cells ($CD3^+$) (%) | Viable $CD4^+$ T cells ($CD3^+CD4^+CD14^-$ $CD19^-$) (%) |
| Input | 6,650,000 | 85.71% | - | 10000 | 81.46% | 87.74% | 46.31% | 24.26% |
| Output selection + | 453,333 | 94.12% | 15.49% | 2012 | 52.39% | 90.23% | 42.17% | 16.82% |
| Output selection - | 720,000 | 93.06% | 24.61% | 20000 | 88.94% | 94.63% | 83.77% | 62.85% |

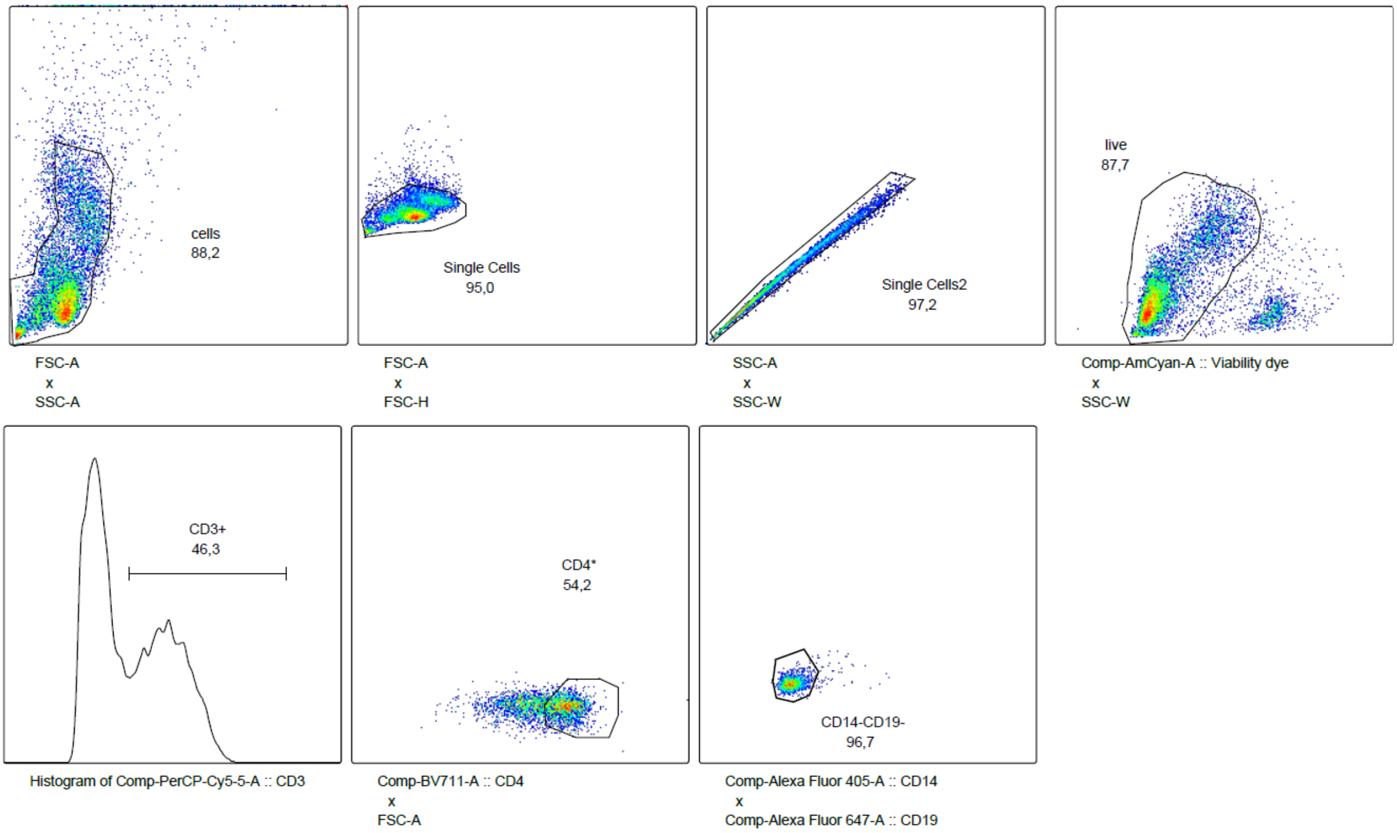

#### Figure S1 — Temporary gating strategy in the CD4+ T cell enrichment trial

The sample represented is the parent sample (input) from where $CD4^+$ T cell enrichment occurred. The order is: FSC-A *vs.* SSC-A, FSC-A *vs.* FSC-H, SSC-A *vs.* SSC-W, Fixable Viability Dye eFluor™ 506 *vs.* SSC-W, CD3-PerCP-Cy5.5 (histogram), CD4-BV711 *vs.* FSC-A, and CD14-eFluor450 *vs.* CD19-Alexa Fluor 647. The gaiting is aimed at identifying viable $CD4^+$ T cells ($CD3^+CD4^+CD14^-CD19^-$).

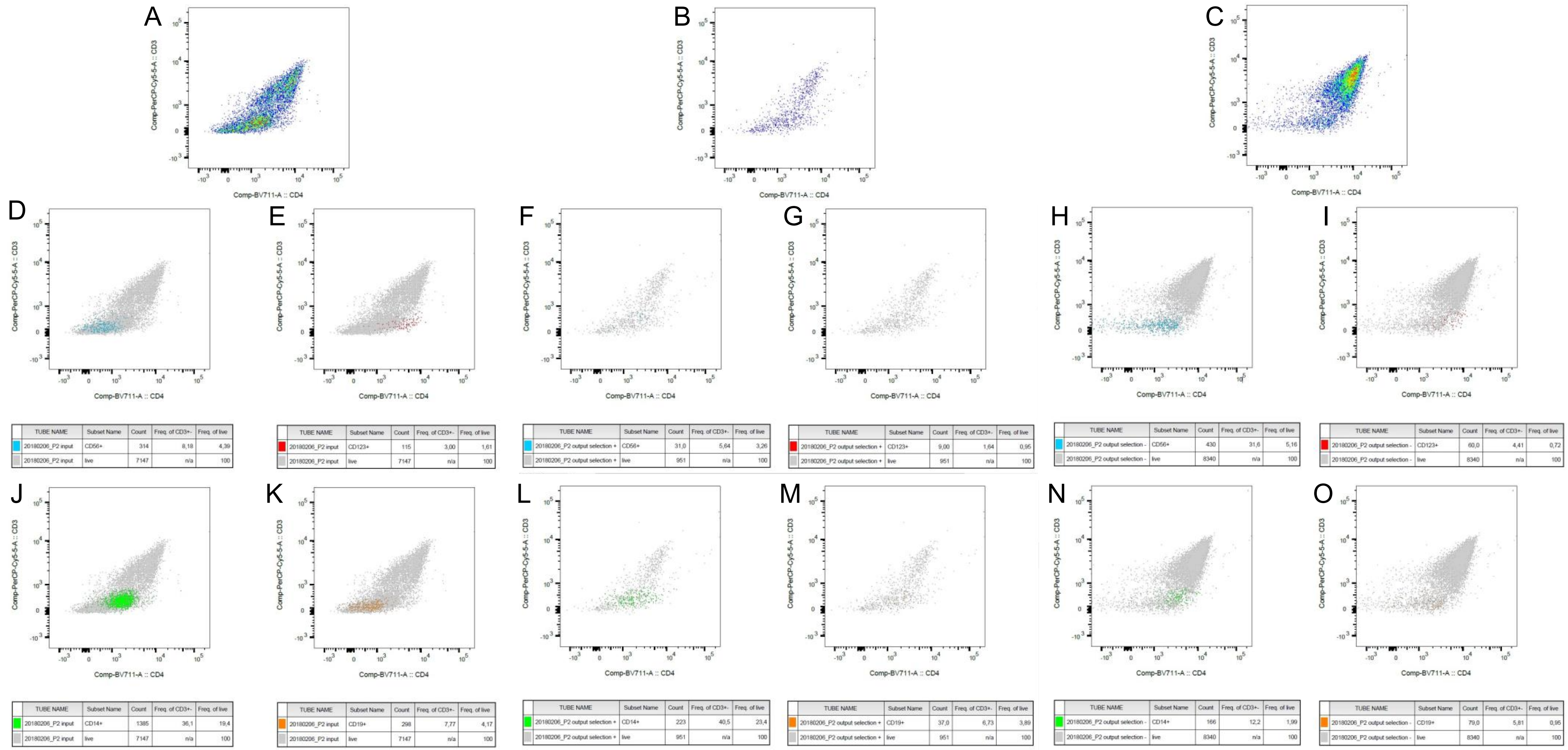

**Figure S2 — Analysis of the contaminants in the CD4⁺ T cell enrichment trial**

(**A,B,C**) Viable CD3⁺ cells (gating strategy in Figure S1) plotted with CD4-BV711 *vs.* CD3-PerCP-Cy5.5 from the parent sample, the positive selection, and the negative selection, respectively; (**D,F,H**) CD3⁺CD56⁺ cells from the parent sample, the positive selection, and the negative selection, respectively; (**E,G,I**) CD3⁺CD123⁺ cells from the parent sample, the positive selection, and the negative selection, respectively; (**J,L,N**) CD3⁺CD14⁺ cells from the parent sample, the positive selection, and the negative selection, respectively; (**K,M,O**) CD3⁺CD19⁺ cells from the parent sample, the positive selection, and the negative selection, respectively.

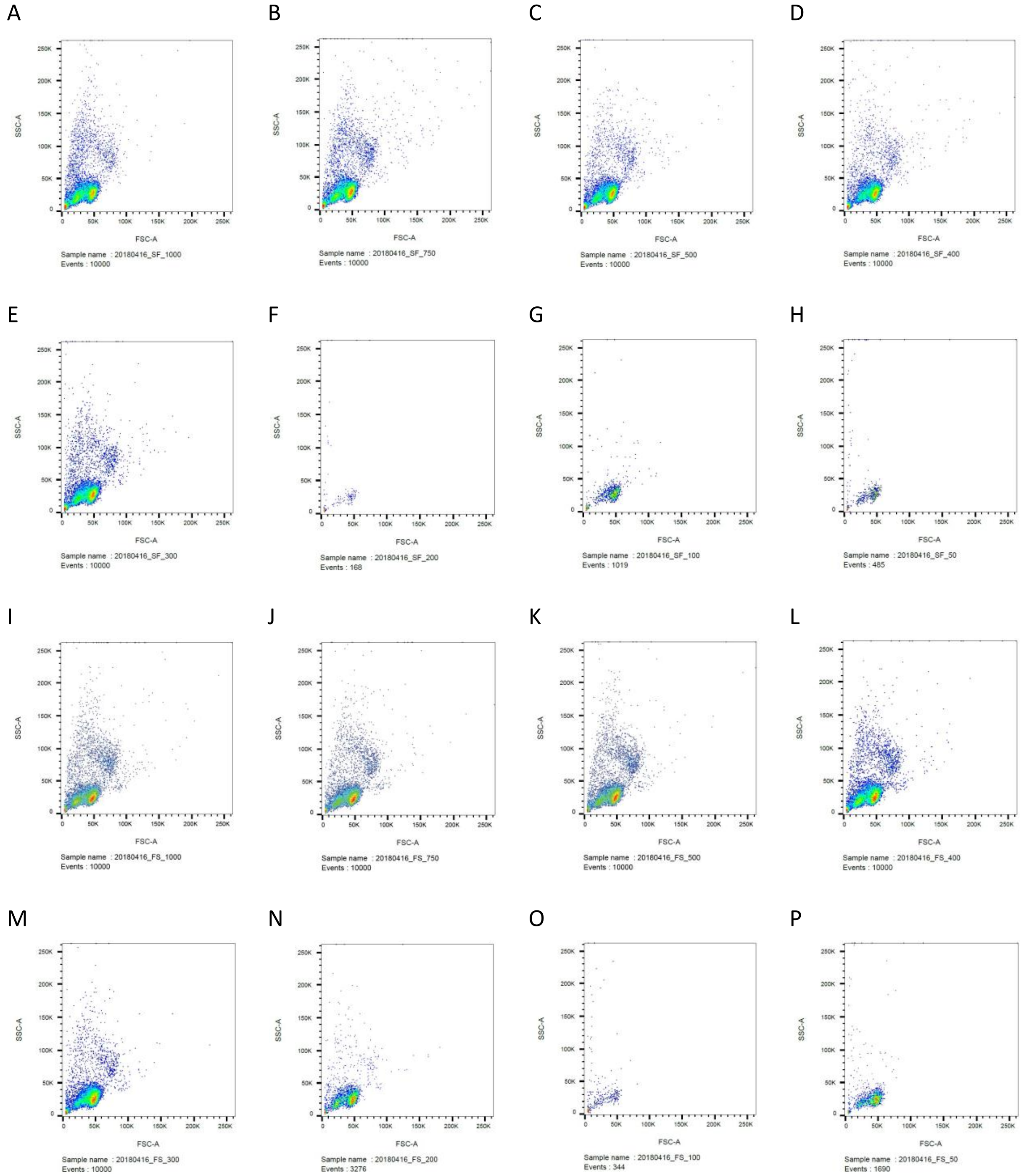

#### Figure S3 — Staining and fixation protocol optimization, with multiple cell amounts

(**A–H**) Samples analyzed with flow cytometry, with staining before fixation, with 1,000,000 cells, 750,000 cells, 500,000 cells, 400,000 cells, 300,000 cells, 200,000 cells, 100,000 cells, and 50,000 cells, respectively; (**I–P**) Samples analyzed with flow cytometry, with fixation before staining, with 1,000,000 cells, 750,000 cells, 500,000 cells, 400,000 cells, 300,000 cells, 200,000 cells, 100,000 cells, and 50,000 cells, respectively.

#### Table S2 — Staining and fixation protocol optimization: Summary

| Method | Number of cells (x1000) | Number of events | Single cells (%) | CD4+ T cells ($CD3^+CD4^+CD56^-CD19^-$) (%) |
|---|---|---|---|---|
| Stain-Fix | All | 6459 | 96.42% | 27.83% |
| Stain-Fix | ≥300 | 10000 | 96.35% | 27.10% |
| Stain-Fix | <300 | 557.33 | 96.54% | 29.07% |
| Stain-Fix | 50 | 485 | 95.05% | 35.14% |
| Stain-Fix | 100 | 1019 | 98.14% | 28.60% |
| Stain-Fix | 200 | 168 | 96.43% | 23.46% |
| Stain-Fix | 300 | 10000 | 97.17% | 25.48% |
| Stain-Fix | 400 | 10000 | 96.93% | 26.21% |
| Stain-Fix | 500 | 10000 | 97.05% | 29.35% |
| Stain-Fix | 750 | 10000 | 94.29% | 28.35% |
| Stain-Fix | 1000 | 10000 | 96.30% | 26.09% |
| Fix-Stain | All | 6914 | 96.95% | 29.25% |
| Fix-Stain | ≥300 | 10000 | 96.78% | 31.49% |
| Fix-Stain | <300 | 1770 | 97.24% | 25.52% |
| Fix-Stain | 50 | 1690 | 98.22% | 28.98% |
| Fix-Stain | 100 | 344 | 95.35% | 13.72% |
| Fix-Stain | 200 | 3276 | 98.14% | 33.87% |
| Fix-Stain | 300 | 10000 | 98.27% | 28.56% |
| Fix-Stain | 400 | 10000 | 96.67% | 32.99% |
| Fix-Stain | 500 | 10000 | 96.68% | 31.32% |
| Fix-Stain | 750 | 10000 | 96.80% | 33.73% |
| Fix-Stain | 1000 | 10000 | 95.46% | 30.85% |

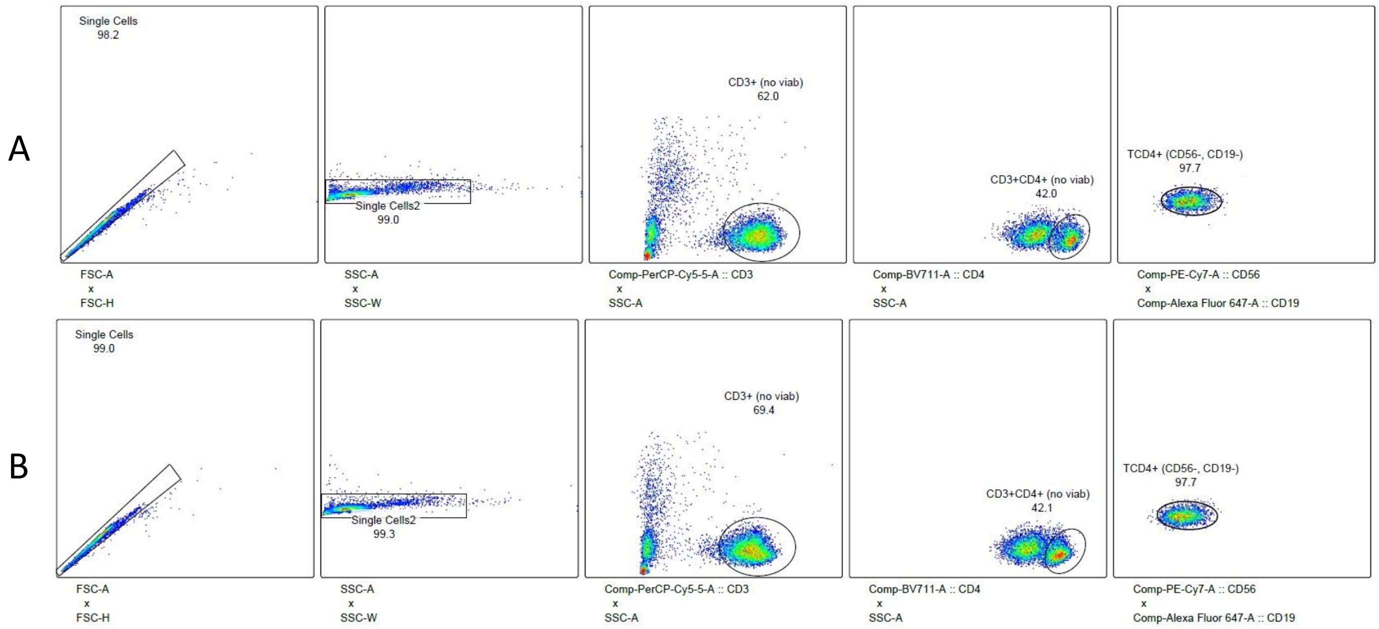

#### Figure S4 — Staining and fixation protocol optimization: Temporary gating strategy

The order is: FSC-A *vs*. FSC-H, SSC-A *vs*. SSC-W, CD3-PerCP-Cy5.5 *vs.* SSC-A, CD4-BV711 *vs.* SSC-A, and CD56-PE-Cy7 *vs.* CD19-Alexa Fluor 647. The gaiting is aimed at identifying CD4+ T cells (CD3+CD4+CD56-CD19-). (**A**) Gating strategy for sample with 300,000 cells that was stained before fixation; (**B**) Gating strategy for sample with 300,000 cells that was fixed before staining.

#### Table S3 — Samples analyzed with flow cytometry

| Subject Type | ID | Separation date | Analysis date | Input or output? | Frozen? | Number of single cells | Single cells (%) | $CD4^+$ T cells ($CD3^+CD4^+CD19^-CD56^-$) |
|---|---|---|---|---|---|---|---|---|
| HC | HC_2 | 21/04/2018 | 02/05/2018 | input | Yes | 7710 | 96.58% | 31.21% |
| HC | HC_2 | 21/04/2018 | 02/05/2018 | output | Yes | 3806 | 97.89% | 73.33% |
| V1 | V1_1 | 21/04/2018 | 02/05/2018 | output | Yes | 964 | 92.78% | 40.77% |
| V1 | V1_2 | 21/04/2018 | 02/05/2018 | input | Yes | 1815 | 95.58% | 34.05% |
| V1 | V1_2 | 21/04/2018 | 02/05/2018 | output | Yes | 2631 | 96.48% | 64.31% |
| V2 | V2_1 | 21/04/2018 | 02/05/2018 | output | Yes | 8615 | 98.61% | 48.52% |
| HC | HC_3 | 22/04/2018 | 02/05/2018 | input | Yes | 5280 | 98.40% | 24.41% |
| HC | HC_3 | 22/04/2018 | 02/05/2018 | output | Yes | 5389 | 94.86% | 54.85% |
| V1 | V1_4 | 22/04/2018 | 02/05/2018 | input | Yes | 2074 | 94.62% | 27.15% |
| V1 | V1_4 | 22/04/2018 | 02/05/2018 | output | Yes | 5895 | 99.48% | 79.56% |
| V2 | V2_3 | 22/04/2018 | 02/05/2018 | input | Yes | 880 | 83.41% | 15.23% |
| V2 | V2_3 | 22/04/2018 | 02/05/2018 | output | Yes | 4145 | 97.76% | 43.23% |
| HC | HC_5 | 29/04/2018 | 10/05/2018 | input | Yes | 28650 | 95.50% | 22.02% |
| HC | HC_5 | 29/04/2018 | 10/05/2018 | output | Yes | 27878 | 92.93% | 54.23% |
| V1 | V1_5 | 29/04/2018 | 10/05/2018 | input | Yes | 29135 | 97.12% | 27.43% |
| V1 | V1_5 | 29/04/2018 | 10/05/2018 | output | Yes | 27296 | 90.99% | 47.00% |
| V2 | V2_5 | 29/04/2018 | 10/05/2018 | input | Yes | 28857 | 96.19% | 30.25% |
| V2 | V2_5 | 29/04/2018 | 10/05/2018 | output | Yes | 28329 | 94.43% | 65.44% |
| HC | HC_8 | 30/04/2018 | 10/05/2018 | input | Yes | 29278 | 97.59% | 23.37% |
| HC | HC_8 | 30/04/2018 | 10/05/2018 | output | Yes | 27533 | 91.78% | 60.54% |
| V1 | V1_8 | 30/04/2018 | 10/05/2018 | output | Yes | 29203 | 97.34% | 72.52% |
| V2 | V2_8 | 30/04/2018 | 10/05/2018 | input | Yes | 29205 | 97.35% | 25.84% |
| V2 | V2_8 | 30/04/2018 | 10/05/2018 | output | Yes | 27330 | 91.10% | 54.53% |
| HC | HC_9 | 05/05/2018 | 09/05/2018 | input | | 28338 | 94.46% | 40.09% |
| HC | HC_9 | 05/05/2018 | 09/05/2018 | output | | 28675 | 95.57% | 72.92% |
| V1 | V1_10 | 05/05/2018 | 09/05/2018 | input | | 29151 | 96.31% | 52.55% |
| V1 | V1_10 | 05/05/2018 | 09/05/2018 | output | | 28842 | 95.95% | 77.90% |
| V2 | V2_10 | 05/05/2018 | 09/05/2018 | input | | 29178 | 96.39% | 46.75% |
| V2 | V2_10 | 05/05/2018 | 09/05/2018 | output | | 28957 | 96.52% | 70.50% |
| HC | HC_11 | 06/05/2018 | 09/05/2018 | input | | 28486 | 94.95% | 42.79% |
| HC | HC_11 | 06/05/2018 | 09/05/2018 | output | | 28187 | 93.96% | 74.96% |
| V1 | V1_11 | 06/05/2018 | 09/05/2018 | input | | 28441 | 94.80% | 46.46% |
| V1 | V1_11 | 06/05/2018 | 09/05/2018 | output | | 28095 | 93.65% | 72.96% |
| V2 | V2_11 | 06/05/2018 | 09/05/2018 | input | | 28780 | 95.93% | 53.84% |
| V2 | V2_11 | 06/05/2018 | 09/05/2018 | output | | 29009 | 96.70% | 81.09% |
| HC | HC_13 | 11/05/2018 | 16/05/2018 | input | | 28813 | 96.04% | 27.50% |
| HC | HC_13 | 11/05/2018 | 16/05/2018 | output | | 28516 | 95.05% | 56.63% |
| V1 | V1_13 | 11/05/2018 | 16/05/2018 | input | | 27929 | 93.10% | 45.06% |
| V1 | V1_13 | 11/05/2018 | 16/05/2018 | output | | 28671 | 95.57% | 60.23% |
| V2 | V2_13 | 11/05/2018 | 16/05/2018 | input | | 28936 | 96.45% | 48.29% |
| V2 | V2_13 | 11/05/2018 | 16/05/2018 | output | | 29501 | 98.34% | 62.55% |
| HC | HC_16 | 12/05/2018 | 16/05/2018 | input | | 20151 | 98.24% | 30.44% |
| HC | HC_16 | 12/05/2018 | 16/05/2018 | output | | 28319 | 94.40% | 61.22% |

#### Table S3 — Samples analyzed with flow cytometry (continued)

| Subject Type | ID | Separation date | Analysis date | Input or output? | Frozen? | Number of single cells | Single cells (%) | CD4+ T cells ($CD3^{+}CD4^{+}CD19^{-}CD56^{-}$) |
|---|---|---|---|---|---|---|---|---|
| V1 | V1_16 | 12/05/2018 | 16/05/2018 | input | | 28731 | 95.77% | 39.76% |
| V1 | V1_16 | 12/05/2018 | 16/05/2018 | output | | 29105 | 97.02% | 72.97% |
| V2 | V2_15 | 12/05/2018 | 16/05/2018 | input | | 29315 | 97.72% | 38.35% |
| V2 | V2_15 | 12/05/2018 | 16/05/2018 | output | | 28750 | 95.83% | 64.70% |
| V1 | V1_17 | 14/05/2018 | 19/05/2018 | input | | 29160 | 97.20% | 52.43% |
| V1 | V1_17 | 14/05/2018 | 19/05/2018 | output | | 28894 | 96.21% | 79.26% |
| V2 | V2_18 | 14/05/2018 | 19/05/2018 | input | | 29685 | 98.95% | 43.85% |
| V2 | V2_18 | 14/05/2018 | 19/05/2018 | output | | 29595 | 98.65% | 66.31% |
| HC | HC_20 | 15/05/2018 | 19/05/2018 | input | | 28901 | 96.34% | 28.02% |
| HC | HC_20 | 15/05/2018 | 19/05/2018 | output | | 27965 | 93.22% | 61.93% |
| V1 | V1_20 | 15/05/2018 | 19/05/2018 | input | | 29396 | 97.99% | 44.18% |
| V1 | V1_20 | 15/05/2018 | 19/05/2018 | output | | 29072 | 96.91% | 71.38% |
| V2 | V2_19 | 15/05/2018 | 19/05/2018 | input | | 29210 | 97.37% | 33.09% |
| V2 | V2_19 | 15/05/2018 | 19/05/2018 | output | | 28344 | 94.48% | 66.30% |
| HC | HC_21 | 24/05/2019 | 27/05/2019 | AfterCD4 | | 25757 | 97.23% | 70.61% |
| HC | HC_21 | 24/05/2019 | 27/05/2019 | AfterCD14 | | 32958 | 95.08% | 3.23% |
| HC | HC_21 | 24/05/2019 | 27/05/2019 | BeforeCD14 | | 48928 | 95.04% | 1.28% |
| HC | HC_57 | 24/05/2019 | 27/05/2019 | AfterCD4 | | 30278 | 98.17% | 52.58% |
| HC | HC_57 | 24/05/2019 | 27/05/2019 | AfterCD14 | | 29016 | 95.77% | 0.26% |
| HC | HC_57 | 24/05/2019 | 27/05/2019 | BeforeCD14 | | 22499 | 95.72% | 0.48% |
| V1 | V1_21 | 24/05/2019 | 27/05/2019 | AfterCD4 | | 9198 | 95.42% | 54.40% |
| V1 | V1_21 | 24/05/2019 | 27/05/2019 | AfterCD14 | | 24624 | 93.99% | 2.27% |
| V2 | V2_21 | 24/05/2019 | 27/05/2019 | AfterCD14 | | 32566 | 95.81% | 1.16% |
| HC | HC_38 | 29/05/2019 | 02/06/2019 | input | | 50769 | 94.23% | 21.78% |
| HC | HC_38 | 29/05/2019 | 02/06/2019 | output | | 31727 | 97.03% | 73.64% |
| V1 | V1_45 | 30/05/2019 | 02/06/2019 | output | | 30923 | 98.35% | 91.39% |
| V2 | V2_45 | 30/05/2019 | 02/06/2019 | input | | 55499 | 93.69% | 34.42% |
| V2 | V2_45 | 30/05/2019 | 02/06/2019 | output | | 32563 | 96.11% | 81.68% |
| HC | HC_35 | 31/05/2019 | 02/06/2019 | output | | 18786 | 96.34% | 71.20% |
| V1 | V1_50 | 31/05/2019 | 02/06/2019 | output | | 33237 | 91.62% | 59.46% |
| V2 | V2_35 | 31/05/2019 | 02/06/2019 | output | | 8887 | 92.34% | 35.06% |
| HC | HC_44 | 01/06/2019 | 02/06/2019 | input | | 56248 | 95.44% | 15.15% |
| HC | HC_44 | 01/06/2019 | 02/06/2019 | output | | 34832 | 92.00% | 54.31% |
| V1 | V1_44 | 01/06/2019 | 02/06/2019 | output | | 37810 | 91.05% | 57.32% |
| V2 | V2_44 | 01/06/2019 | 02/06/2019 | output | | 38185 | 88.14% | 51.84% |
| HC | HC_55 | 16/06/2019 | 17/06/2019 | input | | 49031 | 94.84% | 24.91% |
| HC | HC_55 | 16/06/2019 | 17/06/2019 | output | | 32833 | 93.50% | 74.18% |
| V1 | V1_55 | 16/06/2019 | 17/06/2019 | output | | 14810 | 82.85% | 32.78% |
| V2 | V2_47 | 16/06/2019 | 17/06/2019 | output | | 19463 | 96.36% | 52.10% |
| HC | HC_22 | 19/06/2019 | 21/06/2019 | output | | 31154 | 95.79% | 78.74% |
| HC | HC_58 | 19/06/2019 | 21/06/2019 | input | | 62526 | 93.74% | 15.66% |
| HC | HC_58 | 19/06/2019 | 21/06/2019 | output | | 35522 | 93.78% | 64.43% |
| V1 | V1_22 | 19/06/2019 | 21/06/2019 | output | | 30529 | 98.47% | 84.92% |
| V2 | V2_58 | 19/06/2019 | 21/06/2019 | output | | 37227 | 91.16% | 52.74% |

#### Table S3 — Samples analyzed with flow cytometry (continued)

| Subject Type | ID | Separation date | Analysis date | Input or output? | Frozen? | Number of single cells | Single cells (%) | CD4⁺ T cells ($CD3^+CD4^+CD19^-CD56^-$) |
|---|---|---|---|---|---|---|---|---|
| HC | HC_24 | 20/06/2019 | 21/06/2019 | output | | 31725 | 97.43% | 82.71% |
| HC | HC_61 | 20/06/2019 | 21/06/2019 | input | | 74544 | 96.13% | 18.34% |
| HC | HC_61 | 20/06/2019 | 21/06/2019 | output | | 32200 | 96.25% | 79.86% |
| V1 | V1_61 | 20/06/2019 | 21/06/2019 | output | | 32298 | 96.06% | 80.43% |
| V2 | V2_24 | 20/06/2019 | 21/06/2019 | output | | 34559 | 93.56% | 66.69% |
| HC | HC_59 | 26/06/2019 | 28/06/2019 | input | | 58174 | 94.81% | 15.33% |
| HC | HC_59 | 26/06/2019 | 28/06/2019 | output | | 34640 | 90.20% | 60.82% |
| V1 | V1_39 | 26/06/2019 | 28/06/2019 | output | | 25663 | 92.68% | 74.52% |
| V2 | V2_59 | 26/06/2019 | 28/06/2019 | input | | 67924 | 96.45% | 20.71% |
| V2 | V2_59 | 26/06/2019 | 28/06/2019 | output | | 32369 | 95.82% | 78.11% |
| HC | HC_23 | 27/06/2019 | 28/06/2019 | input | | 70070 | 97.45% | 14.27% |
| HC | HC_23 | 27/06/2019 | 28/06/2019 | output | | 37649 | 89.69% | 59.52% |
| V1 | V1_42 | 27/06/2019 | 28/06/2019 | output | | 32743 | 93.61% | 79.21% |
| V2 | V2_42 | 27/06/2019 | 28/06/2019 | input | | 62336 | 96.37% | 23.52% |
| V2 | V2_42 | 27/06/2019 | 28/06/2019 | output | | 34279 | 94.66% | 74.26% |
| HC | HC_40 | 02/07/2019 | 04/07/2019 | input | | 74232 | 94.16% | 14.97% |
| HC | HC_40 | 02/07/2019 | 04/07/2019 | output | | 35794 | 90.91% | 62.81% |
| V1 | V1_40 | 02/07/2019 | 04/07/2019 | output | | 33408 | 93.38% | 79.15% |
| HC | HC_51 | 03/07/2019 | 04/07/2019 | output | | 35464 | 92.86% | 68.45% |
| HC | HC_52 | 03/07/2019 | 04/07/2019 | input | | 69810 | 96.93% | 16.18% |
| HC | HC_52 | 03/07/2019 | 04/07/2019 | output | | 33785 | 95.47% | 72.98% |
| V1 | V1_51 | 03/07/2019 | 04/07/2019 | output | | 37565 | 90.56% | 58.81% |
| V1 | V1_52 | 03/07/2019 | 04/07/2019 | input | | 81832 | 95.85% | 18.61% |
| V1 | V1_52 | 03/07/2019 | 04/07/2019 | output | | 33064 | 95.45% | 80.65% |
| V2 | V2_51 | 03/07/2019 | 04/07/2019 | output | | 34099 | 91.09% | 71.45% |
| V2 | V2_52 | 03/07/2019 | 04/07/2019 | output | | 31346 | 97.01% | 87.97% |
| HC | HC_29 | 17/07/2019 | 19/07/2019 | input | | 55974 | 94.10% | 22.61% |
| HC | HC_29 | 17/07/2019 | 19/07/2019 | output | | 33272 | 95.16% | 72.09% |
| V2 | V2_29 | 17/07/2019 | 19/07/2019 | output | | 31799 | 96.03% | 80.23% |
| HC | HC_28 | 18/07/2019 | 19/07/2019 | input | | 55953 | 93.98% | 19.24% |
| HC | HC_28 | 18/07/2019 | 19/07/2019 | output | | 33869 | 95.32% | 69.72% |
| V1 | V1_28 | 18/07/2019 | 19/07/2019 | output | | 51012 | 89.98% | 45.26% |
| V2 | V2_41 | 18/07/2019 | 19/07/2019 | output | | 33883 | 92.51% | 74.87% |
| HC | HC_53 | 20/07/2019 | 25/07/2019 | input | | 48227 | 94.13% | 33.17% |
| HC | HC_53 | 20/07/2019 | 25/07/2019 | output | | 31770 | 96.35% | 72.55% |
| V1 | V1_53 | 20/07/2019 | 25/07/2019 | output | | 34281 | 93.27% | 34.56% |
| HC | HC_60 | 21/07/2019 | 25/07/2019 | input | | 50347 | 92.08% | 20.42% |
| HC | HC_60 | 21/07/2019 | 25/07/2019 | output | | 34385 | 89.20% | 60.26% |
| V1 | V1_60 | 21/07/2019 | 25/07/2019 | output | | 34905 | 92.43% | 60.41% |
| V2 | V2_60 | 21/07/2019 | 25/07/2019 | output | | 33070 | 95.94% | 58.88% |
| HC | HC_31 | 15/08/2019 | 19/08/2019 | input | | 38484 | 97.95% | 27.26% |
| HC | HC_31 | 15/08/2019 | 19/08/2019 | output | | 32284 | 98.60% | 65.57% |
| V2 | V2_31 | 15/08/2019 | 19/08/2019 | output | | 10155 | 98.33% | 64.30% |
| HC | HC_26 | 16/08/2019 | 19/08/2019 | input | | 33807 | 94.12% | 27.61% |

#### Table S3 — Samples analyzed with flow cytometry (continued)

| Subject Type | ID | Separation date | Analysis date | Input or output? | Frozen? | Number of single cells | Single cells (%) | CD4$^+$ T cells (CD3$^+$CD4$^+$CD19$^-$CD56$^-$) |
|---|---|---|---|---|---|---|---|---|
| HC | HC_26 | 16/08/2019 | 19/08/2019 | output | | 31716 | 95.38% | 74.78% |
| V1 | V1_26 | 16/08/2019 | 19/08/2019 | output | | 30452 | 98.47% | 84.11% |
| HC | HC_54 | 17/08/2019 | 19/08/2019 | input | | 39576 | 98.84% | 18.07% |
| HC | HC_54 | 17/08/2019 | 19/08/2019 | output | | 30936 | 96.60% | 76.87% |
| V1 | V1_32 | 17/08/2019 | 19/08/2019 | input | | 39072 | 96.79% | 26.54% |
| V1 | V1_32 | 17/08/2019 | 19/08/2019 | output | | 31878 | 95.89% | 68.53% |
| V1 | V1_27 | 20/08/2019 | 23/08/2019 | input | | 40459 | 95.74% | 31.01% |
| V1 | V1_27 | 20/08/2019 | 23/08/2019 | output | | 32569 | 89.84% | 66.77% |
| V2 | V2_27 | 20/08/2019 | 23/08/2019 | output | | 30849 | 97.33% | 75.29% |
| HC | HC_56 | 21/08/2019 | 23/08/2019 | output | | 31742 | 90.42% | 73.52% |
| V1 | V1_56 | 21/08/2019 | 23/08/2019 | output | | 30571 | 98.04% | 81.31% |

Highlighted samples are exceptions to the overall flow cytometry analysis, as explained previously. Abbreviations: HC — healthy control; V1 — patient with Clinically Isolated Syndrome/Relapsing-Remitting Multiple Sclerosis at visit 1; V2 — patient with Clinically Isolated Syndrome/Relapsing-Remitting Multiple Sclerosis at visit 2.

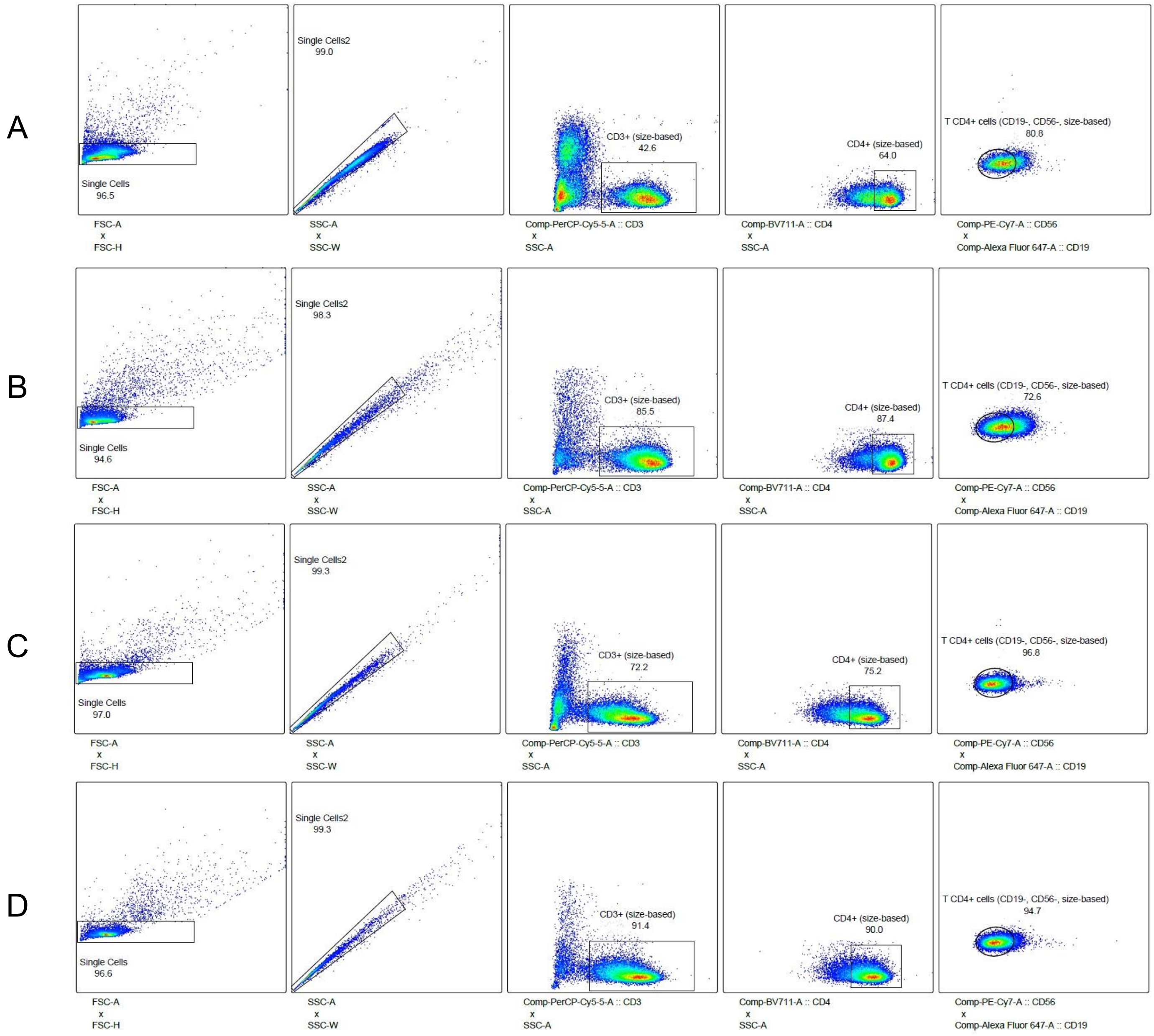

#### Figure S5 — Gating strategy in 2018 (Pilot study)

The order is: FSC-A *vs.* FSC-H, SSC-A *vs.* SSC-W, CD3-PerCP-Cy5.5 *vs.* SSC-A, CD4-BV711 *vs.* SSC-A, and CD56-PE-Cy7 *vs.* CD19-Alexa Fluor 647. The gaiting is aimed at identifying CD4+ T cells (CD3+CD4+CD56-CD19-). (**A,B**) show a previously frozen sample from a healthy control before and after undergoing CD4+ T cell enrichment, respectively; (**C,D**) show a freshly-fixed sample from a patient at VIS1 before and after undergoing CD4+ T cell enrichment, respectively. Abbreviations: VIS1 — visit 1.

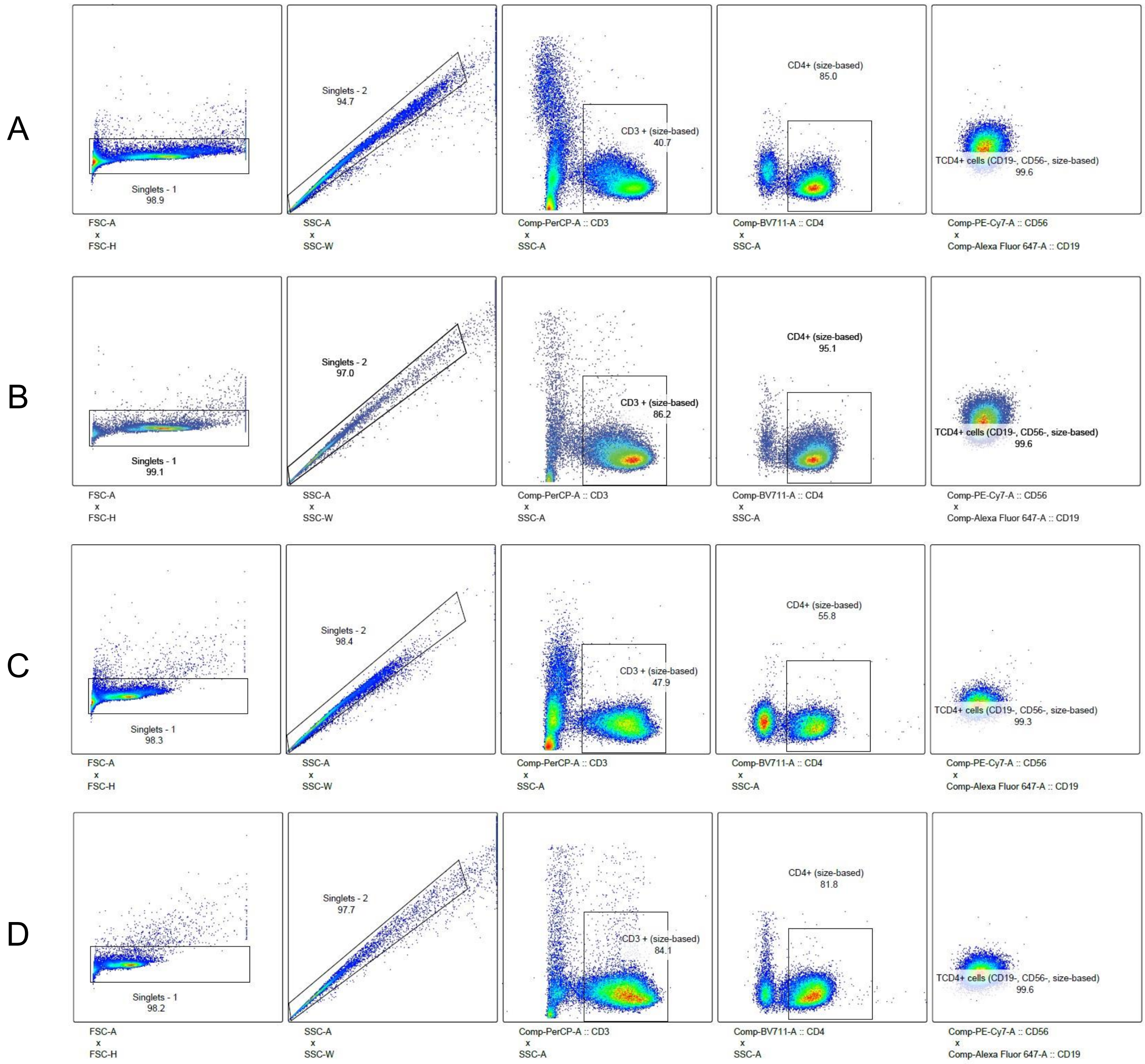

**Figure S6 — Gating strategy in 2019**

The order is: FSC-A *vs*. FSC-H, SSC-A *vs*. SSC-W, CD3-PerCP-Cy5.5 *vs.* SSC-A, CD4-BV711 *vs.* SSC-A, and CD56-PE-Cy7 *vs.* CD19-Alexa Fluor 647. The gating is aimed at identifying $CD4^+$ T cells ($CD3^+CD4^+CD56^-CD19^-$). (**A,B**) show a sample analyzed before the scheduled maintenance of the cell analyzer, on August 12, 2019, from a patient at VIS2 before and after undergoing $CD4^+$ T cell enrichment, respectively; (**C,D**) show a sample analyzed after the scheduled maintenance of the cell analyzer, from a patient at VIS1 before and after undergoing $CD4^+$ T cell enrichment, respectively. Abbreviations: VIS1 — visit 1; VIS2 — visit 2.

#### Table S4 — Flow cytometry analysis: Summary

| Subject type | Input or output? | $CD4^+$ T cells ($CD3^+CD4^+CD19^-CD56^-$) | *N* |
|---|---|---|---|
| HC | input | 23.79% | *25* |
| HC | output | 67.99% | *30* |
| V1 | input | 37.10% | *12* |
| V1 | output | 68.71% | *28* |
| V2 | input | 34.51% | *12* |
| V2 | output | 65.15% | *25* |
| **All** | **input** | **29.68%** | ***49*** |
| **All** | **output** | **67.38%** | ***83*** |

Abbreviations: HC — healthy control; *N* — number; V1 — patient with Clinically Isolated Syndrome/Relapsing-Remitting Multiple Sclerosis at visit 1; V2 — patient with Clinically Isolated Syndrome/Relapsing-Remitting Multiple Sclerosis at visit 2.

### Section 3 — mtDNA extraction and WGS

#### 3.1. DNA extraction

As mentioned in the main manuscript, DNA extraction was performed at the same time for each triplet (PwMS at VIS1&VIS2 and HC), to minimize differences within processing. In 2018, for the pilot study, samples were processed in ascending number of cells after CD4$^+$ T cell enrichment, while in 2019 samples were processed in the same order as their magnetic enrichment (Table S5). Prior to DNA extraction with the QIAamp® DNA Blood Midi Kit (QIAGEN GmbH, Hilden, Germany), samples were thawed with a Thermomixer® (Eppendorf, Hamburg, Germany) set at 37° C. After extraction, DNA concentration was measured through spectral analysis at 1 nm increments from 230 nm to 290 nm with the Synergy HTX Multi-mode Reader, in combination with its Take3 microvolume plate (BioTek Instruments, Winooski, VT, USA). The 260/280 nm absorbance ratio was used for quality control, with the aim of a 1.8 value [19]. Finally, DNA was stored at -20° C.

The enrichment of mtDNA was confirmed through a quantitative polymerase chain reaction (qPCR) on other samples not included in the cohort, which, however, underwent the same magnetic enrichment and DNA extraction protocols (Table S6, Figure S7, and Figure S8). For that, two sets of primers were used: (**I**) aimed at GAPDH [20,21] — 5'-AGGGCTGCTTTTAACTCTGGT-3' (forward) and 5'-CCCCACTTGATTTTGGAGGGA-3' (reverse) —; and (**II**) aimed at mtDNA [22,23] — 5'-CATGCAAGCATCCCCGTTCC-3' (forward) and 5'-CTGTTTCCCGTGGGGGTGTG-3' (reverse). Each sample was run in triplicate, with a no template control (NTC) for each primer set. Each non-NTC well consisted of 0.7 µL of a forward primer (10 µM), 0.7 µL of a reverse primer (10 µM), 5 µL of 10 ng of DNA resuspended in DNAse/RNase-free water, and 3.6 µL of iTaq™ Universal SYBR® Green Supermix (#1725124, Bio-Rad Laboratories, Inc., Hercules, CA, USA), in an Applied Biosystems™ MicroAmp™ Optical 384-Well Reaction Plate with Barcode (#LS4309849, Thermo Fisher Scientific, Waltham, MA, USA) with a compatible optical adhesive film. Finally, plates were centrifuged at 900 x *g* for 5 min at room temperature and analyzed on an Applied Biosystems® ViiA™ 7 Real-Time PCR System (Thermo Fisher Scientific, Waltham, MA, USA) for 40 cycles, with a denaturing temperature of 95° C, and an annealing temperature of 58° C.

#### 3.2. Bioinformatic processing and data analysis

For bioinformatic processing, we used the PrecisionCallerPipeline (PCP) method, which we had previously developed to output fully aligned BAM files mapped to the commonly used reference sequence, the revised Cambridge Reference Sequence (rCRS) [24], from the FASTQ files received from the sequencing facility [25]. Briefly, the workflow is based on Snakemake [26] and incorporates: (**I**) Awk, for SAM file editing [27]; (**II**) BEDTools, for BAM to FASTQ conversion [28]; (**III**) BWA-MEM, for read alignment [29]; (**IV**) Pycision, for amplicon delimitation and selection [30]; (**V**) SAMtools for BAM conversion, sorting, indexing, and merging

[31]; (**VI**) Trimmomatic for read quality control and trimming [32]; and (**VII**) RtN! for the removal of nuclear insertions of mitochondrial DNA (NUMTs) [33].

Samples were processed in a Linux-based system in a secure server. As done previously [25], reads were cropped at 160 base pairs (bps).

As explained in the main manuscript, variant calling was performed with mutserve v2 [34,35]. We used a base quality score of 20 and excluded variants in positions 302–315, 523–524, and 3104–3110. Only single nucleotide substitutions were considered, and variants with a variant level (VL) < 2.5% were filtered [25].

Haplogroup calling was carried out through HaploGrep v2.4.0 [36] and a contamination check was done with Haplocheck v1.3.3 [35], based on the output from mutserve.

Regarding the variants from the Ion Torrent Suite™ Software (TSS), data from each run was processed with TSS, using the reference sequence PrecisionID_mtDNA_rCRS, and target regions PrecisionID_mtDNA_WG_targets with the plugins CoverageAnalysis and VariantCaller. FASTQ and BAM files were generated using the plugin FileExporter. The software versions ranged from v5.8, v5.10, and v5.12, according to the date of each run. We compiled the VCF files arising from the sequencing runs and corrected all positions > 16,569 to the first 80 bps in the rCRS. When we found the same variants with discordant coverages and VLs, particularly in the first 80 bps, we calculated the mean coverage and the mean VL per mutation. Only single nucleotide substitutions were considered.

To account for false positives [25], variants with a VL ≥ 10% were only accepted if they were found with both PCP and TSS, while variants with a VL < 10% only present in PCP were only accepted if: (**I**) the normalized coverage, coverage ratio, mean value of reported NUMTs, and the distance to the amplicon's edge was not lower, higher, higher, or lower, respectively, than their corresponding outliers for variants found in both PCP and TSS, calculated through the Tukey's fences method [37]; and (**II**) variants were previously reported in either the HelixMTdb database [38], the ClinVar database (until October 2021) [39] through MitImpact 3D 3.0.7 [40], or in the MITOMAP database [41]. Variants only present in TSS were excluded.

Data analysis was performed with R version 4.1.1 [42] in RStudio [43] with the packages extrafont [44], infer [45], magick [46], patchwork [47], readxl [48], remotes [49], rstatix [50], scales [51], svglite [52], and tidyverse [53], as well as Excel 2016 (Microsoft Corporation, Redmond, WA, USA).

#### 3.3. Figures and Tables

#### Table S5 — DNA extraction from clinical samples after CD4$^{+}$ T cell enrichment

| Subject Type | ID | Separation date | DNA extraction date | Number of cells after enrichment (millions) | Mortality | Flow cytometry analysis? | DNA (Synergy HTX) (ng/µL) | 260/280 ratio | DNA (Qubit®) (ng/µL) |
|---|---|---|---|---|---|---|---|---|---|
| HC | HC_1 | 21/04/2018 | 06/07/2018 | 2.63 | 48.05% | No | 20.12 | 1.96 | 13.00 |
| V1 | V1_1 | 21/04/2018 | 06/07/2018 | 4.11 | 8.62% | Yes, output only | 38.40 | 1.86 | 21.00 |
| V2 | V2_1 | 21/04/2018 | 06/07/2018 | 2.38 | 10.73% | Yes, output only | 6.89 | 2.38 | 9.64 |
| HC | HC_2 | 21/04/2018 | 06/07/2018 | 4.92 | 8.71% | Yes | 38.83 | 1.92 | 26.00 |
| V1 | V1_2 | 21/04/2018 | 06/07/2018 | 3.94 | 6.50% | Yes | 23.06 | 1.93 | 14.80 |
| V2 | V2_2 | 21/04/2018 | 06/07/2018 | 2.62 | 11.18% | No | 15.83 | 2.01 | 13.00 |
| HC | HC_3 | 22/04/2018 | 19/07/2018 | 4.42 | 15.27% | Yes | 24.32 | 1.91 | 9.22 |
| V1 | V1_3 | 22/04/2018 | 19/07/2018 | 3.83 | 12.47% | No | 20.16 | 1.94 | 16.10 |
| V2 | V2_3 | 22/04/2018 | 19/07/2018 | 6.09 | 11.61% | Yes | 28.31 | 2.18 | 26.80 |
| HC | HC_4 | 22/04/2018 | 14/06/2018 | 1.27 | 11.67% | No | 5.25 | 1.86 | 5.40 |
| V1 | V1_4 | 22/04/2018 | 14/06/2018 | 1.93 | 6.78% | Yes | 7.30 | 2.02 | 4.80 |
| V2 | V2_4 | 22/04/2018 | 14/06/2018 | 5.40 | 9.76% | No | 26.78 | 1.95 | 19.70 |
| HC | HC_5 | 29/04/2018 | 11/07/2018 | 2.69 | 10.70% | Yes | 24.29 | 1.66 | 9.20 |
| V1 | V1_5 | 29/04/2018 | 11/07/2018 | 6.40 | 9.53% | Yes | 45.33 | 1.83 | 37.00 |
| V2 | V2_5 | 29/04/2018 | 11/07/2018 | 5.29 | 13.49% | Yes | 33.65 | 2.10 | 17.10 |
| HC | HC_6 | 29/04/2018 | 14/06/2018 | 4.62 | 13.58% | No | 45.79 | 2.28 | 29.80 |
| V1 | V1_6 | 29/04/2018 | 14/06/2018 | 1.80 | 12.65% | No | 6.30 | 3.05 | 6.10 |
| V2 | V2_6 | 29/04/2018 | 14/06/2018 | 2.13 | 22.36% | No | 14.70 | 2.34 | 10.60 |
| HC | HC_7 | 30/04/2018 | 29/06/2018 | 4.00 | 11.67% | No | 26.96 | 2.11 | 18.20 |
| V1 | V1_7 | 30/04/2018 | 29/06/2018 | 1.81 | 11.63% | No | 10.87 | 1.19 | 4.80 |
| V2 | V2_7 | 30/04/2018 | 29/06/2018 | 3.08 | 12.46% | No | 15.93 | 1.97 | 11.30 |
| HC | HC_8 | 30/04/2018 | 06/07/2018 | 8.27 | 15.29% | Yes | 65.24 | 1.91 | 56.00 |
| V1 | V1_8 | 30/04/2018 | 06/07/2018 | 2.30 | 16.08% | Yes, output only | 2.93 | 1.36 | 5.50 |
| V2 | V2_8 | 30/04/2018 | 06/07/2018 | 3.50 | 19.52% | Yes | 34.04 | 2.00 | 19.70 |
| HC | HC_9 | 05/05/2018 | 11/07/2018 | 2.66 | 12.56% | Yes | 17.53 | 1.55 | 16.30 |
| V1 | V1_9 | 05/05/2018 | 11/07/2018 | 3.07 | 17.78% | No | 16.22 | 2.06 | 14.10 |
| V2 | V2_9 | 05/05/2018 | 11/07/2018 | 4.01 | 15.96% | No | 25.25 | 1.94 | 22.40 |
| HC | HC_10 | 05/05/2018 | 19/07/2018 | 4.34 | 16.96% | No | 26.07 | 2.08 | 16.90 |
| V1 | V1_10 | 05/05/2018 | 19/07/2018 | 4.78 | 11.08% | Yes | 20.79 | 2.43 | 23.60 |
| V2 | V2_10 | 05/05/2018 | 19/07/2018 | 8.10 | 8.63% | Yes | 53.46 | 1.92 | 27.20 |
| HC | HC_11 | 06/05/2018 | 19/07/2018 | 2.78 | 13.57% | Yes | 9.67 | 2.19 | 8.10 |
| V1 | V1_11 | 06/05/2018 | 19/07/2018 | 5.27 | 10.21% | Yes | 27.86 | 1.84 | 19.30 |
| V2 | V2_11 | 06/05/2018 | 19/07/2018 | 7.38 | 9.60% | Yes | 26.71 | 1.99 | 20.80 |
| HC | HC_12 | 06/05/2018 | 29/06/2018 | 1.97 | 14.04% | No | 6.66 | 2.02 | 6.20 |
| V1 | V1_12 | 06/05/2018 | 29/06/2018 | 5.59 | 11.12% | No | 28.59 | 1.78 | 19.30 |
| V2 | V2_12 | 06/05/2018 | 29/06/2018 | 4.42 | 8.94% | No | 32.19 | 1.31 | 26.20 |
| HC | HC_13 | 11/05/2018 | 11/07/2018 | 5.43 | 11.70% | Yes | 39.35 | 2.04 | 27.00 |
| V1 | V1_13 | 11/05/2018 | 11/07/2018 | 2.64 | 12.98% | Yes | 12.47 | 1.96 | 9.48 |
| V2 | V2_13 | 11/05/2018 | 11/07/2018 | 3.88 | 8.35% | Yes | 17.34 | 1.91 | 13.90 |
| HC | HC_14 | 11/05/2018 | 29/06/2018 | 1.94 | 40.28% | No | 8.86 | 1.51 | 6.50 |
| V1 | V1_14 | 11/05/2018 | 29/06/2018 | 2.36 | 17.60% | No | 11.13 | 1.94 | 7.96 |
| V2 | V2_14 | 11/05/2018 | 29/06/2018 | 4.13 | 12.67% | No | 24.46 | 1.98 | 17.40 |

#### Table S5 — DNA extraction from clinical samples after CD4+ T cell enrichment (continued)

| Subject Type | ID | Separation date | DNA extraction date | Number of cells after enrichment (millions) | Mortality | Flow cytometry analysis? | DNA (Synergy HTX) (ng/µL) | 260/280 ratio | DNA (Qubit®) (ng/µL) |
|---|---|---|---|---|---|---|---|---|---|
| HC | HC_15 | 12/05/2018 | 29/06/2018 | 4.57 | 10.73% | No | 20.80 | 1.97 | 17.50 |
| V1 | V1_15 | 12/05/2018 | 29/06/2018 | 2.08 | 12.78% | No | 11.49 | 1.94 | 9.60 |
| V2 | V2_15 | 12/05/2018 | 29/06/2018 | 2.75 | 9.90% | Yes | 11.57 | 1.82 | 8.40 |
| HC | HC_16 | 12/05/2018 | 11/07/2018 | 3.77 | 29.30% | Yes | 11.70 | 2.06 | 11.50 |
| V1 | V1_16 | 12/05/2018 | 11/07/2018 | 2.59 | 14.16% | Yes | 9.56 | 1.69 | 5.90 |
| V2 | V2_16 | 12/05/2018 | 11/07/2018 | 4.54 | 21.02% | No | 24.41 | 1.98 | 17.30 |
| HC | HC_17 | 14/05/2018 | 28/05/2018 | 1.81 | 11.75% | No | 7.36 | 1.76 | 8.00 |
| V1 | V1_17 | 14/05/2018 | 28/05/2018 | 3.19 | 7.75% | Yes | 13.16 | 1.90 | 10.80 |
| V2 | V2_17 | 14/05/2018 | 28/05/2018 | 1.01 | 14.46% | No | 3.48 | 1.66 | 3.70 |
| HC | HC_18 | 14/05/2018 | 28/05/2018 | 0.70 | 60.08% | No | 1.45 | 1.19 | 2.70 |
| V1 | V1_18 | 14/05/2018 | 28/05/2018 | 2.39 | 15.39% | No | 9.87 | 1.76 | 7.80 |
| V2 | V2_18 | 14/05/2018 | 28/05/2018 | 5.54 | 13.74% | Yes | 33.12 | 2.17 | 19.90 |
| HC | HC_19 | 15/05/2018 | 19/07/2018 | 4.39 | 15.90% | No | 20.63 | 1.90 | 15.20 |
| V1 | V1_19 | 15/05/2018 | 19/07/2018 | 3.56 | 11.32% | No | 16.58 | 1.75 | 12.20 |
| V2 | V2_19 | 15/05/2018 | 19/07/2018 | 4.17 | 12.50% | Yes | 18.32 | 1.93 | 13.80 |
| HC | HC_20 | 15/05/2018 | 06/07/2018 | 4.65 | 16.55% | Yes | 20.96 | 2.16 | 19.00 |
| V1 | V1_20 | 15/05/2018 | 06/07/2018 | 3.83 | 9.08% | Yes | 19.14 | 1.80 | 13.90 |
| V2 | V2_20 | 15/05/2018 | 06/07/2018 | 2.67 | 21.30% | No | 11.55 | 2.21 | 10.30 |
| HC | HC_21 | 24/05/2019 | 05/06/2019 | 2.20 | 14.65% | Yes | 15.18 | 2.39 | 9.62 |
| V1 | V1_21 | 24/05/2019 | 05/06/2019 | 1.75 | 30.09% | Yes | 6.60 | 2.78 | 5.34 |
| V2 | V2_21 | 24/05/2019 | 05/06/2019 | 1.22 | 28.13% | Yes, input only | 11.12 | 1.99 | 6.10 |
| HC | HC_57 | 24/05/2019 | 05/06/2019 | 1.70 | 14.49% | Yes | 12.66 | 1.99 | 6.40 |
| V1 | V1_57 | 24/05/2019 | 05/06/2019 | 1.68 | 22.56% | No | 13.77 | 3.37 | 6.90 |
| V2 | V2_57 | 24/05/2019 | 05/06/2019 | 1.57 | 18.99% | No | 10.08 | 2.31 | 5.38 |
| HC | HC_38 | 29/05/2019 | 05/06/2019 | 2.12 | 5.36% | Yes | 12.55 | 1.97 | 7.80 |
| V1 | V1_38 | 29/05/2019 | 05/06/2019 | 0.86 | 9.27% | No | 11.98 | 1.67 | 4.44 |
| V2 | V2_38 | 29/05/2019 | 05/06/2019 | 1.07 | 7.99% | No | 10.22 | 2.07 | 2.12 |
| HC | HC_30 | 29/05/2019 | 05/06/2019 | 1.70 | 12.42% | No | 10.50 | 2.07 | 6.78 |
| V1 | V1_30 | 29/05/2019 | 05/06/2019 | 1.77 | 23.77% | No | 8.81 | 1.69 | 5.00 |
| V2 | V2_30 | 29/05/2019 | 05/06/2019 | 0.10 | 56.25% | No | 4.26 | 2.24 | 0.70 |
| HC | HC_37 | 30/05/2019 | 05/06/2019 | 1.54 | 18.75% | No | 7.37 | 1.58 | 5.20 |
| V1 | V1_37 | 30/05/2019 | 05/06/2019 | 1.75 | 19.29% | No | 16.70 | 1.81 | 8.36 |
| V2 | V2_37 | 30/05/2019 | 05/06/2019 | 2.37 | 9.54% | No | 16.86 | 1.90 | 7.58 |
| HC | HC_45 | 30/05/2019 | 05/06/2019 | 1.51 | 22.38% | No | 11.88 | 2.01 | 7.54 |
| V1 | V1_45 | 30/05/2019 | 05/06/2019 | 2.26 | 5.57% | Yes, output only | 17.20 | 1.83 | 12.20 |
| V2 | V2_45 | 30/05/2019 | 05/06/2019 | 3.26 | 14.26% | Yes | 25.09 | 2.00 | 16.70 |
| HC | HC_50 | 31/05/2019 | 05/06/2019 | 0.37 | 70.86% | No | 6.36 | 1.82 | 1.91 |
| V1 | V1_50 | 31/05/2019 | 05/06/2019 | 2.29 | 15.04% | Yes, output only | 17.54 | 2.03 | 11.00 |
| V2 | V2_50 | 31/05/2019 | 05/06/2019 | 1.25 | 12.10% | No | 7.32 | 1.89 | 4.22 |
| HC | HC_35 | 31/05/2019 | 05/06/2019 | 1.78 | 14.05% | Yes, output only | 11.54 | 1.97 | 6.52 |
| V1 | V1_35 | 31/05/2019 | 05/06/2019 | 1.33 | 13.33% | No | 7.88 | 1.86 | 4.70 |
| V2 | V2_35 | 31/05/2019 | 05/06/2019 | 2.17 | 33.86% | Yes, output only | 12.18 | 2.09 | 5.94 |
| HC | HC_46 | 01/06/2019 | 05/06/2019 | 2.71 | 8.97% | No | 13.52 | 1.83 | 9.60 |

#### Table S5 — DNA extraction from clinical samples after CD4+ T cell enrichment (continued)

| Subject Type | ID | Separation date | DNA extraction date | Number of cells after enrichment (millions) | Mortality | Flow cytometry analysis? | DNA (Synergy HTX) (ng/µL) | 260/280 ratio | DNA (Qubit®) (ng/µL) |
|---|---|---|---|---|---|---|---|---|---|
| V1 | V1_46 | 01/06/2019 | 05/06/2019 | 1.73 | 23.08% | No | 10.71 | 1.86 | 6.28 |
| V2 | V2_46 | 01/06/2019 | 05/06/2019 | 0.59 | 32.71% | No | 8.30 | 1.91 | 4.72 |
| HC | HC_44 | 01/06/2019 | 05/06/2019 | 2.20 | 10.72% | Yes | 18.04 | 2.01 | 10.90 |
| V1 | V1_44 | 01/06/2019 | 05/06/2019 | 1.65 | 14.21% | Yes, output only | 11.38 | 1.84 | 8.34 |
| V2 | V2_44 | 01/06/2019 | 05/06/2019 | 1.85 | 22.78% | Yes, output only | 13.67 | 2.21 | 9.90 |
| HC | HC_47 | 16/06/2019 | 14/07/2019 | 2.55 | 8.50% | No | 23.12 | 1.95 | 16.10 |
| V1 | V1_47 | 16/06/2019 | 14/07/2019 | 2.04 | 23.17% | No | 15.60 | 2.63 | 7.80 |
| V2 | V2_47 | 16/06/2019 | 14/07/2019 | 2.04 | 22.69% | Yes, output only | 11.35 | 1.94 | 6.90 |
| HC | HC_55 | 16/06/2019 | 14/07/2019 | 2.23 | 4.34% | Yes | 17.82 | 1.98 | 10.90 |
| V1 | V1_55 | 16/06/2019 | 14/07/2019 | 1.75 | 18.90% | Yes, output only | 12.15 | 1.99 | 7.90 |
| V2 | V2_55 | 16/06/2019 | 14/07/2019 | 1.36 | 20.56% | No | 9.31 | 1.79 | 8.10 |
| HC | HC_22 | 19/06/2019 | 14/07/2019 | 3.64 | 7.64% | Yes, output only | 17.40 | 1.98 | 14.80 |
| V1 | V1_22 | 19/06/2019 | 14/07/2019 | 1.75 | 14.50% | Yes, output only | 15.86 | 1.80 | 6.60 |
| V2 | V2_22 | 19/06/2019 | 14/07/2019 | 1.01 | 28.21% | No | 6.58 | 2.34 | 4.50 |
| HC | HC_58 | 19/06/2019 | 14/07/2019 | 2.75 | 5.00% | Yes | 20.14 | 1.95 | 13.00 |
| V1 | V1_58 | 19/06/2019 | 14/07/2019 | 0.57 | 17.66% | No | 26.26 | 1.89 | 16.90 |
| V2 | V2_58 | 19/06/2019 | 14/07/2019 | 2.09 | 25.35% | Yes, output only | 13.42 | 1.90 | 7.96 |
| HC | HC_61 | 20/06/2019 | 14/07/2019 | 2.90 | 8.05% | Yes | 13.83 | 2.62 | 7.84 |
| V1 | V1_61 | 20/06/2019 | 14/07/2019 | 1.69 | 17.91% | Yes, output only | 14.03 | 2.23 | 7.82 |
| V2 | V2_61 | 20/06/2019 | 14/07/2019 | 1.92 | 8.40% | No | 14.34 | 2.03 | 10.40 |
| HC | HC_24 | 20/06/2019 | 14/07/2019 | 8.92 | 4.42% | Yes, output only | 53.27 | 2.11 | 35.40 |
| V1 | V1_24 | 20/06/2019 | 14/07/2019 | 2.06 | 33.41% | No | 9.72 | 1.97 | 6.40 |
| V2 | V2_24 | 20/06/2019 | 14/07/2019 | 1.78 | 18.54% | Yes, output only | 10.10 | 1.74 | 6.20 |
| HC | HC_59 | 26/06/2019 | 14/07/2019 | 4.30 | 7.99% | Yes | 24.79 | 1.93 | 17.70 |
| V1 | V1_59 | 26/06/2019 | 14/07/2019 | 2.17 | 12.85% | No | 11.23 | 1.89 | 8.30 |
| V2 | V2_59 | 26/06/2019 | 14/07/2019 | 3.18 | 9.92% | Yes | 16.52 | 1.91 | 12.30 |
| HC | HC_39 | 26/06/2019 | 14/07/2019 | 3.26 | 5.12% | No | 19.04 | 1.94 | 13.90 |
| V1 | V1_39 | 26/06/2019 | 14/07/2019 | 1.88 | 18.36% | Yes, output only | 9.09 | 1.94 | 6.10 |
| V2 | V2_39 | 26/06/2019 | 14/07/2019 | 3.16 | 11.40% | No | 25.08 | 2.33 | 13.10 |
| HC | HC_42 | 27/06/2019 | 14/07/2019 | 4.56 | 7.16% | No | 35.64 | 1.74 | 24.80 |
| V1 | V1_42 | 27/06/2019 | 14/07/2019 | 2.31 | 9.20% | Yes, output only | 20.34 | 1.87 | 10.60 |
| V2 | V2_42 | 27/06/2019 | 14/07/2019 | 2.71 | 14.03% | Yes | 17.69 | 1.96 | 12.00 |
| HC | HC_23 | 27/06/2019 | 14/07/2019 | 2.85 | 8.70% | Yes | 24.91 | 1.96 | 17.10 |
| V1 | V1_23 | 27/06/2019 | 14/07/2019 | 1.73 | 12.42% | No | 13.64 | 2.08 | 8.24 |
| V2 | V2_23 | 27/06/2019 | 14/07/2019 | 2.30 | 11.66% | No | 24.65 | 1.91 | 16.50 |
| HC | HC_40 | 02/07/2019 | 23/07/2019 | 3.39 | 26.22% | Yes | 24.42 | 1.76 | 18.00 |
| V1 | V1_40 | 02/07/2019 | 23/07/2019 | 3.01 | 5.96% | Yes, output only | 20.75 | 1.75 | 15.50 |
| V2 | V2_40 | 02/07/2019 | 23/07/2019 | 1.95 | 18.27% | No | 9.57 | 1.71 | 7.42 |
| HC | HC_51 | 03/07/2019 | 23/07/2019 | 3.94 | 11.57% | Yes, output only | 23.99 | 1.93 | 19.30 |
| V1 | V1_51 | 03/07/2019 | 23/07/2019 | 1.65 | 20.00% | Yes, output only | 11.74 | 1.91 | 7.06 |
| V2 | V2_51 | 03/07/2019 | 23/07/2019 | 2.12 | 14.73% | Yes, output only | 10.68 | 1.80 | 9.08 |
| HC | HC_52 | 03/07/2019 | 23/07/2019 | 1.68 | 10.18% | Yes | 6.67 | 1.98 | 5.32 |
| V1 | V1_52 | 03/07/2019 | 23/07/2019 | 4.03 | 9.62% | Yes | 26.70 | 1.91 | 19.00 |

#### Table S5 — DNA extraction from clinical samples after CD4+ T cell enrichment (continued)

| Subject Type | ID | Separation date | DNA extraction date | Number of cells after enrichment (millions) | Mortality | Flow cytometry analysis? | DNA (Synergy HTX) (ng/µL) | 260/280 ratio | DNA (Qubit®) (ng/µL) |
|---|---|---|---|---|---|---|---|---|---|
| V2 | V2_52 | 03/07/2019 | 23/07/2019 | 1.95 | 7.43% | Yes, output only | 10.71 | 2.15 | 5.60 |
| HC | HC_49 | 03/07/2019 | 23/07/2019 | 2.30 | 13.51% | No | 14.37 | 1.61 | 7.66 |
| V1 | V1_49 | 03/07/2019 | 23/07/2019 | 2.24 | 8.97% | No | 15.27 | 1.93 | 9.58 |
| V2 | V2_49 | 03/07/2019 | 23/07/2019 | 1.68 | 16.63% | No | 10.62 | 1.85 | 7.66 |
| HC | HC_34 | 17/07/2019 | 23/07/2019 | 1.77 | 23.99% | No | 11.59 | 1.73 | 8.64 |
| V1 | V1_34 | 17/07/2019 | 23/07/2019 | 1.44 | 9.69% | No | 11.03 | 1.77 | 8.80 |
| V2 | V2_34 | 17/07/2019 | 23/07/2019 | 1.12 | 16.38% | No | 10.71 | 1.80 | 5.22 |
| HC | HC_29 | 17/07/2019 | 24/07/2019 | 3.09 | 12.20% | Yes | 20.51 | 1.89 | 13.10 |
| V1 | V1_29 | 17/07/2019 | 24/07/2019 | 1.75 | 30.36% | No | 16.37 | 1.94 | 12.00 |
| V2 | V2_29 | 17/07/2019 | 24/07/2019 | 1.60 | 13.95% | Yes, output only | 7.57 | 2.32 | 6.12 |
| HC | HC_41 | 18/07/2019 | 24/07/2019 | 2.57 | 8.68% | No | 19.95 | 1.81 | 14.60 |
| V1 | V1_41 | 18/07/2019 | 24/07/2019 | 2.20 | 14.68% | No | 15.91 | 1.80 | 10.40 |
| V2 | V2_41 | 18/07/2019 | 24/07/2019 | 3.24 | 19.28% | Yes, output only | 15.86 | 1.81 | 10.60 |
| HC | HC_28 | 18/07/2019 | 24/07/2019 | 2.52 | 8.97% | Yes | 12.55 | 1.84 | 10.40 |
| V1 | V1_28 | 18/07/2019 | 24/07/2019 | 1.92 | 9.93% | Yes, output only | 16.30 | 1.81 | 13.80 |
| V2 | V2_28 | 18/07/2019 | 24/07/2019 | 1.31 | 21.77% | No | 9.95 | 1.46 | 5.10 |
| HC | HC_48 | 20/07/2019 | 24/07/2019 | 3.86 | 5.20% | No | 25.23 | 1.83 | 16.20 |
| V1 | V1_48 | 20/07/2019 | 24/07/2019 | 2.09 | 15.38% | No | 10.62 | 1.77 | 7.70 |
| V2 | V2_48 | 20/07/2019 | 24/07/2019 | 1.20 | 22.99% | No | 13.44 | 1.77 | 8.32 |
| HC | HC_53 | 20/07/2019 | 24/07/2019 | 3.77 | 20.66% | Yes | 25.47 | 1.84 | 17.40 |
| V1 | V1_53 | 20/07/2019 | 24/07/2019 | 2.50 | 15.13% | Yes, output only | 13.93 | 1.99 | 10.10 |
| V2 | V2_53 | 20/07/2019 | 24/07/2019 | 1.20 | 11.93% | No | 4.91 | 5.20 | 4.24 |
| HC | HC_25 | 21/07/2019 | 26/08/2019 | 2.12 | 7.86% | No | 18.61 | 1.65 | 12.60 |
| V1 | V1_25 | 21/07/2019 | 26/08/2019 | 1.77 | 11.39% | No | 14.99 | 1.69 | 7.92 |
| V2 | V2_25 | 21/07/2019 | 26/08/2019 | 0.33 | 18.75% | No | 11.50 | 1.56 | 7.18 |
| HC | HC_60 | 21/07/2019 | 26/08/2019 | 3.10 | 6.31% | Yes | 27.14 | 1.71 | 18.80 |
| V1 | V1_60 | 21/07/2019 | 26/08/2019 | 3.50 | 12.42% | Yes, output only | 24.05 | 1.77 | 16.20 |
| V2 | V2_60 | 21/07/2019 | 26/08/2019 | 1.67 | 19.09% | Yes, output only | 13.11 | 1.47 | 8.90 |
| HC | HC_31 | 15/08/2019 | 26/08/2019 | 5.72 | 7.71% | Yes | 41.07 | 1.82 | 28.20 |
| V1 | V1_31 | 15/08/2019 | 26/08/2019 | 1.84 | 8.05% | No | 17.67 | 1.47 | 10.20 |
| V2 | V2_31 | 15/08/2019 | 26/08/2019 | 1.64 | 22.81% | Yes, output only | 7.01 | 1.28 | 3.20 |
| HC | HC_43 | 15/08/2019 | 26/08/2019 | 2.63 | 7.92% | No | 18.82 | 1.65 | 12.10 |
| V1 | V1_43 | 15/08/2019 | 26/08/2019 | 1.69 | 23.02% | No | 12.40 | 1.52 | 7.04 |
| V2 | V2_43 | 15/08/2019 | 26/08/2019 | 0.64 | 43.65% | No | 11.74 | 1.15 | 7.84 |
| HC | HC_26 | 16/08/2019 | 26/08/2019 | 2.78 | 7.65% | Yes | 23.59 | 1.62 | 15.60 |
| V1 | V1_26 | 16/08/2019 | 26/08/2019 | 1.58 | 10.01% | Yes, output only | 14.90 | 1.50 | 8.68 |
| V2 | V2_26 | 16/08/2019 | 26/08/2019 | 1.22 | 26.24% | No | 9.13 | 1.51 | 5.34 |
| HC | HC_36 | 16/08/2019 | 26/08/2019 | 1.49 | 16.99% | No | 11.14 | 1.50 | 6.36 |
| V1 | V1_36 | 16/08/2019 | 26/08/2019 | 0.82 | 26.92% | No | 11.77 | 1.75 | 2.54 |
| V2 | V2_36 | 16/08/2019 | 26/08/2019 | 0.53 | 15.79% | No | 25.70 | 1.66 | 16.40 |
| HC | HC_54 | 17/08/2019 | 26/08/2019 | 2.49 | 19.20% | Yes | 15.78 | 1.58 | 8.00 |
| V1 | V1_54 | 17/08/2019 | 26/08/2019 | 0.93 | 27.33% | No | 8.08 | 1.36 | 4.26 |
| V2 | V2_54 | 17/08/2019 | 26/08/2019 | 0.74 | 27.21% | No | 8.13 | 1.30 | 3.12 |

#### Table S5 — DNA extraction from clinical samples after CD4$^+$ T cell enrichment (continued)

| Subject Type | ID | Separation date | DNA extraction date | Number of cells after enrichment (millions) | Mortality | Flow cytometry analysis? | DNA (Synergy HTX) (ng/µL) | 260/280 ratio | DNA (Qubit®) (ng/µL) |
|---|---|---|---|---|---|---|---|---|---|
| HC | HC_32 | 17/08/2019 | 26/08/2019 | 3.19 | 11.45% | No | 24.14 | 1.72 | 17.20 |
| V1 | V1_32 | 17/08/2019 | 26/08/2019 | 3.08 | 20.46% | Yes | 20.36 | 1.61 | 15.20 |
| V2 | V2_32 | 17/08/2019 | 26/08/2019 | 0.75 | 18.45% | No | 9.64 | 1.39 | 5.58 |
| HC | HC_33 | 20/08/2019 | 26/08/2019 | 1.16 | 17.73% | No | 11.71 | 1.59 | 5.84 |
| V1 | V1_33 | 20/08/2019 | 26/08/2019 | 0.89 | 16.36% | No | 11.49 | 1.61 | 4.78 |
| V2 | V2_33 | 20/08/2019 | 26/08/2019 | 1.58 | 19.39% | No | 13.98 | 1.52 | 7.48 |
| HC | HC_27 | 20/08/2019 | 26/08/2019 | 1.68 | 24.50% | No | 18.20 | 1.54 | 10.60 |
| V1 | V1_27 | 20/08/2019 | 26/08/2019 | 2.29 | 14.13% | Yes | 23.33 | 1.76 | 16.00 |
| V2 | V2_27 | 20/08/2019 | 26/08/2019 | 3.02 | 14.01% | Yes, output only | 21.56 | 1.68 | 13.00 |
| HC | HC_56 | 21/08/2019 | 26/08/2019 | 2.14 | 7.77% | Yes, output only | 15.95 | 1.67 | 9.00 |
| V1 | V1_56 | 21/08/2019 | 26/08/2019 | 1.71 | 13.11% | Yes, output only | 11.44 | 1.54 | 6.14 |
| V2 | V2_56 | 21/08/2019 | 26/08/2019 | 1.60 | 17.46% | No | 10.73 | 1.39 | 5.10 |

Abbreviations: HC — healthy control; V1 — patient with Clinically Isolated Syndrome/Relapsing-Remitting Multiple Sclerosis at visit 1; V2 — patient with Clinically Isolated Syndrome/Relapsing-Remitting Multiple Sclerosis at visit 2.

#### Table S6 — qPCR on a set of trial samples: mtDNA *vs.* nuclear DNA

| Target | Sample | Ct SD (internal) | Mean Ct |
|---|---|---|---|
| mtDNA | NTC | 0.912 | 32.604 |
| mtDNA | FCF 1 — P1 CD4+ | 0.108 | 14.696 |
| mtDNA | FCF 2 — P1 CD4- | 0.115 | 14.881 |
| mtDNA | FCF 3 — P2 CD4- | 0.260 | 15.045 |
| GAPDH | NTC | | 36.986 |
| GAPDH | FCF 1 — P1 CD4+ | 0.105 | 17.027 |
| GAPDH | FCF 2 — P1 CD4- | 0.184 | 17.152 |
| GAPDH | FCF 3 — P2 CD4- | 0.445 | 18.171 |

Abbreviations: Ct — threshold cycle; mtDNA — mitochondrial DNA; NTC — no template control; qPCR — quantitative polymerase chain reaction; SD — standard deviation.

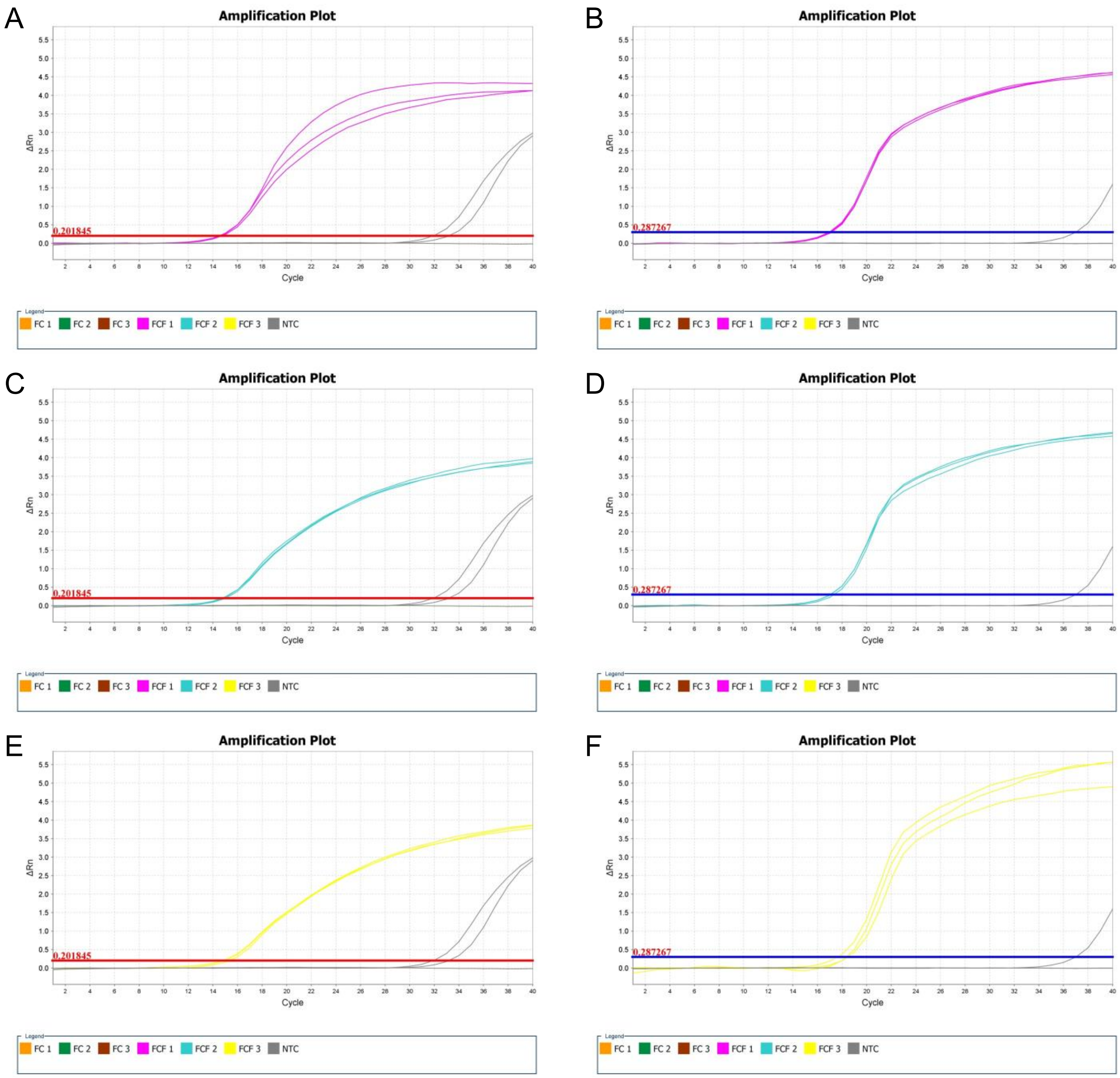

#### Figure S7 — Amplification plots of a qPCR from DNA extracted from trial samples

(**A,B**) Sample FCF 1 — P1 CD4+'s amplification plots of the mtDNA wells and the GAPDH wells, respectively; (**C,D**) Sample FCF 2 — P1 CD4-'s amplification plots of the mtDNA wells and the GAPDH wells, respectively; (**E,F**) Sample FCF 3 — P2 CD4-'s amplification plots of the mtDNA wells and the GAPDH wells, respectively. Grey depicts NTC wells from each set of primers. Abbreviations: mtDNA — mitochondrial DNA; NTC — no template control; qPCR — quantitative polymerase chain reaction.

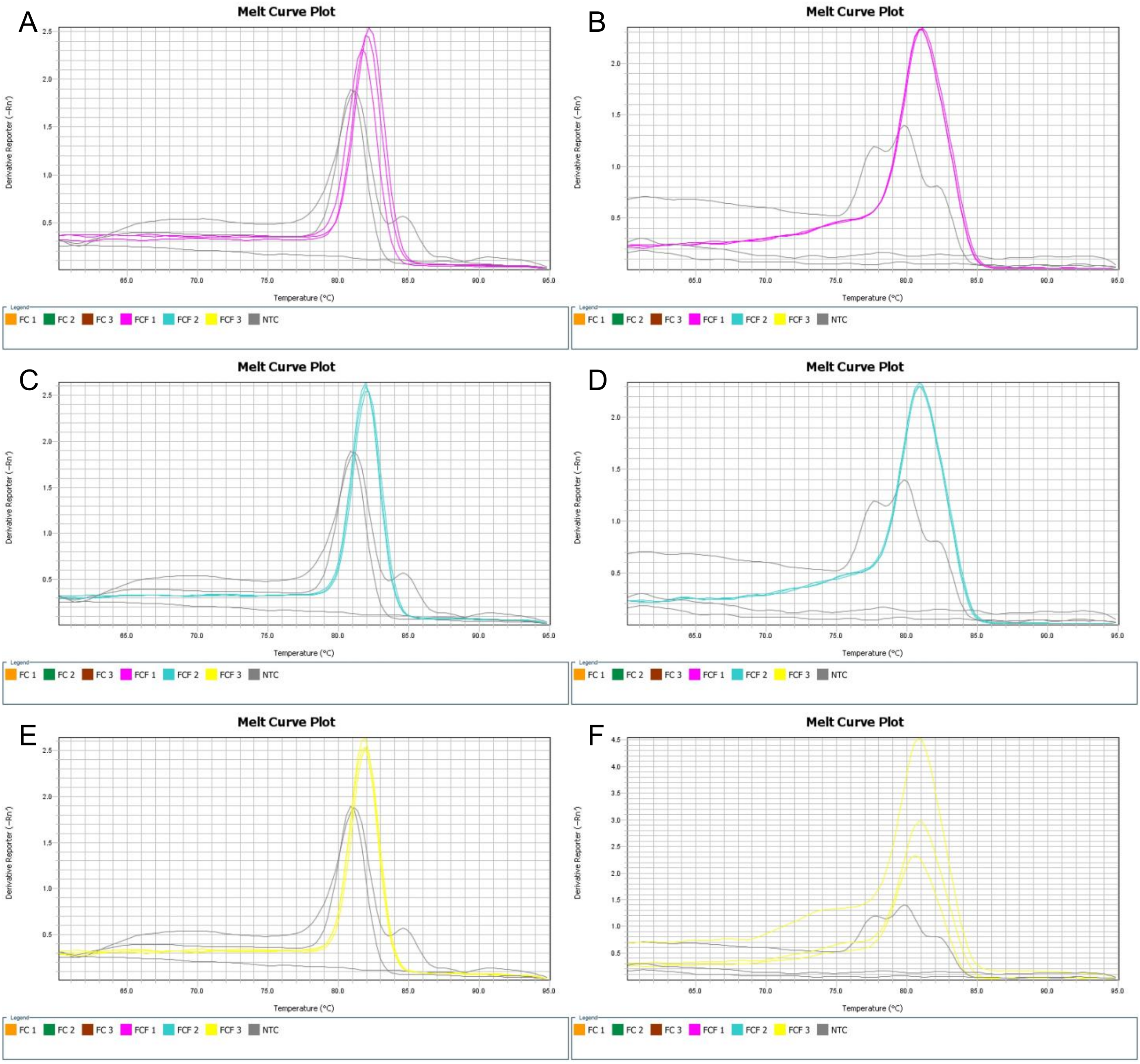

**Figure S8 — Melt curve plots of a qPCR from DNA extracted from trial samples**

(**A,B**) Sample FCF 1 — P1 CD4+'s melt curve plots of the mtDNA wells and the GAPDH wells, respectively; (**C,D**) Sample FCF 2 — P1 CD4-'s melt curve plots of the mtDNA wells and the GAPDH wells, respectively; (**E,F**) Sample FCF 3 — P2 CD4-'s melt curve plots of the mtDNA wells and the GAPDH wells, respectively. Grey depicts NTC wells from each set of primers. Abbreviations: mtDNA — mitochondrial DNA; NTC — no template control; qPCR — quantitative polymerase chain reaction.

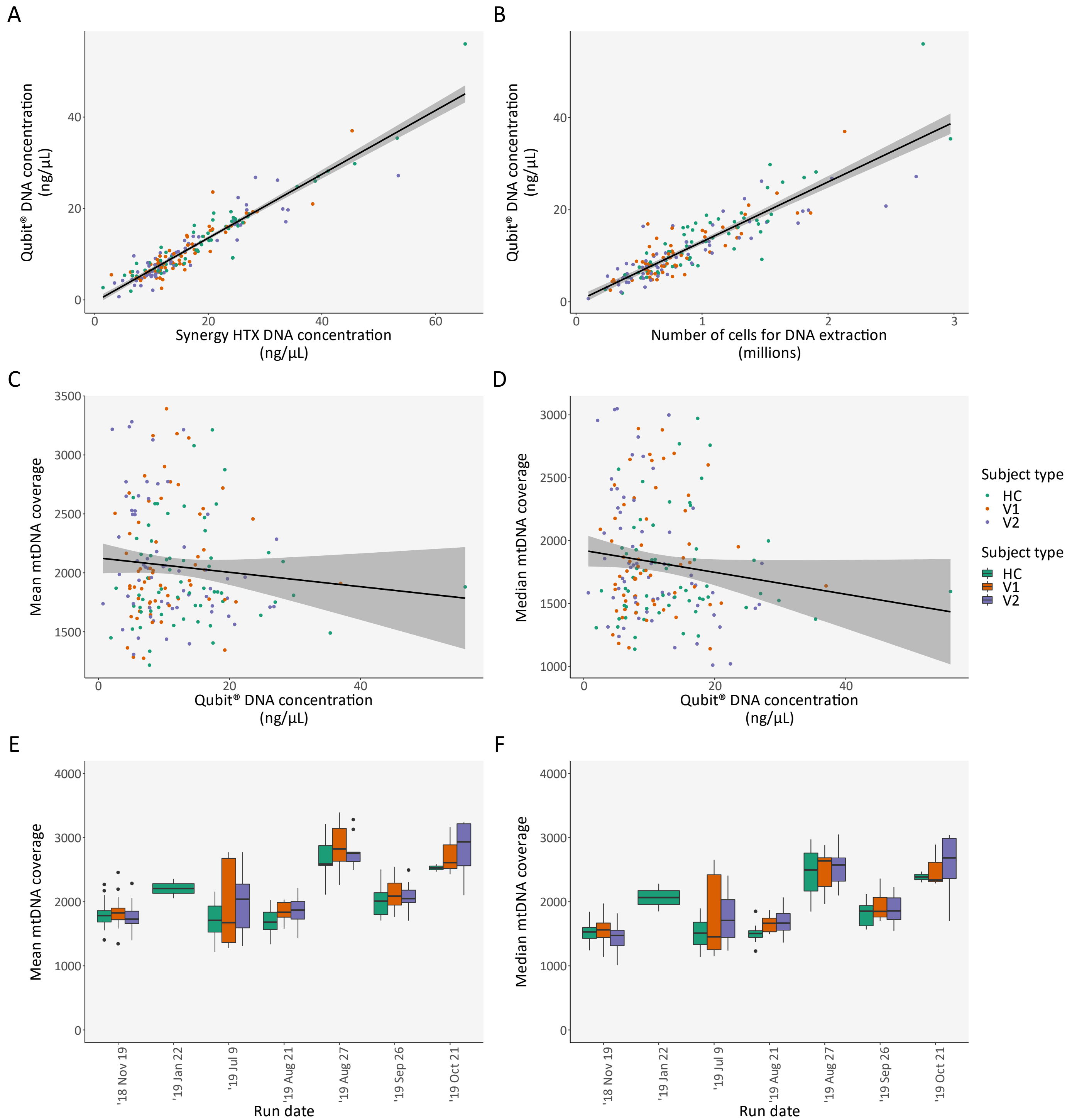

#### Figure S9 — WGS coverage analysis

(**A**) Correlation between two DNA measurements: one with the Synergy HTX Multi-mode Reader (BioTek Instruments, Winooski, VT, USA), and the other with a Qubit® 3.0 fluorometer (Thermo Fisher Scientific, Waltham, MA, USA); (**B**) Correlation between the number of cells used for DNA extraction and DNA concentration with a Qubit® 3.0 fluorometer; (**C,D**) Correlation between DNA concentration with a Qubit® 3.0 fluorometer and mean and median mtDNA coverage, respectively; (**E,F**) Mean and median mtDNA coverage, per subject type and per run date, respectively. Abbreviations: HC — healthy control; mtDNA — mitochondrial DNA; WGS — whole genome sequencing; V1 — patient with Clinically Isolated Syndrome/Relapsing-Remitting Multiple Sclerosis at visit 1; V2 — patient with Clinically Isolated Syndrome/Relapsing-Remitting Multiple Sclerosis at visit 2.

#### Table S7 — WGS in the CIS/RRMS cohort: Overall look

| ID | Mappability >30 reads/base | Mappability >100 reads/base | Mean coverage | Haplogroup | Haplogroup (simplified) | Haplogroup quality (%) | Contamination? | Haplogroup origin | Number of variants |
|---|---|---|---|---|---|---|---|---|---|
| HC_1 | 99.95% | 98.96% | 2103.88 | U4a1a | U | 97.29% | No | Europe | 33 |
| V1_1 | 99.94% | 98.78% | 1753.64 | H6a1a4 | H | 97.56% | No | Europe | 18 |
| V2_1 | 99.94% | 98.48% | 1757.07 | H6a1a4 | H | 97.56% | No | Europe | 17 |
| HC_2 | 99.94% | 99.20% | 2171.30 | H5a2 | H | 100.00% | No | Europe | 14 |
| V1_2 | 99.97% | 99.39% | 2068.36 | I1a1 | I | 93.90% | No | Europe | 42 |
| V2_2 | 99.96% | 98.29% | 1848.08 | I1a1 | I | 90.83% | No | Europe | 51 |
| HC_3 | 99.94% | 97.91% | 1833.79 | H6a1a | H | 94.18% | - | Europe | 16 |
| V1_3 | 99.92% | 98.59% | 2195.22 | J1c2c2 | J | 95.46% | No | Europe | 39 |
| V2_3 | 99.91% | 96.56% | 1713.43 | J1c2c2 | J | 95.46% | No | Europe | 38 |
| HC_4 | 99.92% | 98.96% | 1718.53 | H | H | 75.99% | No | Europe | 21 |
| V1_4 | 99.95% | 98.70% | 1888.68 | T2b1 | T | 99.61% | No | Europe | 35 |
| V2_4 | 99.90% | 97.43% | 1949.34 | T2b1 | T | 99.61% | No | Europe | 34 |
| HC_5 | 99.92% | 96.96% | 1706.79 | H1b2 | H | 92.33% | No | Europe | 14 |
| V1_5 | 99.65% | 97.54% | 1910.67 | H1a | H | 92.20% | No | Europe | 14 |
| V2_5 | 99.66% | 98.71% | 1865.97 | H1a1 | H | 94.96% | No | Europe | 15 |
| HC_6 | 98.90% | 96.84% | 1809.44 | U5a2b1a | U | 92.35% | No | Europe | 29 |
| V1_6 | 99.94% | 97.88% | 1782.18 | H1au | H | 89.76% | No | Europe | 13 |
| V2_6 | 99.92% | 96.64% | 1739.10 | H1au | H | 93.56% | No | Europe | 16 |
| HC_7 | 99.92% | 96.60% | 1837.93 | H7a1 | H | 92.82% | No | Europe | 13 |
| V1_7 | 99.89% | 97.02% | 1867.96 | J1c4 | J | 97.79% | No | Europe | 31 |
| V2_7 | 99.93% | 97.21% | 1697.30 | J1c4 | J | 97.79% | No | Europe | 30 |
| HC_8 | 99.91% | 98.35% | 1880.49 | H5a1j | H | 100.00% | No | Europe | 13 |
| V1_8 | 99.94% | 97.29% | 1879.02 | H2a2a1 | H | 50.00% | No | Europe | 4 |
| V2_8 | 99.93% | 97.07% | 1785.10 | H2a2a1 | H | 50.00% | No | Europe | 4 |
| HC_9 | 99.97% | 99.93% | 2357.52 | H1ap | H | 92.83% | No | Europe | 13 |
| V1_9 | 99.92% | 99.00% | 1897.20 | J1c3 | J | 96.67% | No | Europe | 31 |
| V2_9 | 99.70% | 97.06% | 1963.22 | J1c3 | J | 95.34% | No | Europe | 30 |
| HC_10 | 99.98% | 99.81% | 2055.22 | H1au | H | 89.76% | No | Europe | 17 |
| V1_10 | 99.92% | 98.56% | 2456.93 | T2f1a1 | T | 91.19% | No | Europe | 39 |
| V2_10 | 99.95% | 98.15% | 2285.53 | T2f1a1 | T | 91.19% | No | Europe | 39 |
| HC_11 | 99.94% | 98.80% | 2270.24 | H13a2b2a | H | 96.74% | No | Europe | 16 |
| V1_11 | 99.93% | 98.68% | 1772.31 | H15a1 | H | 98.69% | No | Europe | 14 |
| V2_11 | 99.95% | 97.77% | 1562.48 | H15a1 | H | 98.69% | No | Europe | 14 |
| HC_12 | 99.93% | 98.66% | 1810.32 | H5c1 | H | 94.58% | No | Europe | 14 |
| V1_12 | 99.93% | 97.54% | 1344.66 | V*1 | V | 93.25% | No | Europe | 18 |
| V2_12 | 99.93% | 97.49% | 1709.44 | V*1 | V | 93.25% | No | Europe | 18 |
| HC_13 | 99.92% | 97.50% | 1751.79 | W1e1a | W | 97.13% | No | Europe | 40 |
| V1_13 | 99.92% | 99.12% | 1894.20 | H1z1 | H | 100.00% | - | Europe | 13 |
| V2_13 | 99.90% | 97.05% | 1397.82 | H1z1 | H | 100.00% | No | Europe | 14 |
| HC_14 | 99.82% | 98.77% | 1668.46 | K1a+150 | K | 94.66% | No | Europe | 35 |
| V1_14 | 99.46% | 98.69% | 1643.22 | H1+152 | H | 81.34% | No | Europe | 19 |
| V2_14 | 99.89% | 99.02% | 1719.68 | H1+152 | H | 81.34% | No | Europe | 19 |

#### Table S7 — WGS in the CIS/RRMS cohort: Overall look (continued)

| ID | Mappability >30 reads/base | Mappability >100 reads/base | Mean coverage | Haplogroup | Haplogroup (simplified) | Haplogroup quality (%) | Contamination? | Haplogroup origin | Number of variants |
|---|---|---|---|---|---|---|---|---|---|
| HC_15 | 99.92% | 96.48% | 1404.58 | H1b | H | 93.28% | No | Europe | 13 |
| V1_15 | 99.92% | 97.04% | 1582.60 | J1c2t | J | 100.00% | No | Europe | 30 |
| V2_15 | 99.90% | 98.18% | 1591.83 | J1c2t | J | 100.00% | No | Europe | 30 |
| HC_16 | 99.89% | 97.47% | 1553.89 | V11 | V | 97.25% | No | Europe | 17 |
| V1_16 | 99.92% | 98.18% | 1613.20 | K1b2a1 | K | 98.43% | No | Europe | 36 |
| V2_16 | 97.46% | 92.96% | 1669.34 | K1b2a1 | K | 98.43% | No | Europe | 35 |
| HC_17 | 99.92% | 96.95% | 1675.36 | U5b2b1a1 | U | 95.66% | No | Europe | 35 |
| V1_17 | 99.90% | 96.93% | 1722.89 | U5b2a1a1 | U | 96.05% | No | Europe | 29 |
| V2_17 | 99.91% | 96.55% | 1517.45 | U5b2a1a1 | U | 95.51% | No | Europe | 29 |
| HC_18 | 99.94% | 98.25% | 1869.41 | X2b+226 | X | 96.12% | No | Europe | 28 |
| V1_18 | 99.87% | 97.49% | 1706.42 | J1b1a1b | J | 95.45% | No | Europe | 39 |
| V2_18 | 99.60% | 97.09% | 1631.77 | J1b1a1b | J | 96.69% | No | Europe | 39 |
| HC_19 | 99.16% | 96.80% | 1584.42 | K1a1a | K | 97.99% | No | Europe | 33 |
| V1_19 | 99.93% | 99.11% | 1780.61 | K1c1f | K | 95.47% | No | Europe | 34 |
| V2_19 | 99.89% | 98.56% | 1813.83 | K1c1f | K | 97.10% | No | Europe | 34 |
| HC_20 | 99.97% | 98.27% | 1757.74 | I1a1c | I | 97.35% | No | Europe | 39 |
| V1_20 | 99.95% | 98.73% | 1933.71 | H16b | H | 100.00% | No | Europe | 9 |
| V2_20 | 99.95% | 98.34% | 2061.30 | H16b | H | 100.00% | No | Europe | 9 |
| HC_21 | 99.95% | 97.38% | 1646.22 | J1c1b1 | J | 98.58% | No | Europe | 32 |
| V1_21 | 99.92% | 95.67% | 1284.49 | H28a | H | 91.40% | No | Europe | 12 |
| V2_21 | 99.91% | 95.57% | 1446.05 | H28a | H | 91.40% | No | Europe | 14 |
| HC_22 | 99.96% | 98.49% | 1722.85 | K1a1b2a1a | K | 94.18% | No | Europe | 41 |
| V1_22 | 99.95% | 97.89% | 1865.40 | H1c | H | 90.83% | - | Europe | 13 |
| V2_22 | 99.95% | 97.78% | 1801.82 | H1c | H | 90.83% | No | Europe | 13 |
| HC_23 | 99.95% | 98.80% | 1550.76 | HV0e | HV | 90.08% | No | Europe | 22 |
| V1_23 | 99.94% | 97.15% | 1580.13 | H | H | 84.45% | No | Europe | 11 |
| V2_23 | 99.95% | 97.92% | 1799.29 | H | H | 84.45% | No | Europe | 11 |
| HC_24 | 99.96% | 96.56% | 1489.69 | H1c1 | H | 94.85% | No | Europe | 13 |
| V1_24 | 99.94% | 97.74% | 2033.71 | K1a+150 | K | 88.85% | No | Europe | 36 |
| V2_24 | 99.95% | 99.02% | 1936.47 | K1a+150 | K | 88.85% | No | Europe | 37 |
| HC_25 | 99.98% | 99.90% | 1922.72 | H3 | H | 87.84% | No | Europe | 11 |
| V1_25 | 99.97% | 99.91% | 2038.19 | H3ae | H | 87.65% | No | Europe | 12 |
| V2_25 | 99.98% | 99.88% | 2049.16 | H3ae | H | 84.17% | No | Europe | 12 |
| HC_26 | 99.98% | 99.87% | 1722.38 | J1c2t | J | 98.51% | No | Europe | 30 |
| V1_26 | 99.99% | 99.80% | 1762.09 | U2e1b1 | U | 96.62% | No | Europe | 38 |
| V2_26 | 99.99% | 99.92% | 1737.76 | U2e1b1 | U | 95.31% | No | Europe | 37 |
| HC_27 | 99.99% | 99.94% | 2503.62 | H | H | 75.99% | No | Europe | 21 |
| V1_27 | 99.99% | 99.94% | 2544.61 | J2b1 | J | 94.21% | No | Europe | 33 |
| V2_27 | 99.99% | 99.96% | 3213.83 | J2b1 | J | 94.21% | No | Europe | 33 |
| HC_28 | 99.97% | 99.91% | 2111.60 | H5a1 | H | 95.32% | No | Europe | 11 |
| V1_28 | 99.98% | 99.93% | 3144.49 | H1c | H | 96.20% | No | Europe | 10 |
| V2_28 | 99.98% | 99.94% | 3281.08 | H1c | H | 96.20% | No | Europe | 12 |
| HC_29 | 99.98% | 99.29% | 2565.71 | V1a | V | 95.13% | No | Europe | 16 |
| V1_29 | 99.98% | 99.95% | 3180.15 | H2a1 | H | 100.00% | No | Europe | 15 |
| V2_29 | 99.99% | 99.96% | 2653.38 | H2a1 | H | 100.00% | No | Europe | 7 |

#### Table S7 — WGS in the CIS/RRMS cohort: Overall look (continued)

| ID | Mappability >30 reads/base | Mappability >100 reads/base | Mean coverage | Haplogroup | Haplogroup (simplified) | Haplogroup quality (%) | Contamination? | Haplogroup origin | Number of variants |
|---|---|---|---|---|---|---|---|---|---|
| HC_30 | 99.63% | 98.03% | 1773.66 | X2c1a | X | 95.54% | No | Europe | 29 |
| V1_30 | 99.92% | 97.73% | 1628.40 | H7 | H | 91.58% | No | Europe | 11 |
| V2_30 | 99.95% | 97.31% | 1737.71 | H7 | H | 91.58% | No | Europe | 11 |
| HC_31 | 99.99% | 99.90% | 2095.59 | H24a | H | 87.49% | No | Europe | 12 |
| V1_31 | 99.97% | 99.86% | 1869.81 | H3*1 | H | 88.86% | No | Europe | 11 |
| V2_31 | 99.98% | 99.87% | 1985.97 | H3*1 | H | 88.86% | No | Europe | 12 |
| HC_32 | 99.99% | 99.93% | 2125.28 | H5u1 | H | 100.00% | No | Europe | 13 |
| V1_32 | 99.98% | 99.90% | 2136.68 | J1c2 | J | 97.95% | No | Europe | 30 |
| V2_32 | 99.98% | 99.93% | 2494.11 | J1c2 | J | 97.95% | No | Europe | 30 |
| HC_33 | 99.98% | 99.93% | 2291.16 | T2e1 | T | 94.25% | No | Europe | 37 |
| V1_33 | 99.67% | 99.61% | 2332.25 | J1b1a1+146 | J | 98.06% | No | Europe | 39 |
| V2_33 | 99.66% | 99.02% | 2119.40 | J1b1a1+146 | J | 98.06% | No | Europe | 40 |
| HC_34 | 99.97% | 99.93% | 2586.54 | H36 | H | 96.12% | No | Europe | 18 |
| V1_34 | 99.98% | 99.81% | 2261.72 | H23 | H | 99.26% | No | Europe | 12 |
| V2_34 | 99.98% | 99.93% | 2496.00 | H23 | H | 99.26% | No | Europe | 9 |
| HC_35 | 99.95% | 97.41% | 2157.32 | H2a1 | H | 78.36% | No | Europe | 14 |
| V1_35 | 99.93% | 99.17% | 2677.27 | K1a4c1 | K | 92.62% | No | Europe | 44 |
| V2_35 | 99.98% | 99.89% | 2100.89 | K1a4c1 | K | 94.21% | No | Europe | 70 |
| HC_36 | 99.98% | 99.23% | 1706.13 | T2b3+151 | T | 96.27% | No | Europe | 36 |
| V1_36 | 99.98% | 99.94% | 2504.74 | H10e | H | 100.00% | No | Europe | 15 |
| V2_36 | 99.98% | 99.93% | 2025.57 | H10e | H | 100.00% | No | Europe | 10 |
| HC_37 | 99.93% | 97.21% | 1523.01 | J2a1a1a2 | J | 100.00% | No | Europe | 41 |
| V1_37 | 99.37% | 97.63% | 2020.24 | H | H | 88.79% | No | Europe | 13 |
| V2_37 | 99.35% | 97.06% | 2039.61 | H | H | 94.05% | No | Europe | 12 |
| HC_38 | 99.90% | 96.02% | 1217.10 | H24 | H | 93.19% | No | Europe | 10 |
| V1_38 | 99.90% | 95.08% | 1363.63 | T2b28 | T | 100.00% | No | Europe | 34 |
| V2_38 | 99.99% | 99.96% | 3217.46 | T2b28 | T | 100.00% | No | Europe | 38 |
| HC_39 | 99.97% | 98.77% | 1828.62 | U5a2a | U | 51.90% | No | Europe | 2 |
| V1_39 | 99.96% | 99.48% | 2013.47 | H1c+152 | H | 90.31% | No | Europe | 14 |
| V2_39 | 99.95% | 99.27% | 2218.68 | H1c+152 | H | 90.31% | No | Europe | 13 |
| HC_40 | 99.98% | 99.95% | 2583.79 | HV11 | HV | 90.91% | No | Europe | 20 |
| V1_40 | 99.97% | 99.92% | 2498.29 | H5a1 | H | 100.00% | No | Europe | 11 |
| V2_40 | 99.98% | 99.92% | 2773.15 | H5a1 | H | 100.00% | No | Europe | 11 |
| HC_41 | 99.98% | 99.95% | 3078.20 | V11 | V | 97.84% | No | Europe | 18 |
| V1_41 | 99.98% | 99.94% | 3391.94 | HV6 | HV | 95.99% | No | Europe | 13 |
| V2_41 | 99.98% | 99.90% | 2774.15 | HV6 | HV | 95.99% | No | Europe | 13 |
| HC_42 | 99.94% | 98.22% | 1639.94 | H11a | H | 96.96% | No | Europe | 11 |
| V1_42 | 99.92% | 99.23% | 1806.80 | K1a4d | K | 98.84% | No | Europe | 39 |
| V2_42 | 99.94% | 98.15% | 1705.32 | K1a4d | K | 98.47% | No | Europe | 37 |
| HC_43 | 99.98% | 99.90% | 1792.53 | H1b2 | H | 100.00% | No | Europe | 13 |
| V1_43 | 99.98% | 99.87% | 1921.85 | H1b* | H | 100.00% | No | Europe | 13 |
| V2_43 | 99.98% | 99.87% | 1703.96 | H1b* | H | 100.00% | No | Europe | 13 |
| HC_44 | 99.95% | 99.64% | 2123.54 | U2e2a1a | U | 95.91% | No | Europe | 39 |
| V1_44 | 99.99% | 99.96% | 3163.42 | HV+16311 | HV | 91.58% | No | Europe | 13 |
| V2_44 | 99.94% | 98.80% | 2052.74 | HV+16311 | HV | 91.58% | No | Europe | 13 |

#### Table S7 — WGS in the CIS/RRMS cohort: Overall look (continued)

| ID | Mappability >30 reads/base | Mappability >100 reads/base | Mean coverage | Haplogroup | Haplogroup (simplified) | Haplogroup quality (%) | Contamination? | Haplogroup origin | Number of variants |
|---|---|---|---|---|---|---|---|---|---|
| HC_45 | 99.94% | 96.21% | 1955.70 | T2b4 | T | 94.76% | No | Europe | 34 |
| V1_45 | 99.96% | 97.62% | 2747.78 | H1b | H | 90.51% | No | Europe | 13 |
| V2_45 | 99.96% | 98.87% | 2496.57 | H1b | H | 92.51% | No | Europe | 13 |
| HC_46 | 99.93% | 97.97% | 1862.67 | K1a4a1b2 | K | 99.29% | No | Europe | 39 |
| V1_46 | 99.93% | 99.16% | 1673.86 | U5a1c1 | U | 100.00% | No | Europe | 27 |
| V2_46 | 99.99% | 99.97% | 3238.84 | U5a1c1 | U | 100.00% | No | Europe | 27 |
| HC_47 | 99.95% | 98.85% | 1869.42 | HV1a1a | HV | 95.91% | No | Europe | 19 |
| V1_47 | 99.96% | 98.20% | 1955.61 | T2b34 | T | 100.00% | No | Europe | 45 |
| V2_47 | 99.97% | 97.41% | 2068.20 | T2b34 | T | 100.00% | No | Europe | 43 |
| HC_48 | 99.99% | 99.95% | 2468.72 | H2a | H | 87.12% | No | Europe | 6 |
| V1_48 | 99.98% | 99.95% | 2609.59 | J1c2e1 | J | 98.74% | No | Europe | 32 |
| V2_48 | 99.96% | 99.93% | 3128.08 | J1c2 | J | 97.79% | No | Europe | 32 |
| HC_49 | 99.98% | 99.94% | 2404.45 | T2d1b1 | T | 95.07% | No | Europe | 38 |
| V1_49 | 99.98% | 99.92% | 2631.74 | H1c2 | H | 90.67% | No | Europe | 15 |
| V2_49 | 99.99% | 99.95% | 2629.15 | H1c2 | H | 90.67% | No | Europe | 15 |
| HC_50 | 99.95% | 96.73% | 1448.76 | U5b3b2 | U | 98.54% | No | Europe | 28 |
| V1_50 | 99.95% | 99.22% | 2772.15 | H1 | H | 90.86% | No | Europe | 10 |
| V2_50 | 99.96% | 98.81% | 2772.45 | H1 | H | 90.86% | No | Europe | 10 |
| HC_51 | 99.98% | 99.54% | 2874.89 | H1a1c | H | 90.95% | No | Europe | 17 |
| V1_51 | 99.97% | 99.93% | 2823.02 | HV | HV | 89.69% | No | Europe | 17 |
| V2_51 | 99.98% | 99.94% | 2755.36 | HV | HV | 89.69% | No | Europe | 12 |
| HC_52 | 99.99% | 99.96% | 2638.26 | I3a | I | 97.06% | No | Europe | 34 |
| V1_52 | 99.98% | 99.93% | 2719.08 | J2b1a | J | 94.43% | No | Europe | 35 |
| V2_52 | 99.98% | 99.95% | 2525.17 | J2b1a | J | 94.43% | No | Europe | 37 |
| HC_53 | 99.98% | 99.94% | 3212.76 | U5a2b1c | U | 94.68% | No | Europe | 27 |
| V1_53 | 99.99% | 99.95% | 2901.51 | U5a1a1+152 | U | 93.79% | No | Europe | 32 |
| V2_53 | 99.98% | 99.93% | 2649.38 | U5a1a1+152 | U | 93.79% | No | Europe | 31 |
| HC_54 | 99.69% | 99.63% | 2145.05 | U2d1 | U | 91.43% | No | Europe | 38 |
| V1_54 | 99.98% | 99.92% | 2165.20 | H56 | H | 100.00% | No | Europe | 9 |
| V2_54 | 99.98% | 99.93% | 2303.96 | H56 | H | 100.00% | No | Europe | 10 |
| HC_55 | 99.96% | 98.70% | 2025.92 | H1a1 | H | 100.00% | No | Europe | 17 |
| V1_55 | 99.96% | 97.88% | 1752.22 | H2a2a1 | H | 50.00% | No | Europe | 1 |
| V2_55 | 99.97% | 99.22% | 1959.85 | H2a2a1 | H | 50.00% | No | Europe | 1 |
| HC_56 | 99.99% | 99.96% | 2586.49 | H+152 | H | 96.01% | No | Europe | 12 |
| V1_56 | 99.98% | 99.95% | 2428.34 | H24a | H | 100.00% | No | Europe | 11 |
| V2_56 | 99.99% | 99.95% | 2530.23 | H24a | H | 100.00% | No | Europe | 11 |
| HC_57 | 99.90% | 96.83% | 1535.55 | U4a | U | 96.38% | No | Europe | 27 |
| V1_57 | 99.12% | 94.30% | 1275.44 | K1a1b1f | K | 98.74% | No | Europe | 41 |
| V2_57 | 99.14% | 94.87% | 1307.83 | K1a1b1f | K | 97.25% | No | Europe | 38 |
| HC_58 | 99.95% | 98.44% | 1607.76 | T2b | T | 96.02% | No | Europe | 39 |
| V1_58 | 99.93% | 99.05% | 1776.84 | U5b2a2b | U | 98.50% | No | Europe | 33 |
| V2_58 | 99.96% | 97.56% | 1626.77 | U5b2a2b | U | 98.50% | No | Europe | 33 |
| HC_59 | 99.96% | 98.26% | 1838.69 | H1a | H | 86.75% | No | Europe | 17 |
| V1_59 | 99.95% | 99.52% | 2000.74 | H5a1 | H | 100.00% | No | Europe | 11 |
| V2_59 | 99.96% | 99.49% | 2015.76 | H5a1 | H | 100.00% | No | Europe | 13 |

#### Table S7 — WGS in the CIS/RRMS cohort: Overall look (continued)

| ID | Mappability >30 reads/base | Mappability >100 reads/base | Mean coverage | Haplogroup | Haplogroup (simplified) | Haplogroup quality (%) | Contamination? | Haplogroup origin | Number of variants |
|---|---|---|---|---|---|---|---|---|---|
| HC_60 | 99.97% | 99.86% | 1828.78 | V7a | V | 96.86% | No | Europe | 18 |
| V1_60 | 99.97% | 99.91% | 2025.34 | H85 | H | 95.09% | No | Europe | 12 |
| V2_60 | 99.98% | 99.92% | 2180.39 | H85 | H | 95.09% | No | Europe | 11 |
| HC_61 | 99.95% | 97.28% | 1334.85 | H1a3a | H | 99.06% | No | Europe | 13 |
| V1_61 | 99.95% | 98.04% | 1743.89 | H1a3a3 | H | 97.38% | No | Europe | 18 |
| V2_61 | 99.93% | 97.24% | 1436.93 | H1a3a3 | H | 97.38% | No | Europe | 17 |

Abbreviations: CIS — Clinically Isolated Syndrome; HC — healthy control; MS — Multiple Sclerosis; WGS — whole genome sequencing; V1 — patient with Clinically Isolated Syndrome/Relapsing-Remitting Multiple Sclerosis at visit 1; V2 — patient with Clinically Isolated Syndrome/Relapsing-Remitting Multiple Sclerosis at visit 2.

#### Table S8 — Mean number of mutations per haplogroup (simplified). Dunn test, after a Kruskal-Wallis test, with FDR

| Haplogroup (simplified) | Mean number of mutations | Adjusted *p*-value |
|---|---|---|
| I ~ J | 38.33 ~ 34.00 | 6.11E-01 |
| I ~ K | 38.33 ~ 37.80 | 9.30E-01 |
| I ~ T | 38.33 ~ 37.44 | 9.30E-01 |
| I ~ U | 38.33 ~ 29.79 | 3.48E-01 |
| I ~ V | 38.33 ~ 17.40 | 1.48E-01 |
| K ~ T | 37.80 ~ 37.44 | 9.30E-01 |
| K ~ U | 37.80 ~ 29.79 | 1.84E-01 |
| K ~ V | 37.80 ~ 17.40 | 6.26E-02 |
| T ~ U | 37.44 ~ 29.79 | 2.17E-01 |
| T ~ V | 37.44 ~ 17.40 | 7.92E-02 |
| J ~ K | 34.00 ~ 37.80 | 5.14E-01 |
| J ~ T | 34.00 ~ 37.44 | 5.97E-01 |
| J ~ U | 34.00 ~ 29.79 | 4.94E-01 |
| J ~ V | 34.00 ~ 17.40 | 1.56E-01 |
| U ~ V | 29.79 ~ 17.40 | 3.48E-01 |
| HV ~ I | 17.33 ~ 38.33 | 8.90E-02 |
| HV ~ J | 17.33 ~ 34.00 | 7.92E-02 |
| HV ~ K | 17.33 ~ 37.80 | 2.57E-02 |
| HV ~ T | 17.33 ~ 37.44 | 3.51E-02 |
| HV ~ U | 17.33 ~ 29.79 | 2.17E-01 |
| HV ~ V | 17.33 ~ 17.40 | 9.20E-01 |
| H ~ HV | 13.05 ~ 17.33 | 2.18E-01 |
| H ~ I | 13.05 ~ 38.33 | 1.87E-03 |
| H ~ J | 13.05 ~ 34.00 | 2.03E-07 |
| H ~ K | 13.05 ~ 37.80 | 4.28E-08 |
| H ~ T | 13.05 ~ 37.44 | 2.03E-07 |
| H ~ U | 13.05 ~ 29.79 | 2.84E-05 |
| H ~ V | 13.05 ~ 17.40 | 1.84E-01 |

Significant *p*-values are highlighted. Abbreviations: FDR — false discovery rate.

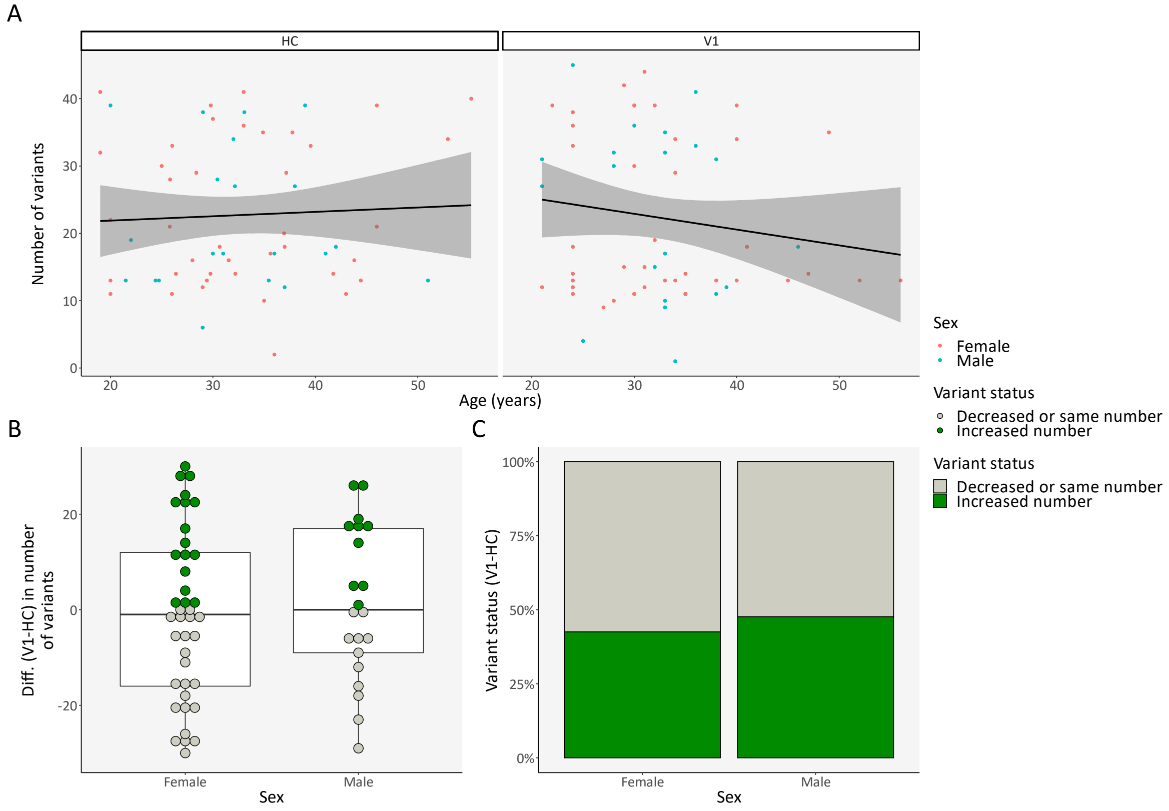

#### Figure S10 — Total number of variants: Cross-sectional comparison (age and sex)

(**A**) Correlation between number of variants and age, per subject type and sex; (**B**) Difference in number of variants per triplet (PwMS at VIS1-HC), per sex; (**C**) Proportion of triplets with an increased or equal/decreased number of variants, per sex. Abbreviations: Diff. — difference; HC — healthy control; PwMS — patient with Clinically Isolated Syndrome/Relapsing-Remitting Multiple Sclerosis; V1 — patient with Clinically Isolated Syndrome/Relapsing-Remitting Multiple Sclerosis at visit 1; VIS1 — visit 1.

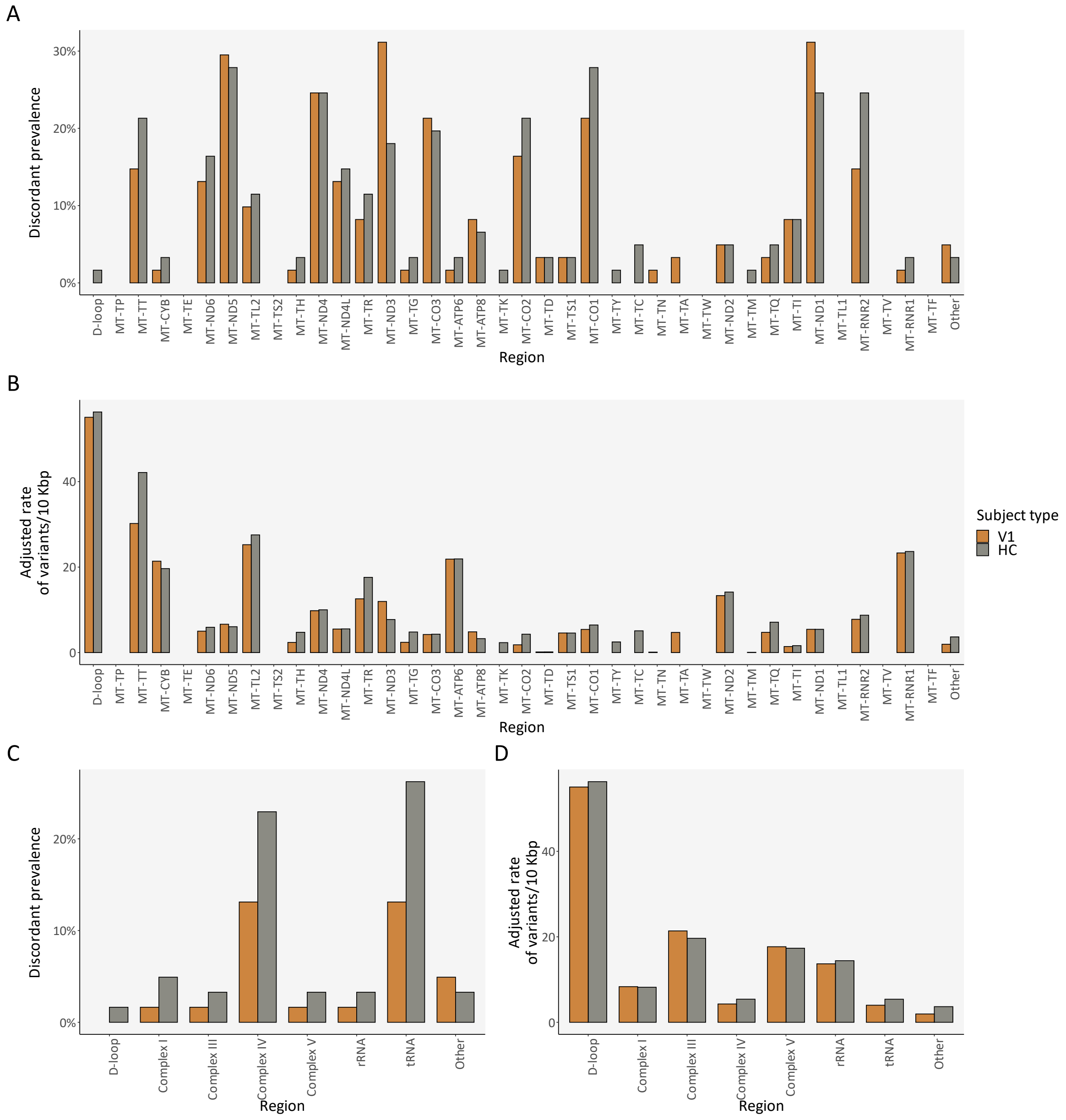

**Figure S11 — Total number of variants: Cross-sectional comparison (regions)**

(**A,C**) Discordant prevalence of variants for each mtDNA region/locus and for each macro mtDNA region, per subject type, respectively; (**B,D**) Relative variant burden for each mtDNA region/locus and for each macro mtDNA region, per subject type, respectively. Region *Other* refers to positions in rCRS with an overlap between the two strands or unannotated. Abbreviations: bp — base pair; HC — healthy control; V1 — patient with Clinically Isolated Syndrome/Relapsing-Remitting Multiple Sclerosis at visit 1.

#### Table S9 — Mutation distribution between HC and PwMS at VIS1. McNemar's test with FDR

| Mutation | HC mutated & V1 non-mutated | HC & V1 mutated | HC & V1 non-mutated | V1 mutated & HC non-mutated | *p*-value | *p*-value (adjusted) | V1 mutation (discordant) | HC mutation (discordant) | V1-HC | Literature |
|---|---|---|---|---|---|---|---|---|---|---|
| 12612G | 3 | 0 | 47 | 11 | 6.14E-02 | 1 | 18.03% | 4.92% | 13.11% | - |
| 3010A | 7 | 5 | 34 | 15 | 1.36E-01 | 1 | 24.59% | 11.48% | 13.11% | - |
| 14798C | 4 | 2 | 44 | 11 | 1.21E-01 | 1 | 18.03% | 6.56% | 11.48% | MS (risk) [54] |
| 16069T | 3 | 0 | 48 | 10 | 9.61E-02 | 1 | 16.39% | 4.92% | 11.48% | - |
| 295T | 3 | 0 | 48 | 10 | 9.61E-02 | 1 | 16.39% | 4.92% | 11.48% | - |
| 489C | 3 | 0 | 48 | 10 | 9.61E-02 | 1 | 16.39% | 4.92% | 11.48% | - |
| 10398G | 9 | 2 | 35 | 15 | 3.07E-01 | 1 | 24.59% | 14.75% | 9.84% | MS (risk) [55] |
| 11251G | 7 | 1 | 40 | 13 | 2.64E-01 | 1 | 21.31% | 11.48% | 9.84% | - |
| 15452A | 7 | 1 | 40 | 13 | 2.64E-01 | 1 | 21.31% | 11.48% | 9.84% | - |
| 16126C | 7 | 2 | 39 | 13 | 2.64E-01 | 1 | 21.31% | 11.48% | 9.84% | - |
| 4216C | 7 | 1 | 40 | 13 | 2.64E-01 | 1 | 21.31% | 11.48% | 9.84% | MS (risk) [56] |
| 462T | 2 | 0 | 51 | 8 | 1.14E-01 | 1 | 13.11% | 3.28% | 9.84% | - |
| 13708A | 4 | 1 | 47 | 9 | 2.67E-01 | 1 | 14.75% | 6.56% | 8.20% | MS (risk) [57] |
| 146C | 3 | 0 | 51 | 7 | 3.43E-01 | 1 | 11.48% | 4.92% | 6.56% | - |
| 185A | 2 | 1 | 52 | 6 | 2.89E-01 | 1 | 9.84% | 3.28% | 6.56% | - |
| 8021G | 0 | 0 | 57 | 4 | 1.34E-01 | 1 | 6.56% | 0.00% | 6.56% | - |
| 8065A | 0 | 0 | 57 | 4 | 1.34E-01 | 1 | 6.56% | 0.00% | 6.56% | - |
| 16311C | 6 | 3 | 43 | 9 | 6.06E-01 | 1 | 14.75% | 9.84% | 4.92% | - |
| 228A | 3 | 0 | 52 | 6 | 5.05E-01 | 1 | 9.84% | 4.92% | 4.92% | - |
| 477C | 1 | 0 | 56 | 4 | 3.71E-01 | 1 | 6.56% | 1.64% | 4.92% | - |
| 9055A | 3 | 1 | 51 | 6 | 5.05E-01 | 1 | 9.84% | 4.92% | 4.92% | MS (risk) [58] |
| 10172A | 0 | 0 | 59 | 2 | 4.80E-01 | 1 | 3.28% | 0.00% | 3.28% | - |
| 10237C | 0 | 0 | 59 | 2 | 4.80E-01 | 1 | 3.28% | 0.00% | 3.28% | - |
| 10550G | 3 | 1 | 52 | 5 | 7.24E-01 | 1 | 8.20% | 4.92% | 3.28% | - |
| 11299C | 3 | 1 | 52 | 5 | 7.24E-01 | 1 | 8.20% | 4.92% | 3.28% | - |
| 1189C | 3 | 1 | 52 | 5 | 7.24E-01 | 1 | 8.20% | 4.92% | 3.28% | - |

#### Table S9 — Mutation distribution between HC and PwMS at VIS1. McNemar's test with FDR (continued)

| Mutation | HC mutated & V1 non-mutated | HC & V1 mutated | HC & V1 non-mutated | V1 mutated & HC non-mutated | *p*-value | *p*-value (adjusted) | V1 mutation (discordant) | HC mutation (discordant) | V1-HC | Literature |
|---|---|---|---|---|---|---|---|---|---|---|
| 12850G | 0 | 0 | 59 | 2 | 4.80E-01 | 1 | 3.28% | 0.00% | 3.28% | - |
| 13879C | 0 | 0 | 59 | 2 | 4.80E-01 | 1 | 3.28% | 0.00% | 3.28% | - |
| 13934T | 0 | 0 | 59 | 2 | 4.80E-01 | 1 | 3.28% | 0.00% | 3.28% | - |
| 14167T | 3 | 1 | 52 | 5 | 7.24E-01 | 1 | 8.20% | 4.92% | 3.28% | - |
| 15812A | 0 | 0 | 59 | 2 | 4.80E-01 | 1 | 3.28% | 0.00% | 3.28% | - |
| 16193T | 0 | 0 | 59 | 2 | 4.80E-01 | 1 | 3.28% | 0.00% | 3.28% | - |
| 16222T | 0 | 0 | 59 | 2 | 4.80E-01 | 1 | 3.28% | 0.00% | 3.28% | - |
| 16224C | 3 | 1 | 52 | 5 | 7.24E-01 | 1 | 8.20% | 4.92% | 3.28% | - |
| 16296T | 2 | 0 | 55 | 4 | 6.83E-01 | 1 | 6.56% | 3.28% | 3.28% | - |
| 16390A | 0 | 0 | 59 | 2 | 4.80E-01 | 1 | 3.28% | 0.00% | 3.28% | - |
| 16399G | 0 | 0 | 59 | 2 | 4.80E-01 | 1 | 3.28% | 0.00% | 3.28% | - |
| 16519C | 14 | 26 | 5 | 16 | 8.55E-01 | 1 | 26.23% | 22.95% | 3.28% | - |
| 188G | 2 | 0 | 55 | 4 | 6.83E-01 | 1 | 6.56% | 3.28% | 3.28% | - |
| 2158C | 0 | 0 | 59 | 2 | 4.80E-01 | 1 | 3.28% | 0.00% | 3.28% | - |
| 242T | 0 | 0 | 59 | 2 | 4.80E-01 | 1 | 3.28% | 0.00% | 3.28% | - |
| 3480G | 3 | 1 | 52 | 5 | 7.24E-01 | 1 | 8.20% | 4.92% | 3.28% | - |
| 4732G | 0 | 0 | 59 | 2 | 4.80E-01 | 1 | 3.28% | 0.00% | 3.28% | - |
| 5460A | 1 | 0 | 57 | 3 | 6.17E-01 | 1 | 4.92% | 1.64% | 3.28% | - |
| 5580C | 0 | 0 | 59 | 2 | 4.80E-01 | 1 | 3.28% | 0.00% | 3.28% | - |
| 5633T | 0 | 0 | 59 | 2 | 4.80E-01 | 1 | 3.28% | 0.00% | 3.28% | - |
| 8269A | 0 | 0 | 59 | 2 | 4.80E-01 | 1 | 3.28% | 0.00% | 3.28% | - |
| 8557A | 0 | 0 | 59 | 2 | 4.80E-01 | 1 | 3.28% | 0.00% | 3.28% | - |
| 8701G | 0 | 0 | 59 | 2 | 4.80E-01 | 1 | 3.28% | 0.00% | 3.28% | - |
| 8715C | 0 | 0 | 59 | 2 | 4.80E-01 | 1 | 3.28% | 0.00% | 3.28% | - |
| 8718G | 0 | 0 | 59 | 2 | 4.80E-01 | 1 | 3.28% | 0.00% | 3.28% | - |
| 9629G | 0 | 0 | 59 | 2 | 4.80E-01 | 1 | 3.28% | 0.00% | 3.28% | - |
| 9698C | 3 | 1 | 52 | 5 | 7.24E-01 | 1 | 8.20% | 4.92% | 3.28% | - |
| 10169T | 0 | 0 | 60 | 1 | 1.00E+00 | 1 | 1.64% | 0.00% | 1.64% | - |

**Table S9 — Mutation distribution between HC and PwMS at VIS1. McNemar's test with FDR (continued)**

| Mutation | HC mutated & V1 non-mutated | HC & V1 mutated | HC & V1 non-mutated | V1 mutated & HC non-mutated | *p*-value | *p*-value (adjusted) | V1 mutation (discordant) | HC mutation (discordant) | V1-HC | Literature |
|---|---|---|---|---|---|---|---|---|---|---|
| 10211T | 0 | 0 | 60 | 1 | 1.00E+00 | 1 | 1.64% | 0.00% | 1.64% | - |
| 10232G | 0 | 0 | 60 | 1 | 1.00E+00 | 1 | 1.64% | 0.00% | 1.64% | - |
| 10394T | 0 | 0 | 60 | 1 | 1.00E+00 | 1 | 1.64% | 0.00% | 1.64% | - |
| 10410C | 0 | 0 | 60 | 1 | 1.00E+00 | 1 | 1.64% | 0.00% | 1.64% | - |
| 10632C | 0 | 0 | 60 | 1 | 1.00E+00 | 1 | 1.64% | 0.00% | 1.64% | - |
| 10680A | 0 | 0 | 60 | 1 | 1.00E+00 | 1 | 1.64% | 0.00% | 1.64% | - |
| 10698T | 0 | 0 | 60 | 1 | 1.00E+00 | 1 | 1.64% | 0.00% | 1.64% | - |
| 11050C | 0 | 0 | 60 | 1 | 1.00E+00 | 1 | 1.64% | 0.00% | 1.64% | - |
| 11071T | 0 | 0 | 60 | 1 | 1.00E+00 | 1 | 1.64% | 0.00% | 1.64% | - |
| 11176A | 0 | 0 | 60 | 1 | 1.00E+00 | 1 | 1.64% | 0.00% | 1.64% | - |
| 11191T | 0 | 0 | 60 | 1 | 1.00E+00 | 1 | 1.64% | 0.00% | 1.64% | - |
| 11204C | 0 | 0 | 60 | 1 | 1.00E+00 | 1 | 1.64% | 0.00% | 1.64% | - |
| 11410C | 0 | 0 | 60 | 1 | 1.00E+00 | 1 | 1.64% | 0.00% | 1.64% | - |
| 11428T | 0 | 0 | 60 | 1 | 1.00E+00 | 1 | 1.64% | 0.00% | 1.64% | - |
| 11470G | 0 | 0 | 60 | 1 | 1.00E+00 | 1 | 1.64% | 0.00% | 1.64% | - |
| 11485C | 1 | 0 | 58 | 2 | 1.00E+00 | 1 | 3.28% | 1.64% | 1.64% | - |
| 11563G | 0 | 0 | 60 | 1 | 1.00E+00 | 1 | 1.64% | 0.00% | 1.64% | - |
| 11617C | 0 | 0 | 60 | 1 | 1.00E+00 | 1 | 1.64% | 0.00% | 1.64% | - |
| 11642A | 0 | 0 | 60 | 1 | 1.00E+00 | 1 | 1.64% | 0.00% | 1.64% | - |
| 11719A | 15 | 10 | 20 | 16 | 1.00E+00 | 1 | 26.23% | 24.59% | 1.64% | - |
| 11778A | 0 | 0 | 60 | 1 | 1.00E+00 | 1 | 1.64% | 0.00% | 1.64% | NARP, LHON [59] |
| 11788T | 0 | 0 | 60 | 1 | 1.00E+00 | 1 | 1.64% | 0.00% | 1.64% | - |
| 12083G | 0 | 0 | 60 | 1 | 1.00E+00 | 1 | 1.64% | 0.00% | 1.64% | - |
| 12738G | 0 | 0 | 60 | 1 | 1.00E+00 | 1 | 1.64% | 0.00% | 1.64% | - |
| 12771A | 0 | 0 | 60 | 1 | 1.00E+00 | 1 | 1.64% | 0.00% | 1.64% | - |
| 12813T | 0 | 0 | 60 | 1 | 1.00E+00 | 1 | 1.64% | 0.00% | 1.64% | - |
| 12814A | 0 | 0 | 60 | 1 | 1.00E+00 | 1 | 1.64% | 0.00% | 1.64% | - |
| 12858T | 0 | 0 | 60 | 1 | 1.00E+00 | 1 | 1.64% | 0.00% | 1.64% | - |

#### Table S9 — Mutation distribution between HC and PwMS at VIS1. McNemar's test with FDR (continued)

| Mutation | HC mutated & V1 non-mutated | HC & V1 mutated | HC & V1 non-mutated | V1 mutated & HC non-mutated | *p*-value | *p*-value (adjusted) | V1 mutation (discordant) | HC mutation (discordant) | V1-HC | Literature |
|---|---|---|---|---|---|---|---|---|---|---|
| 13064C | 0 | 0 | 60 | 1 | 1.00E+00 | 1 | 1.64% | 0.00% | 1.64% | - |
| 13134G | 0 | 0 | 60 | 1 | 1.00E+00 | 1 | 1.64% | 0.00% | 1.64% | - |
| 13145A | 0 | 0 | 60 | 1 | 1.00E+00 | 1 | 1.64% | 0.00% | 1.64% | - |
| 13191C | 0 | 0 | 60 | 1 | 1.00E+00 | 1 | 1.64% | 0.00% | 1.64% | - |
| 13281C | 0 | 0 | 60 | 1 | 1.00E+00 | 1 | 1.64% | 0.00% | 1.64% | - |
| 13590A | 0 | 0 | 60 | 1 | 1.00E+00 | 1 | 1.64% | 0.00% | 1.64% | - |
| 13637G | 0 | 1 | 59 | 1 | 1.00E+00 | 1 | 1.64% | 0.00% | 1.64% | - |
| 13710G | 0 | 0 | 60 | 1 | 1.00E+00 | 1 | 1.64% | 0.00% | 1.64% | - |
| 13781C | 0 | 0 | 60 | 1 | 1.00E+00 | 1 | 1.64% | 0.00% | 1.64% | - |
| 13782A | 0 | 0 | 60 | 1 | 1.00E+00 | 1 | 1.64% | 0.00% | 1.64% | - |
| 13802T | 0 | 0 | 60 | 1 | 1.00E+00 | 1 | 1.64% | 0.00% | 1.64% | - |
| 13827G | 0 | 0 | 60 | 1 | 1.00E+00 | 1 | 1.64% | 0.00% | 1.64% | - |
| 13854T | 0 | 0 | 60 | 1 | 1.00E+00 | 1 | 1.64% | 0.00% | 1.64% | - |
| 13933G | 0 | 0 | 60 | 1 | 1.00E+00 | 1 | 1.64% | 0.00% | 1.64% | - |
| 14016A | 0 | 0 | 60 | 1 | 1.00E+00 | 1 | 1.64% | 0.00% | 1.64% | - |
| 14097T | 0 | 0 | 60 | 1 | 1.00E+00 | 1 | 1.64% | 0.00% | 1.64% | - |
| 14153C | 0 | 0 | 60 | 1 | 1.00E+00 | 1 | 1.64% | 0.00% | 1.64% | - |
| 14218C | 0 | 0 | 60 | 1 | 1.00E+00 | 1 | 1.64% | 0.00% | 1.64% | - |
| 14370G | 0 | 0 | 60 | 1 | 1.00E+00 | 1 | 1.64% | 0.00% | 1.64% | - |
| 14386C | 0 | 0 | 60 | 1 | 1.00E+00 | 1 | 1.64% | 0.00% | 1.64% | - |
| 14470A | 0 | 0 | 60 | 1 | 1.00E+00 | 1 | 1.64% | 0.00% | 1.64% | - |
| 14766T | 15 | 10 | 20 | 16 | 1.00E+00 | 1 | 26.23% | 24.59% | 1.64% | - |
| 14893C | 0 | 0 | 60 | 1 | 1.00E+00 | 1 | 1.64% | 0.00% | 1.64% | - |
| 14953T | 0 | 0 | 60 | 1 | 1.00E+00 | 1 | 1.64% | 0.00% | 1.64% | - |
| 14956C | 0 | 0 | 60 | 1 | 1.00E+00 | 1 | 1.64% | 0.00% | 1.64% | - |
| 15028A | 0 | 0 | 60 | 1 | 1.00E+00 | 1 | 1.64% | 0.00% | 1.64% | - |
| 15067C | 0 | 0 | 60 | 1 | 1.00E+00 | 1 | 1.64% | 0.00% | 1.64% | - |
| 15172A | 0 | 0 | 60 | 1 | 1.00E+00 | 1 | 1.64% | 0.00% | 1.64% | - |

**Table S9 — Mutation distribution between HC and PwMS at VIS1. McNemar's test with FDR (continued)**

| Mutation | HC mutated & V1 non-mutated | HC & V1 mutated | HC & V1 non-mutated | V1 mutated & HC non-mutated | *p*-value | *p*-value (adjusted) | V1 mutation (discordant) | HC mutation (discordant) | V1-HC | Literature |
|---|---|---|---|---|---|---|---|---|---|---|
| 15218G | 1 | 0 | 58 | 2 | 1.00E+00 | 1 | 3.28% | 1.64% | 1.64% | - |
| 15257A | 1 | 0 | 58 | 2 | 1.00E+00 | 1 | 3.28% | 1.64% | 1.64% | MS (risk) [60,61] |
| 15262C | 0 | 0 | 60 | 1 | 1.00E+00 | 1 | 1.64% | 0.00% | 1.64% | - |
| 15388C | 0 | 0 | 60 | 1 | 1.00E+00 | 1 | 1.64% | 0.00% | 1.64% | - |
| 15401G | 0 | 0 | 60 | 1 | 1.00E+00 | 1 | 1.64% | 0.00% | 1.64% | - |
| 15440C | 0 | 0 | 60 | 1 | 1.00E+00 | 1 | 1.64% | 0.00% | 1.64% | - |
| 15449C | 0 | 0 | 60 | 1 | 1.00E+00 | 1 | 1.64% | 0.00% | 1.64% | - |
| 15511C | 0 | 0 | 60 | 1 | 1.00E+00 | 1 | 1.64% | 0.00% | 1.64% | - |
| 15784C | 0 | 0 | 60 | 1 | 1.00E+00 | 1 | 1.64% | 0.00% | 1.64% | - |
| 15906G | 0 | 0 | 60 | 1 | 1.00E+00 | 1 | 1.64% | 0.00% | 1.64% | - |
| 15914G | 0 | 0 | 60 | 1 | 1.00E+00 | 1 | 1.64% | 0.00% | 1.64% | - |
| 15930A | 0 | 0 | 60 | 1 | 1.00E+00 | 1 | 1.64% | 0.00% | 1.64% | - |
| 16093C | 3 | 0 | 54 | 4 | 1.00E+00 | 1 | 6.56% | 4.92% | 1.64% | - |
| 16111T | 0 | 0 | 60 | 1 | 1.00E+00 | 1 | 1.64% | 0.00% | 1.64% | - |
| 16145A | 2 | 0 | 56 | 3 | 1.00E+00 | 1 | 4.92% | 3.28% | 1.64% | - |
| 16147T | 0 | 0 | 60 | 1 | 1.00E+00 | 1 | 1.64% | 0.00% | 1.64% | - |
| 16180G | 0 | 0 | 60 | 1 | 1.00E+00 | 1 | 1.64% | 0.00% | 1.64% | - |
| 16188T | 0 | 0 | 60 | 1 | 1.00E+00 | 1 | 1.64% | 0.00% | 1.64% | - |
| 16217C | 0 | 0 | 60 | 1 | 1.00E+00 | 1 | 1.64% | 0.00% | 1.64% | - |
| 16221T | 0 | 0 | 60 | 1 | 1.00E+00 | 1 | 1.64% | 0.00% | 1.64% | - |
| 16239T | 0 | 0 | 60 | 1 | 1.00E+00 | 1 | 1.64% | 0.00% | 1.64% | - |
| 16245T | 0 | 0 | 60 | 1 | 1.00E+00 | 1 | 1.64% | 0.00% | 1.64% | - |
| 16246T | 0 | 0 | 60 | 1 | 1.00E+00 | 1 | 1.64% | 0.00% | 1.64% | - |
| 16249C | 0 | 0 | 60 | 1 | 1.00E+00 | 1 | 1.64% | 0.00% | 1.64% | - |
| 16256T | 1 | 1 | 57 | 2 | 1.00E+00 | 1 | 3.28% | 1.64% | 1.64% | - |
| 16259T | 0 | 0 | 60 | 1 | 1.00E+00 | 1 | 1.64% | 0.00% | 1.64% | - |
| 16260T | 0 | 0 | 60 | 1 | 1.00E+00 | 1 | 1.64% | 0.00% | 1.64% | - |
| 16262T | 0 | 0 | 60 | 1 | 1.00E+00 | 1 | 1.64% | 0.00% | 1.64% | - |

#### Table S9 — Mutation distribution between HC and PwMS at VIS1. McNemar's test with FDR (continued)

| Mutation | HC mutated & V1 non-mutated | HC & V1 mutated | HC & V1 non-mutated | V1 mutated & HC non-mutated | *p*-value | *p*-value (adjusted) | V1 mutation (discordant) | HC mutation (discordant) | V1-HC | Literature |
|---|---|---|---|---|---|---|---|---|---|---|
| 16265G | 0 | 0 | 60 | 1 | 1.00E+00 | 1 | 1.64% | 0.00% | 1.64% | - |
| 16266T | 0 | 0 | 60 | 1 | 1.00E+00 | 1 | 1.64% | 0.00% | 1.64% | - |
| 16271C | 0 | 0 | 60 | 1 | 1.00E+00 | 1 | 1.64% | 0.00% | 1.64% | - |
| 16287T | 0 | 0 | 60 | 1 | 1.00E+00 | 1 | 1.64% | 0.00% | 1.64% | - |
| 16291T | 0 | 0 | 60 | 1 | 1.00E+00 | 1 | 1.64% | 0.00% | 1.64% | - |
| 16294A | 0 | 0 | 60 | 1 | 1.00E+00 | 1 | 1.64% | 0.00% | 1.64% | - |
| 16325C | 0 | 0 | 60 | 1 | 1.00E+00 | 1 | 1.64% | 0.00% | 1.64% | - |
| 16360T | 0 | 0 | 60 | 1 | 1.00E+00 | 1 | 1.64% | 0.00% | 1.64% | - |
| 16366T | 0 | 0 | 60 | 1 | 1.00E+00 | 1 | 1.64% | 0.00% | 1.64% | - |
| 16368C | 0 | 0 | 60 | 1 | 1.00E+00 | 1 | 1.64% | 0.00% | 1.64% | - |
| 16398A | 0 | 0 | 60 | 1 | 1.00E+00 | 1 | 1.64% | 0.00% | 1.64% | - |
| 1721T | 0 | 1 | 59 | 1 | 1.00E+00 | 1 | 1.64% | 0.00% | 1.64% | - |
| 186A | 0 | 0 | 60 | 1 | 1.00E+00 | 1 | 1.64% | 0.00% | 1.64% | - |
| 199C | 4 | 0 | 52 | 5 | 1.00E+00 | 1 | 8.20% | 6.56% | 1.64% | - |
| 2028A | 0 | 0 | 60 | 1 | 1.00E+00 | 1 | 1.64% | 0.00% | 1.64% | - |
| 214G | 0 | 0 | 60 | 1 | 1.00E+00 | 1 | 1.64% | 0.00% | 1.64% | - |
| 2405A | 0 | 0 | 60 | 1 | 1.00E+00 | 1 | 1.64% | 0.00% | 1.64% | - |
| 263G | 2 | 56 | 0 | 3 | 1.00E+00 | 1 | 4.92% | 3.28% | 1.64% | - |
| 2757G | 0 | 0 | 60 | 1 | 1.00E+00 | 1 | 1.64% | 0.00% | 1.64% | - |
| 2857C | 0 | 0 | 60 | 1 | 1.00E+00 | 1 | 1.64% | 0.00% | 1.64% | - |
| 3212T | 0 | 0 | 60 | 1 | 1.00E+00 | 1 | 1.64% | 0.00% | 1.64% | - |
| 327T | 0 | 0 | 60 | 1 | 1.00E+00 | 1 | 1.64% | 0.00% | 1.64% | - |
| 3335C | 0 | 0 | 60 | 1 | 1.00E+00 | 1 | 1.64% | 0.00% | 1.64% | - |
| 3337A | 0 | 0 | 60 | 1 | 1.00E+00 | 1 | 1.64% | 0.00% | 1.64% | - |
| 3337C | 0 | 0 | 60 | 1 | 1.00E+00 | 1 | 1.64% | 0.00% | 1.64% | - |
| 3396C | 0 | 0 | 60 | 1 | 1.00E+00 | 1 | 1.64% | 0.00% | 1.64% | - |
| 340T | 0 | 0 | 60 | 1 | 1.00E+00 | 1 | 1.64% | 0.00% | 1.64% | - |
| 3507T | 0 | 0 | 60 | 1 | 1.00E+00 | 1 | 1.64% | 0.00% | 1.64% | - |

#### Table S9 — Mutation distribution between HC and PwMS at VIS1. McNemar's test with FDR (continued)

| Mutation | HC mutated & V1 non-mutated | HC & V1 mutated | HC & V1 non-mutated | V1 mutated & HC non-mutated | *p*-value | *p*-value (adjusted) | V1 mutation (discordant) | HC mutation (discordant) | V1-HC | Literature |
|---|---|---|---|---|---|---|---|---|---|---|
| 372C | 0 | 0 | 60 | 1 | 1.00E+00 | 1 | 1.64% | 0.00% | 1.64% | - |
| 3796G | 0 | 0 | 60 | 1 | 1.00E+00 | 1 | 1.64% | 0.00% | 1.64% | - |
| 3912G | 0 | 0 | 60 | 1 | 1.00E+00 | 1 | 1.64% | 0.00% | 1.64% | - |
| 4502C | 0 | 0 | 60 | 1 | 1.00E+00 | 1 | 1.64% | 0.00% | 1.64% | - |
| 4634C | 0 | 0 | 60 | 1 | 1.00E+00 | 1 | 1.64% | 0.00% | 1.64% | - |
| 4755C | 0 | 0 | 60 | 1 | 1.00E+00 | 1 | 1.64% | 0.00% | 1.64% | - |
| 4823C | 0 | 0 | 60 | 1 | 1.00E+00 | 1 | 1.64% | 0.00% | 1.64% | - |
| 508G | 1 | 0 | 58 | 2 | 1.00E+00 | 1 | 3.28% | 1.64% | 1.64% | - |
| 5264T | 0 | 0 | 60 | 1 | 1.00E+00 | 1 | 1.64% | 0.00% | 1.64% | - |
| 5277C | 0 | 0 | 60 | 1 | 1.00E+00 | 1 | 1.64% | 0.00% | 1.64% | - |
| 5302C | 0 | 0 | 60 | 1 | 1.00E+00 | 1 | 1.64% | 0.00% | 1.64% | - |
| 5351G | 0 | 0 | 60 | 1 | 1.00E+00 | 1 | 1.64% | 0.00% | 1.64% | - |
| 5426C | 1 | 0 | 58 | 2 | 1.00E+00 | 1 | 3.28% | 1.64% | 1.64% | - |
| 5437T | 0 | 0 | 60 | 1 | 1.00E+00 | 1 | 1.64% | 0.00% | 1.64% | - |
| 5495C | 0 | 0 | 60 | 1 | 1.00E+00 | 1 | 1.64% | 0.00% | 1.64% | - |
| 55C | 0 | 0 | 60 | 1 | 1.00E+00 | 1 | 1.64% | 0.00% | 1.64% | - |
| 5664G | 0 | 0 | 60 | 1 | 1.00E+00 | 1 | 1.64% | 0.00% | 1.64% | - |
| 57C | 0 | 0 | 60 | 1 | 1.00E+00 | 1 | 1.64% | 0.00% | 1.64% | - |
| 5913A | 0 | 0 | 60 | 1 | 1.00E+00 | 1 | 1.64% | 0.00% | 1.64% | - |
| 6023A | 0 | 0 | 60 | 1 | 1.00E+00 | 1 | 1.64% | 0.00% | 1.64% | - |
| 6119T | 0 | 0 | 60 | 1 | 1.00E+00 | 1 | 1.64% | 0.00% | 1.64% | - |
| 6253C | 0 | 0 | 60 | 1 | 1.00E+00 | 1 | 1.64% | 0.00% | 1.64% | - |
| 6266G | 0 | 0 | 60 | 1 | 1.00E+00 | 1 | 1.64% | 0.00% | 1.64% | - |
| 6302G | 0 | 0 | 60 | 1 | 1.00E+00 | 1 | 1.64% | 0.00% | 1.64% | - |
| 6323G | 0 | 0 | 60 | 1 | 1.00E+00 | 1 | 1.64% | 0.00% | 1.64% | - |
| 6465A | 0 | 0 | 60 | 1 | 1.00E+00 | 1 | 1.64% | 0.00% | 1.64% | - |
| 6489A | 0 | 0 | 60 | 1 | 1.00E+00 | 1 | 1.64% | 0.00% | 1.64% | - |
| 6528T | 0 | 0 | 60 | 1 | 1.00E+00 | 1 | 1.64% | 0.00% | 1.64% | - |

#### Table S9 — Mutation distribution between HC and PwMS at VIS1. McNemar's test with FDR (continued)

| Mutation | HC mutated & V1 non-mutated | HC & V1 mutated | HC & V1 non-mutated | V1 mutated & HC non-mutated | *p*-value | *p*-value (adjusted) | V1 mutation (discordant) | HC mutation (discordant) | V1-HC | Literature |
|---|---|---|---|---|---|---|---|---|---|---|
| 6569A | 2 | 0 | 56 | 3 | 1.00E+00 | 1 | 4.92% | 3.28% | 1.64% | - |
| 6755A | 0 | 0 | 60 | 1 | 1.00E+00 | 1 | 1.64% | 0.00% | 1.64% | - |
| 6776C | 0 | 1 | 59 | 1 | 1.00E+00 | 1 | 1.64% | 0.00% | 1.64% | - |
| 6852A | 0 | 0 | 60 | 1 | 1.00E+00 | 1 | 1.64% | 0.00% | 1.64% | - |
| 6872G | 0 | 0 | 60 | 1 | 1.00E+00 | 1 | 1.64% | 0.00% | 1.64% | - |
| 7051C | 0 | 0 | 60 | 1 | 1.00E+00 | 1 | 1.64% | 0.00% | 1.64% | - |
| 7094C | 0 | 0 | 60 | 1 | 1.00E+00 | 1 | 1.64% | 0.00% | 1.64% | - |
| 7109T | 0 | 0 | 60 | 1 | 1.00E+00 | 1 | 1.64% | 0.00% | 1.64% | - |
| 723C | 0 | 0 | 60 | 1 | 1.00E+00 | 1 | 1.64% | 0.00% | 1.64% | - |
| 735G | 0 | 0 | 60 | 1 | 1.00E+00 | 1 | 1.64% | 0.00% | 1.64% | - |
| 7374G | 0 | 0 | 60 | 1 | 1.00E+00 | 1 | 1.64% | 0.00% | 1.64% | - |
| 7424G | 0 | 0 | 60 | 1 | 1.00E+00 | 1 | 1.64% | 0.00% | 1.64% | - |
| 7476T | 1 | 0 | 58 | 2 | 1.00E+00 | 1 | 3.28% | 1.64% | 1.64% | - |
| 7598A | 0 | 0 | 60 | 1 | 1.00E+00 | 1 | 1.64% | 0.00% | 1.64% | - |
| 7849T | 0 | 0 | 60 | 1 | 1.00E+00 | 1 | 1.64% | 0.00% | 1.64% | - |
| 7852A | 0 | 0 | 60 | 1 | 1.00E+00 | 1 | 1.64% | 0.00% | 1.64% | - |
| 8179G | 0 | 0 | 60 | 1 | 1.00E+00 | 1 | 1.64% | 0.00% | 1.64% | - |
| 8378G | 0 | 0 | 60 | 1 | 1.00E+00 | 1 | 1.64% | 0.00% | 1.64% | - |
| 8461T | 0 | 0 | 60 | 1 | 1.00E+00 | 1 | 1.64% | 0.00% | 1.64% | - |
| 8464T | 0 | 0 | 60 | 1 | 1.00E+00 | 1 | 1.64% | 0.00% | 1.64% | - |
| 8468T | 0 | 0 | 60 | 1 | 1.00E+00 | 1 | 1.64% | 0.00% | 1.64% | - |
| 8558A | 0 | 0 | 60 | 1 | 1.00E+00 | 1 | 1.64% | 0.00% | 1.64% | - |
| 8598C | 0 | 0 | 60 | 1 | 1.00E+00 | 1 | 1.64% | 0.00% | 1.64% | - |
| 8666G | 0 | 0 | 60 | 1 | 1.00E+00 | 1 | 1.64% | 0.00% | 1.64% | - |
| 8793C | 0 | 0 | 60 | 1 | 1.00E+00 | 1 | 1.64% | 0.00% | 1.64% | - |
| 8842C | 0 | 0 | 60 | 1 | 1.00E+00 | 1 | 1.64% | 0.00% | 1.64% | - |
| 8843C | 0 | 0 | 60 | 1 | 1.00E+00 | 1 | 1.64% | 0.00% | 1.64% | - |
| 8850G | 0 | 0 | 60 | 1 | 1.00E+00 | 1 | 1.64% | 0.00% | 1.64% | - |

**Table S9 — Mutation distribution between HC and PwMS at VIS1. McNemar's test with FDR (continued)**

| Mutation | HC mutated & V1 non-mutated | HC & V1 mutated | HC & V1 non-mutated | V1 mutated & HC non-mutated | *p*-value | *p*-value (adjusted) | V1 mutation (discordant) | HC mutation (discordant) | V1-HC | Literature |
|---|---|---|---|---|---|---|---|---|---|---|
| 8865A | 0 | 0 | 60 | 1 | 1.00E+00 | 1 | 1.64% | 0.00% | 1.64% | - |
| 8881C | 0 | 0 | 60 | 1 | 1.00E+00 | 1 | 1.64% | 0.00% | 1.64% | - |
| 8911C | 0 | 0 | 60 | 1 | 1.00E+00 | 1 | 1.64% | 0.00% | 1.64% | - |
| 8961G | 0 | 0 | 60 | 1 | 1.00E+00 | 1 | 1.64% | 0.00% | 1.64% | - |
| 896G | 0 | 0 | 60 | 1 | 1.00E+00 | 1 | 1.64% | 0.00% | 1.64% | - |
| 9010A | 0 | 0 | 60 | 1 | 1.00E+00 | 1 | 1.64% | 0.00% | 1.64% | - |
| 9075T | 2 | 0 | 56 | 3 | 1.00E+00 | 1 | 4.92% | 3.28% | 1.64% | - |
| 9093G | 0 | 0 | 60 | 1 | 1.00E+00 | 1 | 1.64% | 0.00% | 1.64% | - |
| 9123T | 0 | 0 | 60 | 1 | 1.00E+00 | 1 | 1.64% | 0.00% | 1.64% | - |
| 9129T | 0 | 0 | 60 | 1 | 1.00E+00 | 1 | 1.64% | 0.00% | 1.64% | - |
| 9145A | 0 | 0 | 60 | 1 | 1.00E+00 | 1 | 1.64% | 0.00% | 1.64% | - |
| 9163A | 0 | 0 | 60 | 1 | 1.00E+00 | 1 | 1.64% | 0.00% | 1.64% | - |
| 9185C | 0 | 0 | 60 | 1 | 1.00E+00 | 1 | 1.64% | 0.00% | 1.64% | - |
| 9254G | 1 | 0 | 58 | 2 | 1.00E+00 | 1 | 3.28% | 1.64% | 1.64% | - |
| 9355G | 0 | 0 | 60 | 1 | 1.00E+00 | 1 | 1.64% | 0.00% | 1.64% | - |
| 9456G | 0 | 0 | 60 | 1 | 1.00E+00 | 1 | 1.64% | 0.00% | 1.64% | - |
| 9527T | 0 | 0 | 60 | 1 | 1.00E+00 | 1 | 1.64% | 0.00% | 1.64% | - |
| 9540C | 0 | 0 | 60 | 1 | 1.00E+00 | 1 | 1.64% | 0.00% | 1.64% | - |
| 9591A | 0 | 0 | 60 | 1 | 1.00E+00 | 1 | 1.64% | 0.00% | 1.64% | - |
| 9632G | 0 | 0 | 60 | 1 | 1.00E+00 | 1 | 1.64% | 0.00% | 1.64% | - |
| 9861C | 0 | 0 | 60 | 1 | 1.00E+00 | 1 | 1.64% | 0.00% | 1.64% | - |
| 988A | 0 | 0 | 60 | 1 | 1.00E+00 | 1 | 1.64% | 0.00% | 1.64% | - |
| 9948A | 0 | 0 | 60 | 1 | 1.00E+00 | 1 | 1.64% | 0.00% | 1.64% | - |
| 10685A | 1 | 0 | 59 | 1 | 1.00E+00 | 1 | 1.64% | 1.64% | 0.00% | - |
| 10876G | 1 | 0 | 59 | 1 | 1.00E+00 | 1 | 1.64% | 1.64% | 0.00% | - |
| 10915C | 1 | 0 | 59 | 1 | 1.00E+00 | 1 | 1.64% | 1.64% | 0.00% | - |
| 11253C | 1 | 0 | 59 | 1 | 1.00E+00 | 1 | 1.64% | 1.64% | 0.00% | - |
| 11377A | 1 | 0 | 59 | 1 | 1.00E+00 | 1 | 1.64% | 1.64% | 0.00% | - |

#### Table S9 — Mutation distribution between HC and PwMS at VIS1. McNemar's test with FDR (continued)

| Mutation | HC mutated & V1 non-mutated | HC & V1 mutated | HC & V1 non-mutated | V1 mutated & HC non-mutated | *p*-value | *p*-value (adjusted) | V1 mutation (discordant) | HC mutation (discordant) | V1-HC | Literature |
|---|---|---|---|---|---|---|---|---|---|---|
| 114T | 1 | 0 | 59 | 1 | 1.00E+00 | 1 | 1.64% | 1.64% | 0.00% | - |
| 12007A | 2 | 0 | 57 | 2 | 1.00E+00 | 1 | 3.28% | 3.28% | 0.00% | - |
| 12441C | 1 | 0 | 59 | 1 | 1.00E+00 | 1 | 1.64% | 1.64% | 0.00% | - |
| 13020C | 1 | 0 | 59 | 1 | 1.00E+00 | 1 | 1.64% | 1.64% | 0.00% | - |
| 13095C | 1 | 0 | 59 | 1 | 1.00E+00 | 1 | 1.64% | 1.64% | 0.00% | - |
| 13105G | 1 | 0 | 59 | 1 | 1.00E+00 | 1 | 1.64% | 1.64% | 0.00% | - |
| 13260C | 1 | 0 | 59 | 1 | 1.00E+00 | 1 | 1.64% | 1.64% | 0.00% | - |
| 13404C | 1 | 0 | 59 | 1 | 1.00E+00 | 1 | 1.64% | 1.64% | 0.00% | - |
| 13617C | 2 | 2 | 55 | 2 | 1.00E+00 | 1 | 3.28% | 3.28% | 0.00% | - |
| 13734C | 1 | 0 | 59 | 1 | 1.00E+00 | 1 | 1.64% | 1.64% | 0.00% | - |
| 14182C | 1 | 1 | 58 | 1 | 1.00E+00 | 1 | 1.64% | 1.64% | 0.00% | - |
| 1438G | 3 | 55 | 0 | 3 | 1.00E+00 | 1 | 4.92% | 4.92% | 0.00% | - |
| 14793G | 1 | 1 | 58 | 1 | 1.00E+00 | 1 | 1.64% | 1.64% | 0.00% | - |
| 14978G | 0 | 1 | 60 | 0 | - | | 0.00% | 0.00% | 0.00% | - |
| 15043A | 2 | 0 | 57 | 2 | 1.00E+00 | 1 | 3.28% | 3.28% | 0.00% | - |
| 15355A | 1 | 0 | 59 | 1 | 1.00E+00 | 1 | 1.64% | 1.64% | 0.00% | - |
| 15497A | 1 | 0 | 59 | 1 | 1.00E+00 | 1 | 1.64% | 1.64% | 0.00% | - |
| 15607G | 4 | 0 | 53 | 4 | 1.00E+00 | 1 | 6.56% | 6.56% | 0.00% | - |
| 15619T | 1 | 0 | 59 | 1 | 1.00E+00 | 1 | 1.64% | 1.64% | 0.00% | - |
| 15833T | 2 | 0 | 57 | 2 | 1.00E+00 | 1 | 3.28% | 3.28% | 0.00% | - |
| 15907G | 1 | 0 | 59 | 1 | 1.00E+00 | 1 | 1.64% | 1.64% | 0.00% | - |
| 15924G | 3 | 0 | 55 | 3 | 1.00E+00 | 1 | 4.92% | 4.92% | 0.00% | - |
| 15947G | 1 | 0 | 59 | 1 | 1.00E+00 | 1 | 1.64% | 1.64% | 0.00% | - |
| 16129A | 3 | 0 | 55 | 3 | 1.00E+00 | 1 | 4.92% | 4.92% | 0.00% | - |
| 16129C | 1 | 0 | 59 | 1 | 1.00E+00 | 1 | 1.64% | 1.64% | 0.00% | - |
| 16148T | 1 | 0 | 59 | 1 | 1.00E+00 | 1 | 1.64% | 1.64% | 0.00% | - |
| 16172C | 4 | 0 | 53 | 4 | 1.00E+00 | 1 | 6.56% | 6.56% | 0.00% | - |
| 16247G | 1 | 0 | 59 | 1 | 1.00E+00 | 1 | 1.64% | 1.64% | 0.00% | - |

#### Table S9 — Mutation distribution between HC and PwMS at VIS1. McNemar's test with FDR (continued)

| Mutation | HC mutated & V1 non-mutated | HC & V1 mutated | HC & V1 non-mutated | V1 mutated & HC non-mutated | *p*-value | *p*-value (adjusted) | V1 mutation (discordant) | HC mutation (discordant) | V1-HC | Literature |
|---|---|---|---|---|---|---|---|---|---|---|
| 16320T | 1 | 0 | 59 | 1 | 1.00E+00 | 1 | 1.64% | 1.64% | 0.00% | - |
| 16324C | 1 | 0 | 59 | 1 | 1.00E+00 | 1 | 1.64% | 1.64% | 0.00% | - |
| 16354T | 1 | 0 | 59 | 1 | 1.00E+00 | 1 | 1.64% | 1.64% | 0.00% | - |
| 16362C | 3 | 1 | 54 | 3 | 1.00E+00 | 1 | 4.92% | 4.92% | 0.00% | - |
| 16482G | 1 | 0 | 59 | 1 | 1.00E+00 | 1 | 1.64% | 1.64% | 0.00% | - |
| 16T | 1 | 0 | 59 | 1 | 1.00E+00 | 1 | 1.64% | 1.64% | 0.00% | - |
| 1700C | 1 | 0 | 59 | 1 | 1.00E+00 | 1 | 1.64% | 1.64% | 0.00% | - |
| 1811G | 5 | 2 | 49 | 5 | 1.00E+00 | 1 | 8.20% | 8.20% | 0.00% | - |
| 217C | 1 | 0 | 59 | 1 | 1.00E+00 | 1 | 1.64% | 1.64% | 0.00% | - |
| 319C | 1 | 0 | 59 | 1 | 1.00E+00 | 1 | 1.64% | 1.64% | 0.00% | - |
| 3333T | 1 | 0 | 59 | 1 | 1.00E+00 | 1 | 1.64% | 1.64% | 0.00% | - |
| 3720G | 1 | 0 | 59 | 1 | 1.00E+00 | 1 | 1.64% | 1.64% | 0.00% | - |
| 3915A | 1 | 0 | 59 | 1 | 1.00E+00 | 1 | 1.64% | 1.64% | 0.00% | - |
| 3990T | 1 | 0 | 59 | 1 | 1.00E+00 | 1 | 1.64% | 1.64% | 0.00% | - |
| 41T | 1 | 0 | 59 | 1 | 1.00E+00 | 1 | 1.64% | 1.64% | 0.00% | - |
| 4318T | 5 | 9 | 42 | 5 | 1.00E+00 | 1 | 8.20% | 8.20% | 0.00% | - |
| 4727G | 1 | 0 | 59 | 1 | 1.00E+00 | 1 | 1.64% | 1.64% | 0.00% | - |
| 4769G | 3 | 55 | 0 | 3 | 1.00E+00 | 1 | 4.92% | 4.92% | 0.00% | - |
| 4793G | 1 | 0 | 59 | 1 | 1.00E+00 | 1 | 1.64% | 1.64% | 0.00% | - |
| 497T | 4 | 0 | 53 | 4 | 1.00E+00 | 1 | 6.56% | 6.56% | 0.00% | - |
| 5390G | 1 | 0 | 59 | 1 | 1.00E+00 | 1 | 1.64% | 1.64% | 0.00% | - |
| 6045T | 1 | 0 | 59 | 1 | 1.00E+00 | 1 | 1.64% | 1.64% | 0.00% | - |
| 6152C | 1 | 0 | 59 | 1 | 1.00E+00 | 1 | 1.64% | 1.64% | 0.00% | - |
| 6734A | 1 | 0 | 59 | 1 | 1.00E+00 | 1 | 1.64% | 1.64% | 0.00% | - |
| 7521A | 2 | 0 | 57 | 2 | 1.00E+00 | 1 | 3.28% | 3.28% | 0.00% | - |
| 7650T | 1 | 0 | 59 | 1 | 1.00E+00 | 1 | 1.64% | 1.64% | 0.00% | - |
| 7768G | 1 | 1 | 58 | 1 | 1.00E+00 | 1 | 1.64% | 1.64% | 0.00% | - |
| 8140T | 1 | 0 | 59 | 1 | 1.00E+00 | 1 | 1.64% | 1.64% | 0.00% | - |

#### Table S9 — Mutation distribution between HC and PwMS at VIS1. McNemar's test with FDR (continued)

| Mutation | HC mutated & V1 non-mutated | HC & V1 mutated | HC & V1 non-mutated | V1 mutated & HC non-mutated | *p*-value | *p*-value (adjusted) | V1 mutation (discordant) | HC mutation (discordant) | V1-HC | Literature |
|---|---|---|---|---|---|---|---|---|---|---|
| 8277C | 1 | 0 | 59 | 1 | 1.00E+00 | 1 | 1.64% | 1.64% | 0.00% | - |
| 8616T | 1 | 0 | 59 | 1 | 1.00E+00 | 1 | 1.64% | 1.64% | 0.00% | - |
| 8706G | 1 | 0 | 59 | 1 | 1.00E+00 | 1 | 1.64% | 1.64% | 0.00% | - |
| 8943T | 2 | 1 | 56 | 2 | 1.00E+00 | 1 | 3.28% | 3.28% | 0.00% | - |
| 9060A | 1 | 0 | 59 | 1 | 1.00E+00 | 1 | 1.64% | 1.64% | 0.00% | - |
| 9196A | 1 | 0 | 59 | 1 | 1.00E+00 | 1 | 1.64% | 1.64% | 0.00% | - |
| 922A | 1 | 0 | 59 | 1 | 1.00E+00 | 1 | 1.64% | 1.64% | 0.00% | - |
| 930A | 3 | 0 | 55 | 3 | 1.00E+00 | 1 | 4.92% | 4.92% | 0.00% | - |
| 9380A | 1 | 0 | 59 | 1 | 1.00E+00 | 1 | 1.64% | 1.64% | 0.00% | - |
| 9434G | 2 | 0 | 57 | 2 | 1.00E+00 | 1 | 3.28% | 3.28% | 0.00% | - |
| 9477A | 2 | 2 | 55 | 2 | 1.00E+00 | 1 | 3.28% | 3.28% | 0.00% | - |
| 951A | 1 | 0 | 59 | 1 | 1.00E+00 | 1 | 1.64% | 1.64% | 0.00% | - |
| 9530C | 1 | 0 | 59 | 1 | 1.00E+00 | 1 | 1.64% | 1.64% | 0.00% | - |
| 9938C | 1 | 0 | 59 | 1 | 1.00E+00 | 1 | 1.64% | 1.64% | 0.00% | - |
| 9947A | 1 | 0 | 59 | 1 | 1.00E+00 | 1 | 1.64% | 1.64% | 0.00% | - |
| 10034C | 2 | 0 | 58 | 1 | 1.00E+00 | 1 | 1.64% | 3.28% | -1.64% | - |
| 10197A | 1 | 0 | 60 | 0 | 1.00E+00 | 1 | 0.00% | 1.64% | -1.64% | - |
| 10220G | 1 | 0 | 60 | 0 | 1.00E+00 | 1 | 0.00% | 1.64% | -1.64% | - |
| 10238C | 2 | 0 | 58 | 1 | 1.00E+00 | 1 | 1.64% | 3.28% | -1.64% | - |
| 10248C | 1 | 0 | 60 | 0 | 1.00E+00 | 1 | 0.00% | 1.64% | -1.64% | - |
| 10336C | 1 | 0 | 60 | 0 | 1.00E+00 | 1 | 0.00% | 1.64% | -1.64% | - |
| 10352G | 1 | 0 | 60 | 0 | 1.00E+00 | 1 | 0.00% | 1.64% | -1.64% | - |
| 10454C | 1 | 0 | 60 | 0 | 1.00E+00 | 1 | 0.00% | 1.64% | -1.64% | - |
| 10595C | 1 | 0 | 60 | 0 | 1.00E+00 | 1 | 0.00% | 1.64% | -1.64% | - |
| 10609C | 1 | 0 | 60 | 0 | 1.00E+00 | 1 | 0.00% | 1.64% | -1.64% | - |
| 10646A | 1 | 0 | 60 | 0 | 1.00E+00 | 1 | 0.00% | 1.64% | -1.64% | - |
| 10750G | 1 | 0 | 60 | 0 | 1.00E+00 | 1 | 0.00% | 1.64% | -1.64% | - |
| 10966C | 1 | 0 | 60 | 0 | 1.00E+00 | 1 | 0.00% | 1.64% | -1.64% | - |

#### Table S9 — Mutation distribution between HC and PwMS at VIS1. McNemar's test with FDR (continued)

| Mutation | HC mutated & V1 non-mutated | HC & V1 mutated | HC & V1 non-mutated | V1 mutated & HC non-mutated | *p*-value | *p*-value (adjusted) | V1 mutation (discordant) | HC mutation (discordant) | V1-HC | Literature |
|---|---|---|---|---|---|---|---|---|---|---|
| 10993A | 1 | 0 | 60 | 0 | 1.00E+00 | 1 | 0.00% | 1.64% | -1.64% | - |
| 11053G | 1 | 0 | 60 | 0 | 1.00E+00 | 1 | 0.00% | 1.64% | -1.64% | - |
| 11260C | 1 | 0 | 60 | 0 | 1.00E+00 | 1 | 0.00% | 1.64% | -1.64% | - |
| 11467G | 7 | 5 | 43 | 6 | 1.00E+00 | 1 | 9.84% | 11.48% | -1.64% | - |
| 11653G | 1 | 0 | 60 | 0 | 1.00E+00 | 1 | 0.00% | 1.64% | -1.64% | - |
| 11654G | 1 | 0 | 60 | 0 | 1.00E+00 | 1 | 0.00% | 1.64% | -1.64% | - |
| 11662C | 1 | 0 | 60 | 0 | 1.00E+00 | 1 | 0.00% | 1.64% | -1.64% | - |
| 11674T | 1 | 0 | 60 | 0 | 1.00E+00 | 1 | 0.00% | 1.64% | -1.64% | - |
| 11812G | 5 | 0 | 52 | 4 | 1.00E+00 | 1 | 6.56% | 8.20% | -1.64% | - |
| 11827C | 1 | 0 | 60 | 0 | 1.00E+00 | 1 | 0.00% | 1.64% | -1.64% | - |
| 11840T | 1 | 0 | 60 | 0 | 1.00E+00 | 1 | 0.00% | 1.64% | -1.64% | - |
| 11893G | 1 | 0 | 60 | 0 | 1.00E+00 | 1 | 0.00% | 1.64% | -1.64% | - |
| 11899C | 1 | 0 | 60 | 0 | 1.00E+00 | 1 | 0.00% | 1.64% | -1.64% | - |
| 11944C | 1 | 0 | 60 | 0 | 1.00E+00 | 1 | 0.00% | 1.64% | -1.64% | - |
| 11947G | 1 | 0 | 60 | 0 | 1.00E+00 | 1 | 0.00% | 1.64% | -1.64% | - |
| 11978A | 1 | 0 | 60 | 0 | 1.00E+00 | 1 | 0.00% | 1.64% | -1.64% | - |
| 12127A | 1 | 0 | 60 | 0 | 1.00E+00 | 1 | 0.00% | 1.64% | -1.64% | - |
| 12172G | 2 | 0 | 58 | 1 | 1.00E+00 | 1 | 1.64% | 3.28% | -1.64% | - |
| 12308G | 7 | 5 | 43 | 6 | 1.00E+00 | 1 | 9.84% | 11.48% | -1.64% | MS (risk) [62] |
| 12372A | 7 | 5 | 43 | 6 | 1.00E+00 | 1 | 9.84% | 11.48% | -1.64% | - |
| 12373G | 1 | 0 | 60 | 0 | 1.00E+00 | 1 | 0.00% | 1.64% | -1.64% | - |
| 12414C | 1 | 0 | 60 | 0 | 1.00E+00 | 1 | 0.00% | 1.64% | -1.64% | - |
| 1243C | 1 | 0 | 60 | 0 | 1.00E+00 | 1 | 0.00% | 1.64% | -1.64% | - |
| 12501A | 2 | 0 | 58 | 1 | 1.00E+00 | 1 | 1.64% | 3.28% | -1.64% | - |
| 12510T | 1 | 0 | 60 | 0 | 1.00E+00 | 1 | 0.00% | 1.64% | -1.64% | - |
| 12557T | 1 | 0 | 60 | 0 | 1.00E+00 | 1 | 0.00% | 1.64% | -1.64% | - |
| 12618A | 1 | 0 | 60 | 0 | 1.00E+00 | 1 | 0.00% | 1.64% | -1.64% | - |
| 12634G | 1 | 0 | 60 | 0 | 1.00E+00 | 1 | 0.00% | 1.64% | -1.64% | - |

#### Table S9 — Mutation distribution between HC and PwMS at VIS1. McNemar's test with FDR (continued)

| Mutation | HC mutated & V1 non-mutated | HC & V1 mutated | HC & V1 non-mutated | V1 mutated & HC non-mutated | *p*-value | *p*-value (adjusted) | V1 mutation (discordant) | HC mutation (discordant) | V1-HC | Literature |
|---|---|---|---|---|---|---|---|---|---|---|
| 12684A | 1 | 0 | 60 | 0 | 1.00E+00 | 1 | 0.00% | 1.64% | -1.64% | - |
| 12855G | 1 | 0 | 60 | 0 | 1.00E+00 | 1 | 0.00% | 1.64% | -1.64% | - |
| 12937G | 1 | 0 | 60 | 0 | 1.00E+00 | 1 | 0.00% | 1.64% | -1.64% | - |
| 13056T | 1 | 0 | 60 | 0 | 1.00E+00 | 1 | 0.00% | 1.64% | -1.64% | - |
| 13174C | 1 | 0 | 60 | 0 | 1.00E+00 | 1 | 0.00% | 1.64% | -1.64% | - |
| 13326C | 1 | 0 | 60 | 0 | 1.00E+00 | 1 | 0.00% | 1.64% | -1.64% | - |
| 13368A | 5 | 0 | 52 | 4 | 1.00E+00 | 1 | 6.56% | 8.20% | -1.64% | - |
| 13449A | 1 | 0 | 60 | 0 | 1.00E+00 | 1 | 0.00% | 1.64% | -1.64% | - |
| 13488C | 1 | 0 | 60 | 0 | 1.00E+00 | 1 | 0.00% | 1.64% | -1.64% | - |
| 13575T | 1 | 0 | 60 | 0 | 1.00E+00 | 1 | 0.00% | 1.64% | -1.64% | - |
| 13630G | 1 | 0 | 60 | 0 | 1.00E+00 | 1 | 0.00% | 1.64% | -1.64% | - |
| 13722G | 1 | 0 | 60 | 0 | 1.00E+00 | 1 | 0.00% | 1.64% | -1.64% | - |
| 13740C | 1 | 0 | 60 | 0 | 1.00E+00 | 1 | 0.00% | 1.64% | -1.64% | - |
| 13759A | 2 | 0 | 58 | 1 | 1.00E+00 | 1 | 1.64% | 3.28% | -1.64% | - |
| 13762G | 1 | 0 | 60 | 0 | 1.00E+00 | 1 | 0.00% | 1.64% | -1.64% | - |
| 13780G | 2 | 0 | 58 | 1 | 1.00E+00 | 1 | 1.64% | 3.28% | -1.64% | - |
| 13815A | 1 | 0 | 60 | 0 | 1.00E+00 | 1 | 0.00% | 1.64% | -1.64% | - |
| 14133G | 1 | 0 | 60 | 0 | 1.00E+00 | 1 | 0.00% | 1.64% | -1.64% | - |
| 14233G | 5 | 0 | 52 | 4 | 1.00E+00 | 1 | 6.56% | 8.20% | -1.64% | - |
| 14258A | 1 | 0 | 60 | 0 | 1.00E+00 | 1 | 0.00% | 1.64% | -1.64% | - |
| 14527G | 1 | 0 | 60 | 0 | 1.00E+00 | 1 | 0.00% | 1.64% | -1.64% | - |
| 14560A | 1 | 0 | 60 | 0 | 1.00E+00 | 1 | 0.00% | 1.64% | -1.64% | - |
| 1462A | 1 | 0 | 60 | 0 | 1.00E+00 | 1 | 0.00% | 1.64% | -1.64% | - |
| 14770A | 1 | 0 | 60 | 0 | 1.00E+00 | 1 | 0.00% | 1.64% | -1.64% | - |
| 14872T | 1 | 0 | 60 | 0 | 1.00E+00 | 1 | 0.00% | 1.64% | -1.64% | - |
| 14905A | 5 | 0 | 52 | 4 | 1.00E+00 | 1 | 6.56% | 8.20% | -1.64% | - |
| 14926G | 1 | 0 | 60 | 0 | 1.00E+00 | 1 | 0.00% | 1.64% | -1.64% | - |
| 151T | 1 | 0 | 60 | 0 | 1.00E+00 | 1 | 0.00% | 1.64% | -1.64% | - |

#### Table S9 — Mutation distribution between HC and PwMS at VIS1. McNemar's test with FDR (continued)

| Mutation | HC mutated & V1 non-mutated | HC & V1 mutated | HC & V1 non-mutated | V1 mutated & HC non-mutated | *p*-value | *p*-value (adjusted) | V1 mutation (discordant) | HC mutation (discordant) | V1-HC | Literature |
|---|---|---|---|---|---|---|---|---|---|---|
| 15326G | 2 | 58 | 0 | 1 | 1.00E+00 | 1 | 1.64% | 3.28% | -1.64% | - |
| 15380G | 1 | 0 | 60 | 0 | 1.00E+00 | 1 | 0.00% | 1.64% | -1.64% | - |
| 15454C | 1 | 0 | 60 | 0 | 1.00E+00 | 1 | 0.00% | 1.64% | -1.64% | - |
| 15713G | 1 | 0 | 60 | 0 | 1.00E+00 | 1 | 0.00% | 1.64% | -1.64% | - |
| 15721C | 1 | 0 | 60 | 0 | 1.00E+00 | 1 | 0.00% | 1.64% | -1.64% | - |
| 15758G | 1 | 0 | 60 | 0 | 1.00E+00 | 1 | 0.00% | 1.64% | -1.64% | - |
| 15884C | 1 | 0 | 60 | 0 | 1.00E+00 | 1 | 0.00% | 1.64% | -1.64% | - |
| 15903G | 1 | 0 | 60 | 0 | 1.00E+00 | 1 | 0.00% | 1.64% | -1.64% | - |
| 15928A | 5 | 0 | 52 | 4 | 1.00E+00 | 1 | 6.56% | 8.20% | -1.64% | MS (risk) [61] |
| 16051G | 2 | 1 | 57 | 1 | 1.00E+00 | 1 | 1.64% | 3.28% | -1.64% | - |
| 16067T | 1 | 0 | 60 | 0 | 1.00E+00 | 1 | 0.00% | 1.64% | -1.64% | - |
| 16070G | 1 | 0 | 60 | 0 | 1.00E+00 | 1 | 0.00% | 1.64% | -1.64% | - |
| 16092C | 1 | 0 | 60 | 0 | 1.00E+00 | 1 | 0.00% | 1.64% | -1.64% | - |
| 16108T | 1 | 0 | 60 | 0 | 1.00E+00 | 1 | 0.00% | 1.64% | -1.64% | - |
| 16114A | 1 | 0 | 60 | 0 | 1.00E+00 | 1 | 0.00% | 1.64% | -1.64% | - |
| 16134T | 1 | 0 | 60 | 0 | 1.00E+00 | 1 | 0.00% | 1.64% | -1.64% | - |
| 16153A | 2 | 0 | 58 | 1 | 1.00E+00 | 1 | 1.64% | 3.28% | -1.64% | - |
| 16183C | 1 | 0 | 60 | 0 | 1.00E+00 | 1 | 0.00% | 1.64% | -1.64% | - |
| 16218T | 2 | 0 | 58 | 1 | 1.00E+00 | 1 | 1.64% | 3.28% | -1.64% | - |
| 16219G | 1 | 0 | 60 | 0 | 1.00E+00 | 1 | 0.00% | 1.64% | -1.64% | - |
| 16231C | 1 | 0 | 60 | 0 | 1.00E+00 | 1 | 0.00% | 1.64% | -1.64% | - |
| 16234T | 1 | 0 | 60 | 0 | 1.00E+00 | 1 | 0.00% | 1.64% | -1.64% | - |
| 16240G | 1 | 0 | 60 | 0 | 1.00E+00 | 1 | 0.00% | 1.64% | -1.64% | - |
| 16255A | 1 | 0 | 60 | 0 | 1.00E+00 | 1 | 0.00% | 1.64% | -1.64% | - |
| 16258G | 1 | 0 | 60 | 0 | 1.00E+00 | 1 | 0.00% | 1.64% | -1.64% | - |
| 16263C | 1 | 0 | 60 | 0 | 1.00E+00 | 1 | 0.00% | 1.64% | -1.64% | - |
| 16278T | 2 | 0 | 58 | 1 | 1.00E+00 | 1 | 1.64% | 3.28% | -1.64% | - |
| 16293G | 2 | 0 | 58 | 1 | 1.00E+00 | 1 | 1.64% | 3.28% | -1.64% | - |

#### Table S9 — Mutation distribution between HC and PwMS at VIS1. McNemar's test with FDR (continued)

| Mutation | HC mutated & V1 non-mutated | HC & V1 mutated | HC & V1 non-mutated | V1 mutated & HC non-mutated | *p*-value | *p*-value (adjusted) | V1 mutation (discordant) | HC mutation (discordant) | V1-HC | Literature |
|---|---|---|---|---|---|---|---|---|---|---|
| 16295T | 1 | 0 | 60 | 0 | 1.00E+00 | 1 | 0.00% | 1.64% | -1.64% | - |
| 16342C | 1 | 0 | 60 | 0 | 1.00E+00 | 1 | 0.00% | 1.64% | -1.64% | - |
| 16356C | 3 | 1 | 55 | 2 | 1.00E+00 | 1 | 3.28% | 4.92% | -1.64% | - |
| 16361A | 1 | 0 | 60 | 0 | 1.00E+00 | 1 | 0.00% | 1.64% | -1.64% | - |
| 16391A | 2 | 0 | 58 | 1 | 1.00E+00 | 1 | 1.64% | 3.28% | -1.64% | - |
| 16400T | 1 | 0 | 60 | 0 | 1.00E+00 | 1 | 0.00% | 1.64% | -1.64% | - |
| 16463G | 1 | 0 | 60 | 0 | 1.00E+00 | 1 | 0.00% | 1.64% | -1.64% | - |
| 16465T | 1 | 0 | 60 | 0 | 1.00E+00 | 1 | 0.00% | 1.64% | -1.64% | - |
| 1711T | 1 | 0 | 60 | 0 | 1.00E+00 | 1 | 0.00% | 1.64% | -1.64% | - |
| 1737G | 1 | 0 | 60 | 0 | 1.00E+00 | 1 | 0.00% | 1.64% | -1.64% | - |
| 1850C | 1 | 0 | 60 | 0 | 1.00E+00 | 1 | 0.00% | 1.64% | -1.64% | - |
| 1888A | 5 | 0 | 52 | 4 | 1.00E+00 | 1 | 6.56% | 8.20% | -1.64% | - |
| 189G | 2 | 0 | 58 | 1 | 1.00E+00 | 1 | 1.64% | 3.28% | -1.64% | - |
| 1900G | 1 | 0 | 60 | 0 | 1.00E+00 | 1 | 0.00% | 1.64% | -1.64% | - |
| 1901T | 1 | 0 | 60 | 0 | 1.00E+00 | 1 | 0.00% | 1.64% | -1.64% | - |
| 194T | 2 | 0 | 58 | 1 | 1.00E+00 | 1 | 1.64% | 3.28% | -1.64% | - |
| 2065G | 1 | 0 | 60 | 0 | 1.00E+00 | 1 | 0.00% | 1.64% | -1.64% | - |
| 2075C | 1 | 0 | 60 | 0 | 1.00E+00 | 1 | 0.00% | 1.64% | -1.64% | - |
| 215G | 1 | 0 | 60 | 0 | 1.00E+00 | 1 | 0.00% | 1.64% | -1.64% | - |
| 2259T | 1 | 0 | 60 | 0 | 1.00E+00 | 1 | 0.00% | 1.64% | -1.64% | - |
| 225A | 1 | 0 | 60 | 0 | 1.00E+00 | 1 | 0.00% | 1.64% | -1.64% | - |
| 227G | 1 | 0 | 60 | 0 | 1.00E+00 | 1 | 0.00% | 1.64% | -1.64% | - |
| 2294G | 1 | 0 | 60 | 0 | 1.00E+00 | 1 | 0.00% | 1.64% | -1.64% | - |
| 2352C | 1 | 0 | 60 | 0 | 1.00E+00 | 1 | 0.00% | 1.64% | -1.64% | - |
| 235G | 1 | 0 | 60 | 0 | 1.00E+00 | 1 | 0.00% | 1.64% | -1.64% | - |
| 239C | 2 | 0 | 58 | 1 | 1.00E+00 | 1 | 1.64% | 3.28% | -1.64% | - |
| 250C | 2 | 0 | 58 | 1 | 1.00E+00 | 1 | 1.64% | 3.28% | -1.64% | - |
| 2765G | 1 | 0 | 60 | 0 | 1.00E+00 | 1 | 0.00% | 1.64% | -1.64% | - |

#### Table S9 — Mutation distribution between HC and PwMS at VIS1. McNemar's test with FDR (continued)

| Mutation | HC mutated & V1 non-mutated | HC & V1 mutated | HC & V1 non-mutated | V1 mutated & HC non-mutated | *p*-value | *p*-value (adjusted) | V1 mutation (discordant) | HC mutation (discordant) | V1-HC | Literature |
|---|---|---|---|---|---|---|---|---|---|---|
| 2783G | 1 | 0 | 60 | 0 | 1.00E+00 | 1 | 0.00% | 1.64% | -1.64% | - |
| 3094A | 1 | 0 | 60 | 0 | 1.00E+00 | 1 | 0.00% | 1.64% | -1.64% | - |
| 3394C | 1 | 0 | 60 | 0 | 1.00E+00 | 1 | 0.00% | 1.64% | -1.64% | - |
| 3447G | 2 | 0 | 58 | 1 | 1.00E+00 | 1 | 1.64% | 3.28% | -1.64% | - |
| 3505G | 1 | 0 | 60 | 0 | 1.00E+00 | 1 | 0.00% | 1.64% | -1.64% | - |
| 3525A | 1 | 0 | 60 | 0 | 1.00E+00 | 1 | 0.00% | 1.64% | -1.64% | - |
| 3552C | 1 | 0 | 60 | 0 | 1.00E+00 | 1 | 0.00% | 1.64% | -1.64% | - |
| 3561G | 1 | 0 | 60 | 0 | 1.00E+00 | 1 | 0.00% | 1.64% | -1.64% | - |
| 3753C | 1 | 0 | 60 | 0 | 1.00E+00 | 1 | 0.00% | 1.64% | -1.64% | - |
| 3777C | 1 | 0 | 60 | 0 | 1.00E+00 | 1 | 0.00% | 1.64% | -1.64% | - |
| 3849A | 1 | 0 | 60 | 0 | 1.00E+00 | 1 | 0.00% | 1.64% | -1.64% | - |
| 3861G | 1 | 0 | 60 | 0 | 1.00E+00 | 1 | 0.00% | 1.64% | -1.64% | - |
| 4025T | 1 | 0 | 60 | 0 | 1.00E+00 | 1 | 0.00% | 1.64% | -1.64% | - |
| 4122G | 1 | 0 | 60 | 0 | 1.00E+00 | 1 | 0.00% | 1.64% | -1.64% | - |
| 4227G | 1 | 0 | 60 | 0 | 1.00E+00 | 1 | 0.00% | 1.64% | -1.64% | - |
| 4336C | 3 | 0 | 56 | 2 | 1.00E+00 | 1 | 3.28% | 4.92% | -1.64% | - |
| 4456T | 1 | 0 | 60 | 0 | 1.00E+00 | 1 | 0.00% | 1.64% | -1.64% | - |
| 4490T | 1 | 0 | 60 | 0 | 1.00E+00 | 1 | 0.00% | 1.64% | -1.64% | - |
| 4529T | 2 | 0 | 58 | 1 | 1.00E+00 | 1 | 1.64% | 3.28% | -1.64% | - |
| 4553C | 1 | 0 | 60 | 0 | 1.00E+00 | 1 | 0.00% | 1.64% | -1.64% | - |
| 4639C | 1 | 0 | 60 | 0 | 1.00E+00 | 1 | 0.00% | 1.64% | -1.64% | - |
| 464G | 1 | 0 | 60 | 0 | 1.00E+00 | 1 | 0.00% | 1.64% | -1.64% | - |
| 471C | 1 | 0 | 60 | 0 | 1.00E+00 | 1 | 0.00% | 1.64% | -1.64% | - |
| 4736C | 1 | 0 | 60 | 0 | 1.00E+00 | 1 | 0.00% | 1.64% | -1.64% | - |
| 482C | 1 | 0 | 60 | 0 | 1.00E+00 | 1 | 0.00% | 1.64% | -1.64% | - |
| 4859C | 1 | 0 | 60 | 0 | 1.00E+00 | 1 | 0.00% | 1.64% | -1.64% | - |
| 4917G | 5 | 0 | 52 | 4 | 1.00E+00 | 1 | 6.56% | 8.20% | -1.64% | MS (risk) [58,60,61] |
| 4924A | 1 | 0 | 60 | 0 | 1.00E+00 | 1 | 0.00% | 1.64% | -1.64% | - |

#### Table S9 — Mutation distribution between HC and PwMS at VIS1. McNemar's test with FDR (continued)

| Mutation | HC mutated & V1 non-mutated | HC & V1 mutated | HC & V1 non-mutated | V1 mutated & HC non-mutated | *p*-value | *p*-value (adjusted) | V1 mutation (discordant) | HC mutation (discordant) | V1-HC | Literature |
|---|---|---|---|---|---|---|---|---|---|---|
| 5046A | 1 | 0 | 60 | 0 | 1.00E+00 | 1 | 0.00% | 1.64% | -1.64% | - |
| 5408G | 1 | 0 | 60 | 0 | 1.00E+00 | 1 | 0.00% | 1.64% | -1.64% | - |
| 5471A | 1 | 0 | 60 | 0 | 1.00E+00 | 1 | 0.00% | 1.64% | -1.64% | - |
| 5477T | 1 | 0 | 60 | 0 | 1.00E+00 | 1 | 0.00% | 1.64% | -1.64% | - |
| 5747G | 1 | 0 | 60 | 0 | 1.00E+00 | 1 | 0.00% | 1.64% | -1.64% | - |
| 5772A | 1 | 0 | 60 | 0 | 1.00E+00 | 1 | 0.00% | 1.64% | -1.64% | - |
| 5839T | 1 | 0 | 60 | 0 | 1.00E+00 | 1 | 0.00% | 1.64% | -1.64% | - |
| 6040G | 1 | 0 | 60 | 0 | 1.00E+00 | 1 | 0.00% | 1.64% | -1.64% | - |
| 6249A | 1 | 0 | 60 | 0 | 1.00E+00 | 1 | 0.00% | 1.64% | -1.64% | - |
| 6260A | 1 | 0 | 60 | 0 | 1.00E+00 | 1 | 0.00% | 1.64% | -1.64% | - |
| 6278C | 1 | 0 | 60 | 0 | 1.00E+00 | 1 | 0.00% | 1.64% | -1.64% | - |
| 6406C | 1 | 0 | 60 | 0 | 1.00E+00 | 1 | 0.00% | 1.64% | -1.64% | - |
| 6446A | 1 | 0 | 60 | 0 | 1.00E+00 | 1 | 0.00% | 1.64% | -1.64% | - |
| 6620C | 1 | 0 | 60 | 0 | 1.00E+00 | 1 | 0.00% | 1.64% | -1.64% | - |
| 6951A | 1 | 0 | 60 | 0 | 1.00E+00 | 1 | 0.00% | 1.64% | -1.64% | - |
| 709A | 6 | 1 | 49 | 5 | 1.00E+00 | 1 | 8.20% | 9.84% | -1.64% | - |
| 7146G | 1 | 0 | 60 | 0 | 1.00E+00 | 1 | 0.00% | 1.64% | -1.64% | - |
| 7184G | 1 | 0 | 60 | 0 | 1.00E+00 | 1 | 0.00% | 1.64% | -1.64% | - |
| 7226A | 1 | 0 | 60 | 0 | 1.00E+00 | 1 | 0.00% | 1.64% | -1.64% | - |
| 7256T | 1 | 0 | 60 | 0 | 1.00E+00 | 1 | 0.00% | 1.64% | -1.64% | - |
| 7269A | 1 | 0 | 60 | 0 | 1.00E+00 | 1 | 0.00% | 1.64% | -1.64% | - |
| 7278C | 1 | 0 | 60 | 0 | 1.00E+00 | 1 | 0.00% | 1.64% | -1.64% | - |
| 7295G | 1 | 0 | 60 | 0 | 1.00E+00 | 1 | 0.00% | 1.64% | -1.64% | - |
| 7316A | 1 | 0 | 60 | 0 | 1.00E+00 | 1 | 0.00% | 1.64% | -1.64% | - |
| 7340A | 1 | 0 | 60 | 0 | 1.00E+00 | 1 | 0.00% | 1.64% | -1.64% | - |
| 73G | 17 | 13 | 15 | 16 | 1.00E+00 | 1 | 26.23% | 27.87% | -1.64% | - |
| 7444A | 1 | 0 | 60 | 0 | 1.00E+00 | 1 | 0.00% | 1.64% | -1.64% | - |
| 7468T | 1 | 0 | 60 | 0 | 1.00E+00 | 1 | 0.00% | 1.64% | -1.64% | - |

**Table S9 — Mutation distribution between HC and PwMS at VIS1. McNemar's test with FDR (continued)**

| Mutation | HC mutated & V1 non-mutated | HC & V1 mutated | HC & V1 non-mutated | V1 mutated & HC non-mutated | *p*-value | *p*-value (adjusted) | V1 mutation (discordant) | HC mutation (discordant) | V1-HC | Literature |
|---|---|---|---|---|---|---|---|---|---|---|
| 750G | 2 | 58 | 0 | 1 | 1.00E+00 | 1 | 1.64% | 3.28% | -1.64% | - |
| 7691C | 1 | 0 | 60 | 0 | 1.00E+00 | 1 | 0.00% | 1.64% | -1.64% | - |
| 7705C | 1 | 0 | 60 | 0 | 1.00E+00 | 1 | 0.00% | 1.64% | -1.64% | - |
| 770T | 1 | 0 | 60 | 0 | 1.00E+00 | 1 | 0.00% | 1.64% | -1.64% | - |
| 7729G | 1 | 0 | 60 | 0 | 1.00E+00 | 1 | 0.00% | 1.64% | -1.64% | - |
| 7789A | 1 | 0 | 60 | 0 | 1.00E+00 | 1 | 0.00% | 1.64% | -1.64% | - |
| 7853A | 1 | 0 | 60 | 0 | 1.00E+00 | 1 | 0.00% | 1.64% | -1.64% | - |
| 7864T | 1 | 0 | 60 | 0 | 1.00E+00 | 1 | 0.00% | 1.64% | -1.64% | - |
| 8014T | 1 | 0 | 60 | 0 | 1.00E+00 | 1 | 0.00% | 1.64% | -1.64% | - |
| 8098G | 1 | 0 | 60 | 0 | 1.00E+00 | 1 | 0.00% | 1.64% | -1.64% | - |
| 8167C | 2 | 0 | 58 | 1 | 1.00E+00 | 1 | 1.64% | 3.28% | -1.64% | - |
| 8242C | 1 | 0 | 60 | 0 | 1.00E+00 | 1 | 0.00% | 1.64% | -1.64% | - |
| 8322C | 1 | 0 | 60 | 0 | 1.00E+00 | 1 | 0.00% | 1.64% | -1.64% | - |
| 8393T | 1 | 0 | 60 | 0 | 1.00E+00 | 1 | 0.00% | 1.64% | -1.64% | - |
| 8410T | 1 | 0 | 60 | 0 | 1.00E+00 | 1 | 0.00% | 1.64% | -1.64% | - |
| 8448C | 1 | 0 | 60 | 0 | 1.00E+00 | 1 | 0.00% | 1.64% | -1.64% | - |
| 8473C | 1 | 0 | 60 | 0 | 1.00E+00 | 1 | 0.00% | 1.64% | -1.64% | - |
| 8477G | 1 | 0 | 60 | 0 | 1.00E+00 | 1 | 0.00% | 1.64% | -1.64% | - |
| 8578T | 1 | 0 | 60 | 0 | 1.00E+00 | 1 | 0.00% | 1.64% | -1.64% | - |
| 8639C | 1 | 0 | 60 | 0 | 1.00E+00 | 1 | 0.00% | 1.64% | -1.64% | - |
| 8659G | 1 | 0 | 60 | 0 | 1.00E+00 | 1 | 0.00% | 1.64% | -1.64% | - |
| 8697A | 5 | 0 | 52 | 4 | 1.00E+00 | 1 | 6.56% | 8.20% | -1.64% | - |
| 8705C | 1 | 0 | 60 | 0 | 1.00E+00 | 1 | 0.00% | 1.64% | -1.64% | - |
| 8840T | 1 | 0 | 60 | 0 | 1.00E+00 | 1 | 0.00% | 1.64% | -1.64% | - |
| 8860G | 2 | 58 | 0 | 1 | 1.00E+00 | 1 | 1.64% | 3.28% | -1.64% | - |
| 8869G | 1 | 0 | 60 | 0 | 1.00E+00 | 1 | 0.00% | 1.64% | -1.64% | - |
| 8887G | 1 | 0 | 60 | 0 | 1.00E+00 | 1 | 0.00% | 1.64% | -1.64% | - |
| 8938G | 1 | 0 | 60 | 0 | 1.00E+00 | 1 | 0.00% | 1.64% | -1.64% | - |

#### Table S9 — Mutation distribution between HC and PwMS at VIS1. McNemar's test with FDR (continued)

| Mutation | HC mutated & V1 non-mutated | HC & V1 mutated | HC & V1 non-mutated | V1 mutated & HC non-mutated | *p*-value | *p*-value (adjusted) | V1 mutation (discordant) | HC mutation (discordant) | V1-HC | Literature |
|---|---|---|---|---|---|---|---|---|---|---|
| 8982G | 1 | 0 | 60 | 0 | 1.00E+00 | 1 | 0.00% | 1.64% | -1.64% | - |
| 8994A | 1 | 0 | 60 | 0 | 1.00E+00 | 1 | 0.00% | 1.64% | -1.64% | - |
| 9150G | 1 | 0 | 60 | 0 | 1.00E+00 | 1 | 0.00% | 1.64% | -1.64% | - |
| 9151G | 1 | 0 | 60 | 0 | 1.00E+00 | 1 | 0.00% | 1.64% | -1.64% | - |
| 9157A | 1 | 0 | 60 | 0 | 1.00E+00 | 1 | 0.00% | 1.64% | -1.64% | - |
| 9300A | 1 | 0 | 60 | 0 | 1.00E+00 | 1 | 0.00% | 1.64% | -1.64% | - |
| 9336C | 1 | 0 | 60 | 0 | 1.00E+00 | 1 | 0.00% | 1.64% | -1.64% | - |
| 9371T | 1 | 0 | 60 | 0 | 1.00E+00 | 1 | 0.00% | 1.64% | -1.64% | - |
| 93G | 1 | 0 | 60 | 0 | 1.00E+00 | 1 | 0.00% | 1.64% | -1.64% | - |
| 9617G | 1 | 0 | 60 | 0 | 1.00E+00 | 1 | 0.00% | 1.64% | -1.64% | - |
| 961C | 1 | 0 | 60 | 0 | 1.00E+00 | 1 | 0.00% | 1.64% | -1.64% | - |
| 961G | 1 | 0 | 60 | 0 | 1.00E+00 | 1 | 0.00% | 1.64% | -1.64% | - |
| 9800C | 1 | 0 | 60 | 0 | 1.00E+00 | 1 | 0.00% | 1.64% | -1.64% | - |
| 10463C | 6 | 0 | 51 | 4 | 7.52E-01 | 1 | 6.56% | 9.84% | -3.28% | - |
| 10499G | 2 | 0 | 59 | 0 | 4.80E-01 | 1 | 0.00% | 3.28% | -3.28% | - |
| 11332T | 2 | 0 | 59 | 0 | 4.80E-01 | 1 | 0.00% | 3.28% | -3.28% | - |
| 11620G | 2 | 0 | 59 | 0 | 4.80E-01 | 1 | 0.00% | 3.28% | -3.28% | - |
| 11914A | 4 | 0 | 55 | 2 | 6.83E-01 | 1 | 3.28% | 6.56% | -3.28% | - |
| 13966G | 2 | 0 | 59 | 0 | 4.80E-01 | 1 | 0.00% | 3.28% | -3.28% | MS (risk) [54] |
| 14470C | 2 | 0 | 59 | 0 | 4.80E-01 | 1 | 0.00% | 3.28% | -3.28% | - |
| 15098G | 2 | 0 | 59 | 0 | 4.80E-01 | 1 | 0.00% | 3.28% | -3.28% | - |
| 15244G | 2 | 0 | 59 | 0 | 4.80E-01 | 1 | 0.00% | 3.28% | -3.28% | - |
| 152C | 9 | 2 | 43 | 7 | 8.03E-01 | 1 | 11.48% | 14.75% | -3.28% | - |
| 153G | 2 | 0 | 59 | 0 | 4.80E-01 | 1 | 0.00% | 3.28% | -3.28% | - |
| 15693C | 2 | 0 | 59 | 0 | 4.80E-01 | 1 | 0.00% | 3.28% | -3.28% | - |
| 15773A | 2 | 0 | 59 | 0 | 4.80E-01 | 1 | 0.00% | 3.28% | -3.28% | - |
| 15927A | 2 | 0 | 59 | 0 | 4.80E-01 | 1 | 0.00% | 3.28% | -3.28% | MS (risk) [61] |
| 16080G | 2 | 0 | 59 | 0 | 4.80E-01 | 1 | 0.00% | 3.28% | -3.28% | - |

**Table S9 — Mutation distribution between HC and PwMS at VIS1. McNemar's test with FDR (continued)**

| Mutation | HC mutated & V1 non-mutated | HC & V1 mutated | HC & V1 non-mutated | V1 mutated & HC non-mutated | *p*-value | *p*-value (adjusted) | V1 mutation (discordant) | HC mutation (discordant) | V1-HC | Literature |
|---|---|---|---|---|---|---|---|---|---|---|
| 16086C | 2 | 0 | 59 | 0 | 4.80E-01 | 1 | 0.00% | 3.28% | -3.28% | - |
| 16162G | 3 | 1 | 56 | 1 | 6.17E-01 | 1 | 1.64% | 4.92% | -3.28% | - |
| 16184T | 2 | 0 | 59 | 0 | 4.80E-01 | 1 | 0.00% | 3.28% | -3.28% | - |
| 16207G | 2 | 0 | 59 | 0 | 4.80E-01 | 1 | 0.00% | 3.28% | -3.28% | - |
| 16209C | 4 | 0 | 55 | 2 | 6.83E-01 | 1 | 3.28% | 6.56% | -3.28% | - |
| 16261T | 3 | 0 | 57 | 1 | 6.17E-01 | 1 | 1.64% | 4.92% | -3.28% | - |
| 16270T | 3 | 2 | 55 | 1 | 6.17E-01 | 1 | 1.64% | 4.92% | -3.28% | - |
| 16292T | 2 | 0 | 59 | 0 | 4.80E-01 | 1 | 0.00% | 3.28% | -3.28% | - |
| 16294T | 6 | 0 | 51 | 4 | 7.52E-01 | 1 | 6.56% | 9.84% | -3.28% | - |
| 16355T | 3 | 0 | 57 | 1 | 6.17E-01 | 1 | 1.64% | 4.92% | -3.28% | - |
| 207A | 2 | 0 | 59 | 0 | 4.80E-01 | 1 | 0.00% | 3.28% | -3.28% | - |
| 2706G | 18 | 14 | 13 | 16 | 8.64E-01 | 1 | 26.23% | 29.51% | -3.28% | - |
| 3184T | 2 | 0 | 59 | 0 | 4.80E-01 | 1 | 0.00% | 3.28% | -3.28% | - |
| 3197C | 4 | 2 | 53 | 2 | 6.83E-01 | 1 | 3.28% | 6.56% | -3.28% | - |
| 3357A | 2 | 0 | 59 | 0 | 4.80E-01 | 1 | 0.00% | 3.28% | -3.28% | - |
| 449C | 2 | 0 | 59 | 0 | 4.80E-01 | 1 | 0.00% | 3.28% | -3.28% | - |
| 4646C | 2 | 0 | 59 | 0 | 4.80E-01 | 1 | 0.00% | 3.28% | -3.28% | - |
| 499A | 2 | 0 | 59 | 0 | 4.80E-01 | 1 | 0.00% | 3.28% | -3.28% | - |
| 513A | 2 | 0 | 59 | 0 | 4.80E-01 | 1 | 0.00% | 3.28% | -3.28% | - |
| 5147A | 5 | 1 | 52 | 3 | 7.24E-01 | 1 | 4.92% | 8.20% | -3.28% | - |
| 5773A | 2 | 0 | 59 | 0 | 4.80E-01 | 1 | 0.00% | 3.28% | -3.28% | - |
| 5999C | 2 | 0 | 59 | 0 | 4.80E-01 | 1 | 0.00% | 3.28% | -3.28% | - |
| 6047G | 2 | 0 | 59 | 0 | 4.80E-01 | 1 | 0.00% | 3.28% | -3.28% | - |
| 6221C | 2 | 0 | 59 | 0 | 4.80E-01 | 1 | 0.00% | 3.28% | -3.28% | - |
| 6365C | 3 | 0 | 57 | 1 | 6.17E-01 | 1 | 1.64% | 4.92% | -3.28% | - |
| 6371T | 2 | 0 | 59 | 0 | 4.80E-01 | 1 | 0.00% | 3.28% | -3.28% | - |
| 6542T | 2 | 0 | 59 | 0 | 4.80E-01 | 1 | 0.00% | 3.28% | -3.28% | - |
| 7028T | 18 | 14 | 13 | 16 | 8.64E-01 | 1 | 26.23% | 29.51% | -3.28% | - |

**Table S9 — Mutation distribution between HC and PwMS at VIS1. McNemar's test with FDR (continued)**

| Mutation | HC mutated & V1 non-mutated | HC & V1 mutated | HC & V1 non-mutated | V1 mutated & HC non-mutated | *p*-value | *p*-value (adjusted) | V1 mutation (discordant) | HC mutation (discordant) | V1-HC | Literature |
|---|---|---|---|---|---|---|---|---|---|---|
| 72C | 3 | 0 | 57 | 1 | 6.17E-01 | 1 | 1.64% | 4.92% | -3.28% | - |
| 8818T | 2 | 0 | 59 | 0 | 4.80E-01 | 1 | 0.00% | 3.28% | -3.28% | - |
| 9548A | 2 | 0 | 59 | 0 | 4.80E-01 | 1 | 0.00% | 3.28% | -3.28% | - |
| 9908T | 2 | 0 | 59 | 0 | 4.80E-01 | 1 | 0.00% | 3.28% | -3.28% | - |
| 14620T | 3 | 0 | 58 | 0 | 2.48E-01 | 1 | 0.00% | 4.92% | -4.92% | - |
| 150T | 6 | 2 | 50 | 3 | 5.05E-01 | 1 | 4.92% | 9.84% | -4.92% | - |
| 15904T | 4 | 0 | 56 | 1 | 3.71E-01 | 1 | 1.64% | 6.56% | -4.92% | - |
| 16192T | 5 | 0 | 54 | 2 | 4.50E-01 | 1 | 3.28% | 8.20% | -4.92% | - |
| 16298C | 5 | 0 | 54 | 2 | 4.50E-01 | 1 | 3.28% | 8.20% | -4.92% | - |
| 16526A | 4 | 0 | 56 | 1 | 3.71E-01 | 1 | 1.64% | 6.56% | -4.92% | - |
| 204C | 3 | 0 | 58 | 0 | 2.48E-01 | 1 | 0.00% | 4.92% | -4.92% | - |
| 4580A | 4 | 0 | 56 | 1 | 3.71E-01 | 1 | 1.64% | 6.56% | -4.92% | - |
| 16223T | 5 | 0 | 55 | 1 | 2.21E-01 | 1 | 1.64% | 8.20% | -6.56% | - |
| 16304C | 9 | 0 | 47 | 5 | 4.23E-01 | 1 | 8.20% | 14.75% | -6.56% | - |
| 195C | 9 | 0 | 47 | 5 | 4.23E-01 | 1 | 8.20% | 14.75% | -6.56% | - |
| 200G | 4 | 0 | 57 | 0 | 1.34E-01 | 1 | 0.00% | 6.56% | -6.56% | - |
| 456T | 6 | 0 | 53 | 2 | 2.89E-01 | 1 | 3.28% | 9.84% | -6.56% | - |
| 5320T | 5 | 0 | 55 | 1 | 2.21E-01 | 1 | 1.64% | 8.20% | -6.56% | - |
| 12705T | 6 | 0 | 54 | 1 | 1.31E-01 | 1 | 1.64% | 9.84% | -8.20% | - |
| 16189C | 9 | 1 | 47 | 4 | 2.67E-01 | 1 | 6.56% | 14.75% | -8.20% | - |
| 1719A | 6 | 0 | 54 | 1 | 1.31E-01 | 1 | 1.64% | 9.84% | -8.20% | - |
| 6366A | 7 | 0 | 52 | 2 | 1.82E-01 | 1 | 3.28% | 11.48% | -8.20% | - |
| 6383A | 7 | 0 | 52 | 2 | 1.82E-01 | 1 | 3.28% | 11.48% | -8.20% | - |
| 8251A | 7 | 0 | 53 | 1 | 7.71E-02 | 1 | 1.64% | 11.48% | -9.84% | - |

Abbreviations: FDR — false discovery rate; HC — healthy control; LHON — Leber's hereditary optic neuropathy; MS — Multiple Sclerosis; NARP — neuropathy, ataxia, and retinitis pigmentosa; PwMS — patients with Clinically Isolated Syndrome/Relapsing-Remitting Multiple Sclerosis; V1 — patient with Clinically Isolated Syndrome/Relapsing-Remitting Multiple Sclerosis at visit 1; VIS1 — visit 1.

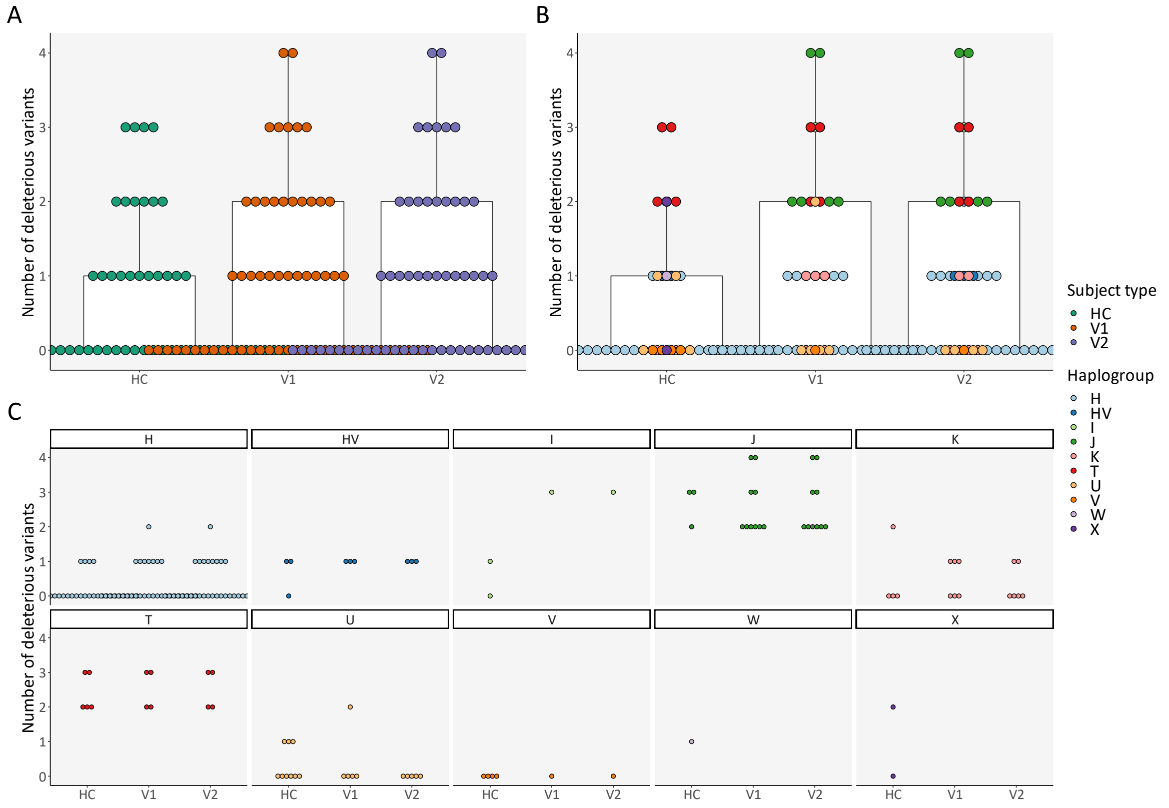

**Figure S12 — Number of deleterious variants: Cross-sectional comparison (subject types and haplogroups)**

(**A**) Number of deleterious variants, per subject type; (**B**) Number of deleterious variants, per subject type and haplogroup; (**C**) Expansion of **Figure S12B** for better visualization. Abbreviations: HC — healthy control; V1 — patient with Clinically Isolated Syndrome/Relapsing-Remitting Multiple Sclerosis at visit 1; V2 — patient with Clinically Isolated Syndrome/Relapsing-Remitting Multiple Sclerosis at visit 2.

**Table S10 — Mean number of deleterious mutations per haplogroup (simplified). Dunn test, after a Kruskal-Wallis test, with FDR**

| Haplogroups | Mean number of deleterious mutations | Adjusted *p*-value |
|---|---|---|
| J ~ K | 2.62 ~ 0.50 | 2.62E-04 |
| J ~ T | 2.62 ~ 2.44 | 9.37E-01 |
| J ~ U | 2.62 ~ 0.36 | 1.11E-05 |
| J ~ V | 2.62 ~ 0.00 | 5.63E-05 |
| T ~ U | 2.44 ~ 0.36 | 9.65E-05 |
| T ~ V | 2.44 ~ 0.00 | 1.55E-04 |
| I ~ J | 1.33 ~ 2.62 | 2.08E-01 |
| I ~ K | 1.33 ~ 0.50 | 3.89E-01 |
| I ~ T | 1.33 ~ 2.44 | 2.44E-01 |
| I ~ U | 1.33 ~ 0.36 | 2.65E-01 |
| I ~ V | 1.33 ~ 0.00 | 1.39E-01 |
| HV ~ I | 0.83 ~ 1.33 | 9.37E-01 |
| HV ~ J | 0.83 ~ 2.62 | 7.91E-02 |
| HV ~ K | 0.83 ~ 0.50 | 3.47E-01 |
| HV ~ T | 0.83 ~ 2.44 | 1.09E-01 |
| HV ~ U | 0.83 ~ 0.36 | 1.99E-01 |
| HV ~ V | 0.83 ~ 0.00 | 9.29E-02 |
| K ~ T | 0.50 ~ 2.44 | 1.20E-03 |
| K ~ U | 0.50 ~ 0.36 | 6.93E-01 |
| K ~ V | 0.50 ~ 0.00 | 3.26E-01 |
| U ~ V | 0.36 ~ 0.00 | 4.36E-01 |
| H ~ HV | 0.22 ~ 0.83 | 7.91E-02 |
| H ~ I | 0.22 ~ 1.33 | 1.63E-01 |
| H ~ J | 0.22 ~ 2.62 | 9.77E-10 |
| H ~ K | 0.22 ~ 0.50 | 3.89E-01 |
| H ~ T | 0.22 ~ 2.44 | 3.46E-07 |
| H ~ U | 0.22 ~ 0.36 | 6.64E-01 |
| H ~ V | 0.22 ~ 0.00 | 5.75E-01 |

Significant *p*-values are highlighted. Abbreviations: FDR — false discovery rate.

#### Table S11 — Mean cumulative deleterious burden per haplogroup (simplified). Dunn test, after a Kruskal-Wallis test, with FDR

| Haplogroups | Mean cumulative deleterious burden | Adjusted *p*-value |
|---|---|---|
| J ~ K | 3.92 ~ 2.60 | 2.47E-01 |
| J ~ T | 3.92 ~ 3.06 | 4.99E-01 |
| J ~ U | 3.92 ~ 2.00 | 1.32E-02 |
| J ~ V | 3.92 ~ 0.90 | 9.92E-06 |
| T ~ U | 3.06 ~ 2.00 | 1.42E-01 |
| T ~ V | 3.06 ~ 0.90 | 3.74E-04 |
| K ~ T | 2.60 ~ 3.06 | 6.13E-01 |
| K ~ U | 2.60 ~ 2.00 | 3.23E-01 |
| K ~ V | 2.60 ~ 0.90 | 1.50E-03 |
| I ~ J | 2.50 ~ 3.92 | 3.95E-01 |
| I ~ K | 2.50 ~ 2.60 | 9.23E-01 |
| I ~ T | 2.50 ~ 3.06 | 6.49E-01 |
| I ~ U | 2.50 ~ 2.00 | 5.56E-01 |
| I ~ V | 2.50 ~ 0.90 | 2.37E-02 |
| U ~ V | 2.00 ~ 0.90 | 1.37E-02 |
| HV ~ I | 1.41 ~ 2.50 | 3.00E-01 |
| HV ~ J | 1.41 ~ 3.92 | 5.22E-03 |
| HV ~ K | 1.41 ~ 2.60 | 1.25E-01 |
| HV ~ T | 1.41 ~ 3.06 | 4.35E-02 |
| HV ~ U | 1.41 ~ 2.00 | 4.32E-01 |
| HV ~ V | 1.41 ~ 0.90 | 1.73E-01 |
| H ~ HV | 1.01 ~ 1.41 | 2.61E-01 |
| H ~ I | 1.01 ~ 2.50 | 2.76E-02 |
| H ~ J | 1.01 ~ 3.92 | 5.12E-11 |
| H ~ K | 1.01 ~ 2.60 | 3.87E-05 |
| H ~ T | 1.01 ~ 3.06 | 5.48E-06 |
| H ~ U | 1.01 ~ 2.00 | 1.50E-03 |
| H ~ V | 1.01 ~ 0.90 | 4.63E-01 |

Significant *p*-values are highlighted. Abbreviations: FDR — false discovery rate.

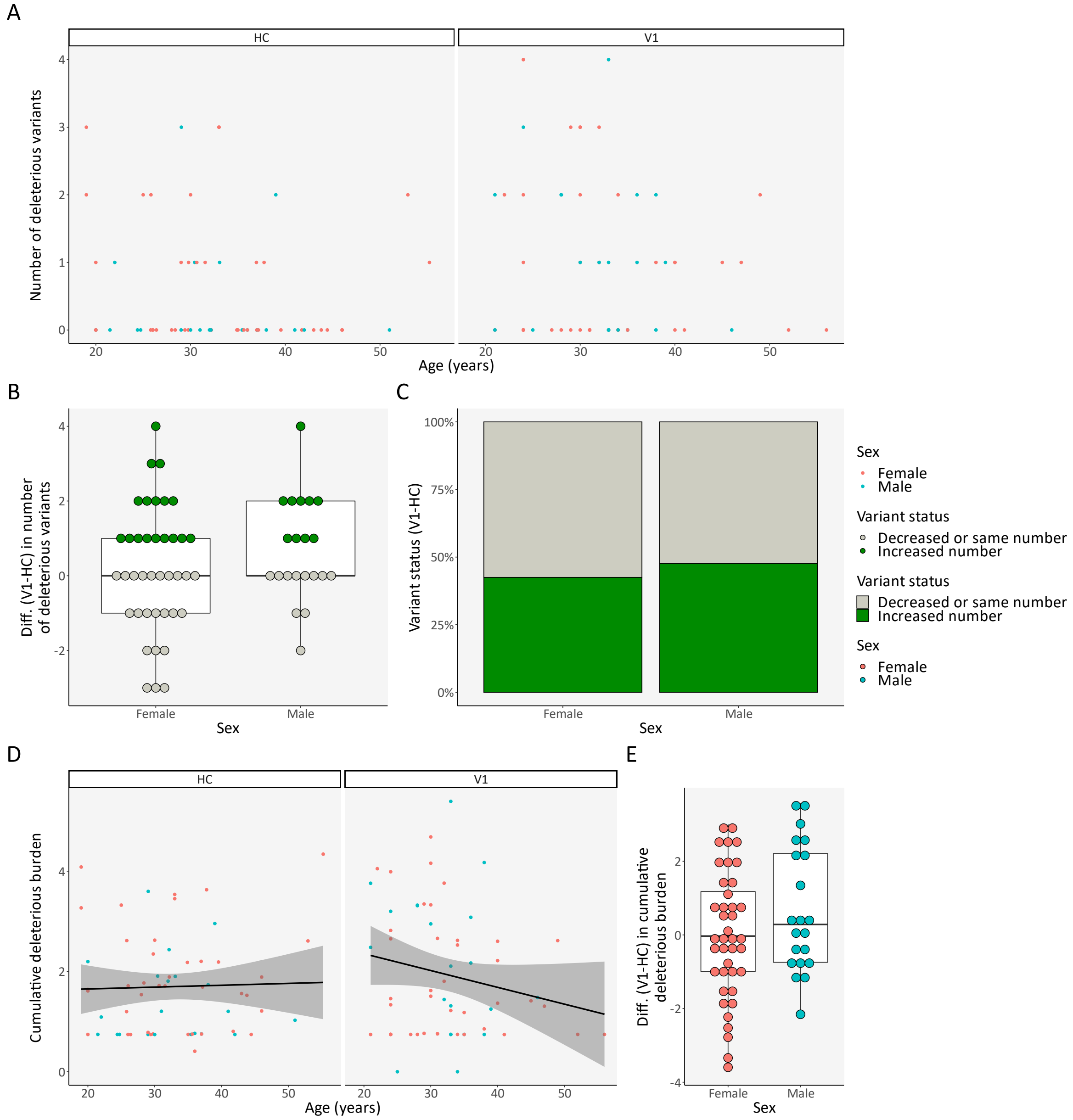

#### Figure S13 — Deleterious variants: Cross-sectional comparison (age and sex)

(**A**) Correlation between number of deleterious variants and age, per subject type and sex; (**B**) Difference in number of deleterious variants per triplet (PwMS at VIS1-HC), per sex; (**C**) Proportion of triplets with an increased or equal/decreased number of deleterious variants, per sex; (**D**) Correlation between cumulative deleterious burden and age, per subject type and sex; (**E**) Difference in cumulative deleterious burden per triplet (PwMS at VIS1-HC), per sex. Abbreviations: Diff. — difference; HC — healthy control; PwMS — patient with Clinically Isolated Syndrome/Relapsing-Remitting Multiple Sclerosis; V1 — patient with Clinically Isolated Syndrome/Relapsing-Remitting Multiple Sclerosis at visit 1; VIS1 — visit 1.

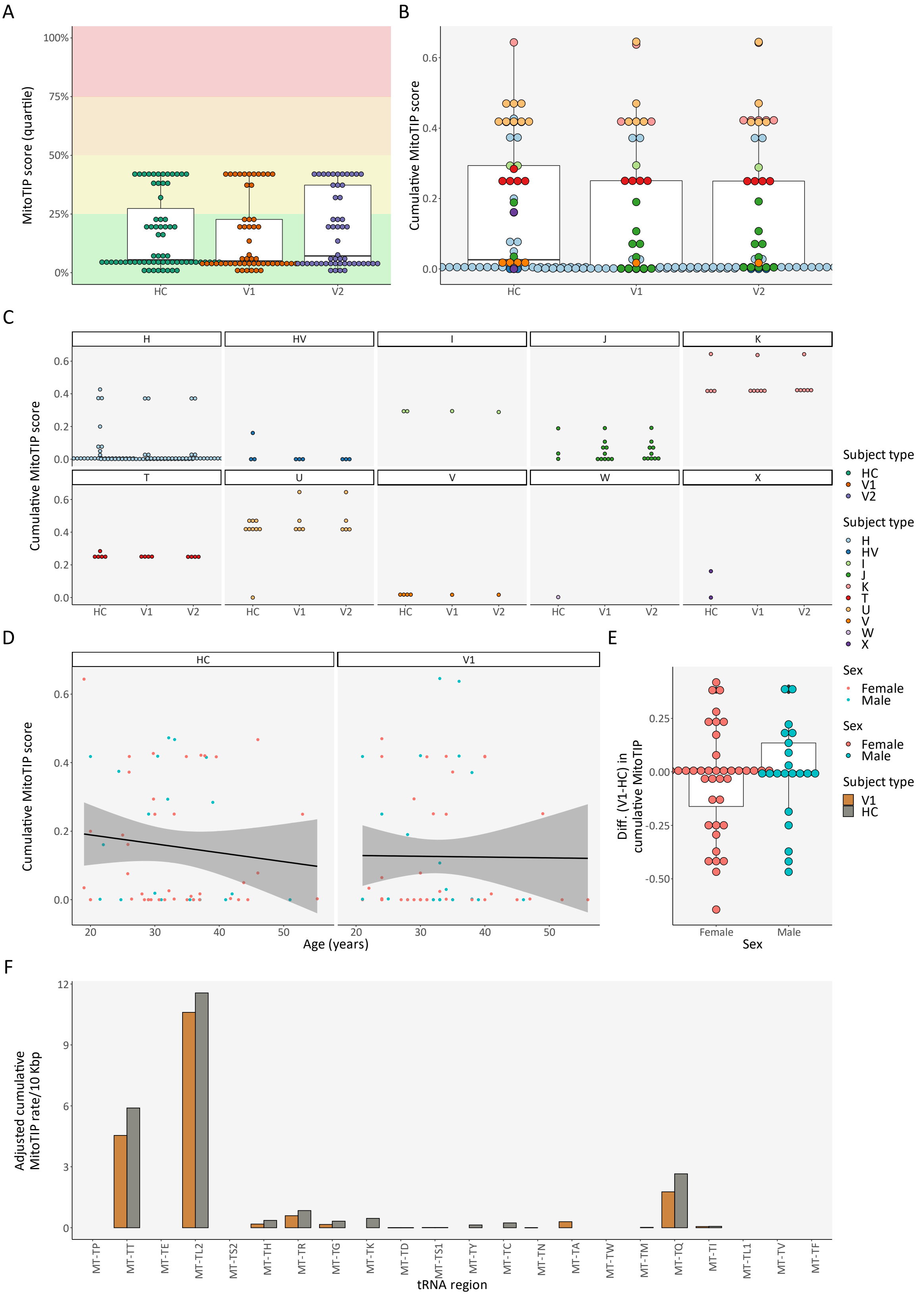

#### Figure S14 — tRNA variants: Cross-sectional comparison

(**A**) MitoTIP score (in quartiles) of all variants, per subject type — the green background indicates the range for "likely benign" variants, yellow for "possibly benign" variants, orange for "possibly pathogenic" variants, and red for "likely pathogenic" variants; (**B**) Cumulative MitoTIP score, per subject type and haplogroup; (**C**) Expansion of **Figure S14B** for better visualization; (**D**) Correlation between cumulative MitoTIP score and age, per subject type and sex; (**E**) Difference in cumulative MitoTIP score per triplet (PwMS at VIS1-HC), per sex; (**F**) Relative cumulative MitoTIP score for each tRNA region/locus, per subject type. Abbreviations: bp — base pair; Diff. — difference; HC — healthy control; PwMS — patient with Clinically Isolated Syndrome/Relapsing-Remitting Multiple Sclerosis; tRNA — transfer RNA; V1 — patient with Clinically Isolated Syndrome/Relapsing-Remitting Multiple Sclerosis at visit 1; VIS1 — visit 1.

#### Table S12 — Mean cumulative MitoTIP score per haplogroup (simplified). Dunn test, after a Kruskal-Wallis test, with FDR

| Haplogroups | Mean cumulative MitoTIP score | Adjusted *p*-value |
|---|---|---|
| K ~ T | 0.46 ~ 0.25 | 2.54E-01 |
| K ~ U | 0.46 ~ 0.42 | 8.95E-01 |
| K ~ V | 0.46 ~ 0.02 | 4.45E-02 |
| U ~ V | 0.42 ~ 0.02 | 4.45E-02 |
| I ~ J | 0.29 ~ 0.05 | 2.26E-01 |
| I ~ K | 0.29 ~ 0.46 | 6.06E-01 |
| I ~ T | 0.29 ~ 0.25 | 8.17E-01 |
| I ~ U | 0.29 ~ 0.42 | 6.28E-01 |
| I ~ V | 0.29 ~ 0.02 | 3.20E-01 |
| T ~ U | 0.25 ~ 0.42 | 2.54E-01 |
| T ~ V | 0.25 ~ 0.02 | 3.17E-01 |
| J ~ K | 0.05 ~ 0.46 | 2.49E-03 |
| J ~ T | 0.05 ~ 0.25 | 1.30E-01 |
| J ~ U | 0.05 ~ 0.42 | 1.52E-03 |
| J ~ V | 0.05 ~ 0.02 | 8.17E-01 |
| H ~ HV | 0.04 ~ 0.03 | 6.65E-01 |
| H ~ I | 0.04 ~ 0.29 | 3.72E-02 |
| H ~ J | 0.04 ~ 0.05 | 2.14E-01 |
| H ~ K | 0.04 ~ 0.46 | 2.20E-07 |
| H ~ T | 0.04 ~ 0.25 | 1.44E-03 |
| H ~ U | 0.04 ~ 0.42 | 7.37E-09 |
| H ~ V | 0.04 ~ 0.02 | 2.54E-01 |
| HV ~ I | 0.03 ~ 0.29 | 4.27E-02 |
| HV ~ J | 0.03 ~ 0.05 | 2.42E-01 |
| HV ~ K | 0.03 ~ 0.46 | 1.99E-04 |
| HV ~ T | 0.03 ~ 0.25 | 1.25E-02 |
| HV ~ U | 0.03 ~ 0.42 | 1.47E-04 |
| HV ~ V | 0.03 ~ 0.02 | 2.42E-01 |

Significant *p*-values are highlighted. Abbreviations: FDR — false discovery rate.

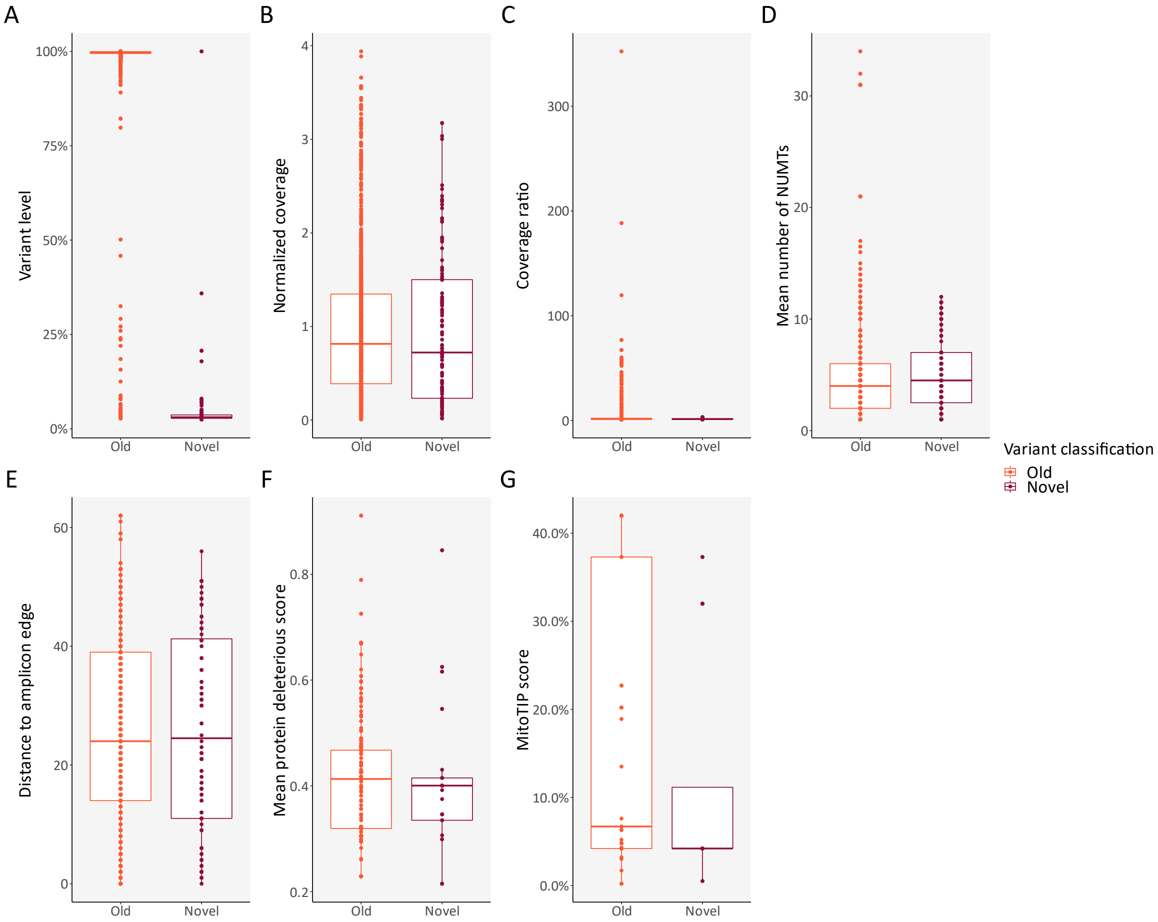

**Figure S15 — Longitudinal changes in PwMS: Novel *vs.* old variants**

(**A–G**) Comparison between novel and old variants for variant level, normalized coverage, coverage ratio, mean number of NUMTs, distance to amplicon edge, mean protein deleterious score, and MitoTIP score, respectively. Abbreviations: NUMTs — nuclear insertions of mitochondrial DNA; PwMS — patients with Clinically Isolated Syndrome/Relapsing-Remitting Multiple Sclerosis.

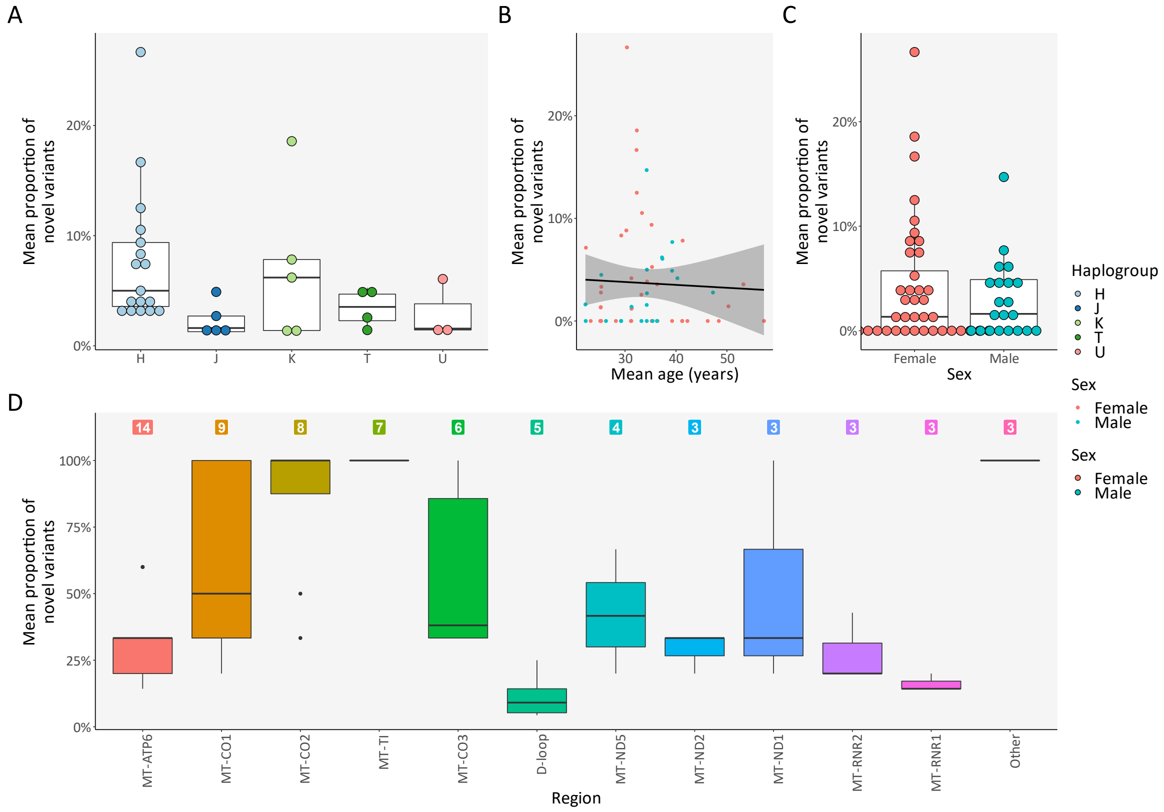

#### Figure S16 — Longitudinal changes in PwMS: Proportion of novel variants

(**A**) Mean proportion of novel variants per PwMS (both VIS1 and VIS2), per haplogroup with at least three samples and excluding samples without novel variants; (**B**) Correlation between mean proportion of novel variants per PwMS (both VIS1 and VIS2) and age; (**C**) Mean proportion of novel variants per PwMS (both VIS1 and VIS2), per sex; (**D**) Mean proportion of novel variants per PwMS (both VIS1 and VIS2), per region with at least three samples — colored labels indicate the number of samples for each region. Abbreviations: PwMS — patient(s) with Clinically Isolated Syndrome/Relapsing-Remitting Multiple Sclerosis; VIS1 — visit 1; VIS2 — visit 2.

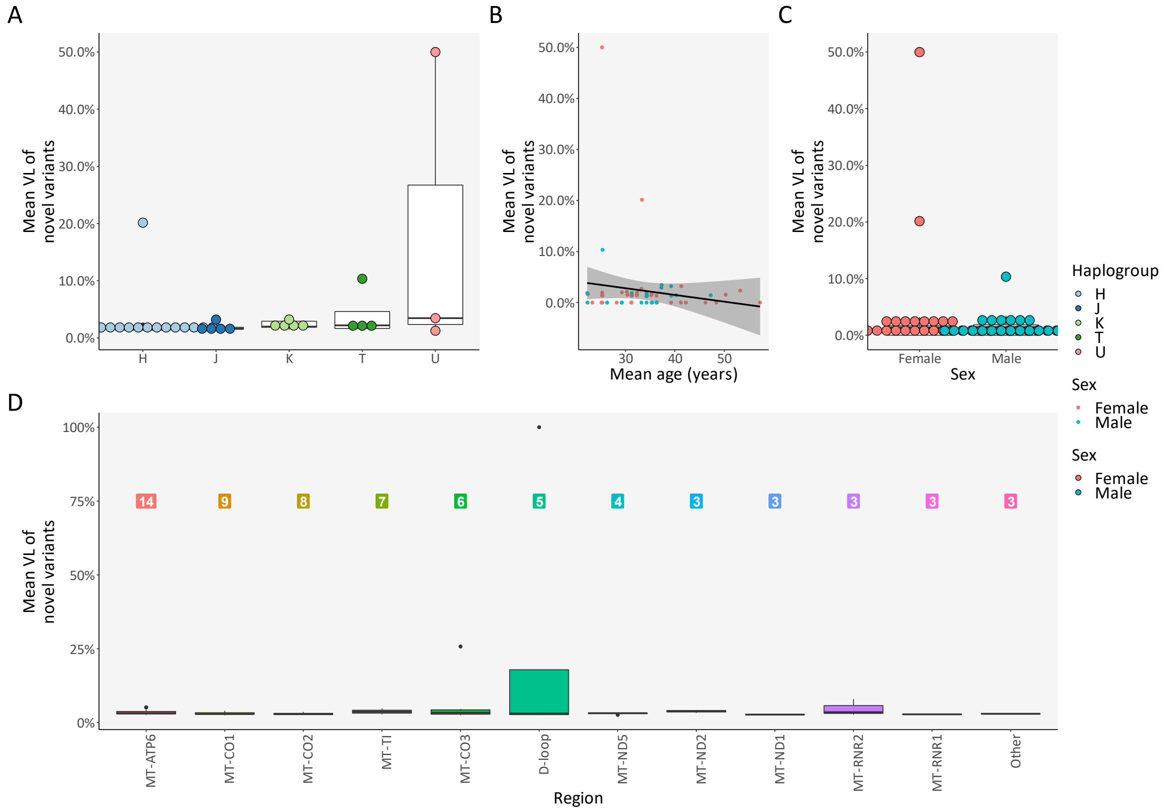

#### Figure S17 — Longitudinal changes in PwMS: Variant level of novel variants

(**A**) Mean variant level of novel variants per PwMS (both VIS1 and VIS2), per haplogroup with at least three samples and excluding samples without novel variants; (**B**) Correlation between mean variant level of novel variants per PwMS (both VIS1 and VIS2) and age; (**C**) Mean variant level of novel variants per PwMS (both VIS1 and VIS2), per sex; (**D**) Mean variant level of novel variants per PwMS (both VIS1 and VIS2), per region with at least three samples — colored labels indicate the number of samples for each region. Abbreviations: PwMS — patient(s) with Clinically Isolated Syndrome/Relapsing-Remitting Multiple Sclerosis; VIS1 — visit 1; VIS2 — visit 2; VL — variant level.

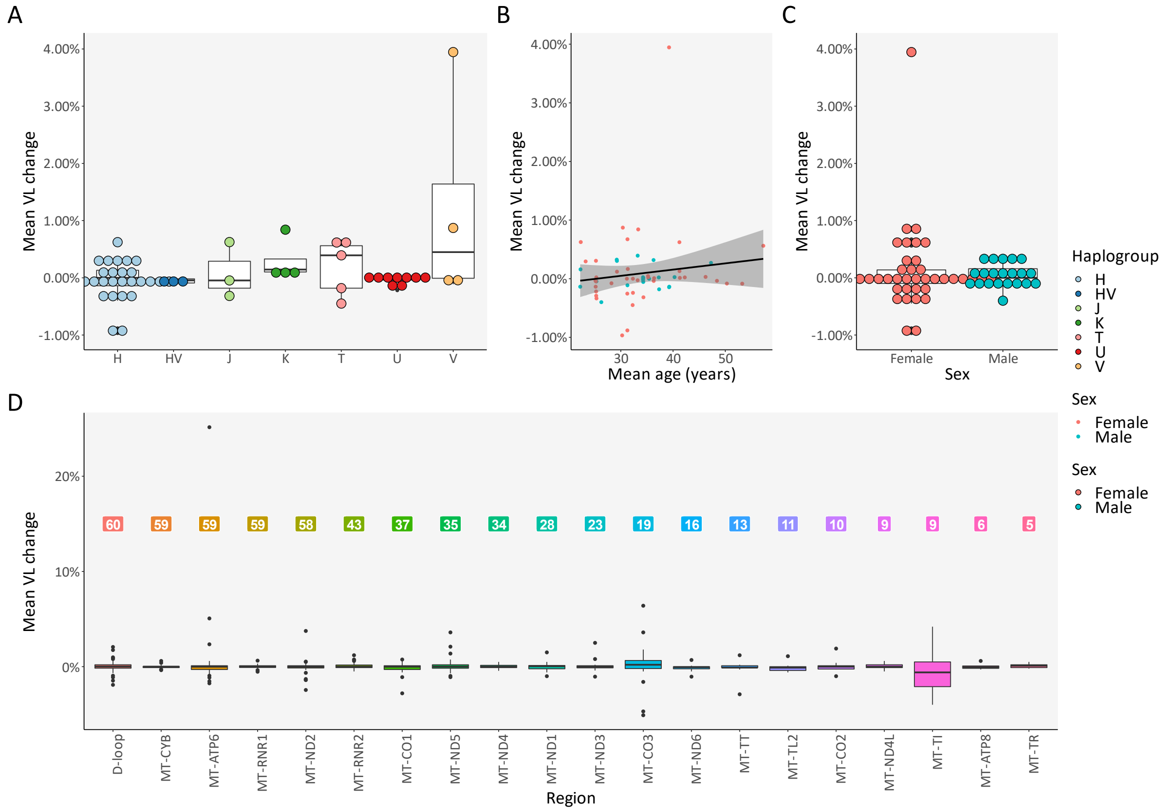

**Figure S18 — Longitudinal changes in PwMS: Variant level change of old variants**

(**A**) Mean variant level change of old variants per PwMS (both VIS1 and VIS2), per haplogroup with at least three samples; (**B**) Correlation between mean variant level change of old variants per PwMS (both VIS1 and VIS2) and age; (**C**) Mean variant level change of old variants per PwMS (both VIS1 and VIS2), per sex; (**D**) Mean variant level change of old variants per PwMS (both VIS1 and VIS2), per region with at least three samples — colored labels indicate the number of samples for each region. Abbreviations: PwMS — patient(s) with Clinically Isolated Syndrome/Relapsing-Remitting Multiple Sclerosis; VIS1 — visit 1; VIS2 — visit 2; VL — variant level.

#### Table S13 — Mean proportion of novel variants per region. Dunn test, after a Kruskal-Wallis test, with FDR

| Regions | Mean proportion of novel variants | Adjusted *p*-value |
|---|---|---|
| MT-TI ~ MT-ND1 | 1.00 ~ 0.51 | 1.98E-01 |
| MT-TI ~ MT-RNR2 | 1.00 ~ 0.28 | 4.36E-02 |
| MT-TI ~ MT-RNR1 | 1.00 ~ 0.16 | 3.93E-03 |
| MT-TI ~ Other | 1.00 ~ 1.00 | 1.00E+00 |
| MT-CO2 ~ MT-CO1 | 0.85 ~ 0.63 | 3.93E-01 |
| MT-CO2 ~ MT-ND2 | 0.85 ~ 0.29 | 1.16E-01 |
| MT-CO2 ~ MT-TI | 0.85 ~ 1.00 | 6.71E-01 |
| MT-CO2 ~ MT-ND1 | 0.85 ~ 0.51 | 2.98E-01 |
| MT-CO2 ~ MT-RNR2 | 0.85 ~ 0.28 | 9.25E-02 |
| MT-CO2 ~ MT-RNR1 | 0.85 ~ 0.16 | 9.64E-03 |
| MT-CO2 ~ Other | 0.85 ~ 1.00 | 7.52E-01 |
| MT-CO1 ~ MT-ND2 | 0.63 ~ 0.29 | 3.26E-01 |
| MT-CO1 ~ MT-TI | 0.63 ~ 1.00 | 2.26E-01 |
| MT-CO1 ~ MT-ND1 | 0.63 ~ 0.51 | 6.57E-01 |
| MT-CO1 ~ MT-RNR2 | 0.63 ~ 0.28 | 2.58E-01 |
| MT-CO1 ~ MT-RNR1 | 0.63 ~ 0.16 | 5.56E-02 |
| MT-CO1 ~ Other | 0.63 ~ 1.00 | 3.42E-01 |
| MT-CO3 ~ MT-ATP6 | 0.57 ~ 0.31 | 2.16E-01 |
| MT-CO3 ~ MT-CO2 | 0.57 ~ 0.85 | 3.93E-01 |
| MT-CO3 ~ MT-CO1 | 0.57 ~ 0.63 | 9.54E-01 |
| MT-CO3 ~ MT-ND2 | 0.57 ~ 0.29 | 3.71E-01 |
| MT-CO3 ~ MT-TI | 0.57 ~ 1.00 | 2.33E-01 |
| MT-CO3 ~ MT-ND1 | 0.57 ~ 0.51 | 7.04E-01 |
| MT-CO3 ~ MT-RNR2 | 0.57 ~ 0.28 | 3.26E-01 |
| MT-CO3 ~ MT-RNR1 | 0.57 ~ 0.16 | 9.07E-02 |
| MT-CO3 ~ Other | 0.57 ~ 1.00 | 3.42E-01 |
| MT-ND1 ~ MT-RNR2 | 0.51 ~ 0.28 | 6.08E-01 |
| MT-ND1 ~ MT-RNR1 | 0.51 ~ 0.16 | 2.38E-01 |
| MT-ND1 ~ Other | 0.51 ~ 1.00 | 2.57E-01 |
| MT-ND5 ~ MT-CO3 | 0.42 ~ 0.57 | 6.57E-01 |
| MT-ND5 ~ MT-ATP6 | 0.42 ~ 0.31 | 5.85E-01 |
| MT-ND5 ~ MT-CO2 | 0.42 ~ 0.85 | 2.26E-01 |
| MT-ND5 ~ MT-CO1 | 0.42 ~ 0.63 | 5.85E-01 |
| MT-ND5 ~ MT-ND2 | 0.42 ~ 0.29 | 6.71E-01 |
| MT-ND5 ~ MT-TI | 0.42 ~ 1.00 | 1.22E-01 |
| MT-ND5 ~ MT-ND1 | 0.42 ~ 0.51 | 9.73E-01 |
| MT-ND5 ~ MT-RNR2 | 0.42 ~ 0.28 | 6.13E-01 |
| MT-ND5 ~ MT-RNR1 | 0.42 ~ 0.16 | 2.33E-01 |
| MT-ND5 ~ Other | 0.42 ~ 1.00 | 2.19E-01 |
| MT-ATP6 ~ MT-CO2 | 0.31 ~ 0.85 | 9.64E-03 |
| MT-ATP6 ~ MT-CO1 | 0.31 ~ 0.63 | 1.16E-01 |
| MT-ATP6 ~ MT-ND2 | 0.31 ~ 0.29 | 1.00E+00 |
| MT-ATP6 ~ MT-TI | 0.31 ~ 1.00 | 3.93E-03 |

**Table S13 — Mean proportion of novel variants per region. Dunn test, after a Kruskal-Wallis test, with FDR (continued)**

| Regions | Mean proportion of novel variants | Adjusted *p*-value |
|---|---|---|
| MT-ATP6 ~ MT-ND1 | 0.31 ~ 0.51 | 5.85E-01 |
| MT-ATP6 ~ MT-RNR2 | 0.31 ~ 0.28 | 9.47E-01 |
| MT-ATP6 ~ MT-RNR1 | 0.31 ~ 0.16 | 3.42E-01 |
| MT-ATP6 ~ Other | 0.31 ~ 1.00 | 3.92E-02 |
| MT-ND2 ~ MT-TI | 0.29 ~ 1.00 | 5.79E-02 |
| MT-ND2 ~ MT-ND1 | 0.29 ~ 0.51 | 6.62E-01 |
| MT-ND2 ~ MT-RNR2 | 0.29 ~ 0.28 | 9.54E-01 |
| MT-ND2 ~ MT-RNR1 | 0.29 ~ 0.16 | 4.70E-01 |
| MT-ND2 ~ Other | 0.29 ~ 1.00 | 1.16E-01 |
| MT-RNR2 ~ MT-RNR1 | 0.28 ~ 0.16 | 5.54E-01 |
| MT-RNR2 ~ Other | 0.28 ~ 1.00 | 9.64E-02 |
| MT-RNR1 ~ Other | 0.16 ~ 1.00 | 1.44E-02 |
| D-loop ~ MT-ND5 | 0.12 ~ 0.42 | 1.35E-01 |
| D-loop ~ MT-CO3 | 0.12 ~ 0.57 | 2.52E-02 |
| D-loop ~ MT-ATP6 | 0.12 ~ 0.31 | 2.12E-01 |
| D-loop ~ MT-CO2 | 0.12 ~ 0.85 | 1.19E-03 |
| D-loop ~ MT-CO1 | 0.12 ~ 0.63 | 9.64E-03 |
| D-loop ~ MT-ND2 | 0.12 ~ 0.29 | 3.42E-01 |
| D-loop ~ MT-TI | 0.12 ~ 1.00 | 4.25E-04 |
| D-loop ~ MT-ND1 | 0.12 ~ 0.51 | 1.59E-01 |
| D-loop ~ MT-RNR2 | 0.12 ~ 0.28 | 3.93E-01 |
| D-loop ~ MT-RNR1 | 0.12 ~ 0.16 | 9.47E-01 |
| D-loop ~ Other | 0.12 ~ 1.00 | 3.93E-03 |

Significant *p*-values are highlighted. Abbreviations: FDR — false discovery rate.

#### Table S14 — PwMS' medications: Summary

| Medication type | Drug | *N* at VIS1 | *N* at VIS2 |
|---|---|---|---|
| Multiple Sclerosis | None | 45 | 32 |
| Multiple Sclerosis | Glatiramer acetate | 9 | 11 |
| Multiple Sclerosis | Interferon beta | 5 | 8 |
| Multiple Sclerosis | Fumarate | 2 | 8 |
| Multiple Sclerosis | Fingolimod | - | 2 |
| CNS and Neuromuscular | Biperiden | 1 | - |
| CNS and Neuromuscular | Lorazepam | 1 | - |
| CNS and Neuromuscular | Ziprasidone | 1 | 1 |
| CNS and Neuromuscular | Zopiclone | 1 | - |
| CNS and Neuromuscular | Baclofen | - | 1 |
| CNS and Neuromuscular | Citalopram | - | 1 |
| CNS and Neuromuscular | Mirtazapine | - | 1 |
| CNS and Neuromuscular | Sertraline | - | 1 |
| Cardiovascular | Ramipril | 2 | 1 |
| Cardiovascular | Irbesartan + Hydrochlorothiazide | 1 | 1 |
| Cardiovascular | Losartan | 1 | 1 |
| Cardiovascular | Metoprolol | 1 | 2 |
| Cardiovascular | Acetylsalicylic acid | - | 2 |
| Cardiovascular | Valsartan | - | 2 |
| Cardiovascular | Nebivolol | - | 1 |
| Endocrine | Levothyroxine | 10 | 11 |
| Endocrine | Oral contraceptive | 7 | 9 |
| Endocrine | Triiodothyronine | - | 1 |
| Supplemental | Vitamin D | 16 | 24 |
| Supplemental | Vitamin B12 | 3 | 3 |
| Supplemental | Selenium | 2 | 1 |
| Supplemental | Vitamin B9 | 2 | - |
| Supplemental | Iron | 1 | 1 |
| Supplemental | Isoflavone | 1 | 1 |
| Supplemental | Lipoic acid | 1 | - |
| Supplemental | Omega-3 fatty acids | 1 | - |
| Supplemental | Vitamin B complex | 1 | 1 |
| Supplemental | Vitamin B6 + Vitamin B12 | 1 | - |
| Supplemental | Vitamin D + Calcium | 1 | - |
| Supplemental | Vitamin E | 1 | - |
| Supplemental | Vitamin K | 1 | - |
| Supplemental | Zinc | 1 | - |
| Supplemental | Magnesium | - | 2 |
| Other | Ibuprofen | 3 | 3 |
| Other | Desloratadine | 1 | - |
| Other | Hydroxychloroquine | 1 | 1 |
| Other | Intrauterine contraceptive device | 1 | 1 |
| Other | Pantoprazole | 1 | 1 |
| Other | Rupatadine | 1 | - |

**Table S14 — PwMS' medications: Summary (continued)**

| Medication type | Drug | *N* at VIS1 | *N* at VIS2 |
|---|---|---|---|
| Other | Topical corticosteroid | 1 | 2 |
| Other | Doxycycline | - | 1 |
| Other | Formoterol | - | 1 |
| Other | Ivermectin (topical) | - | 1 |
| Other | Paracetamol | - | 1 |

Abbreviations: CNS — central nervous system; *N* — number; PwMS — patients with Clinically Isolated Syndrome/Relapsing-Remitting Multiple Sclerosis; VIS1 — visit 1; VIS2 — visit 2.

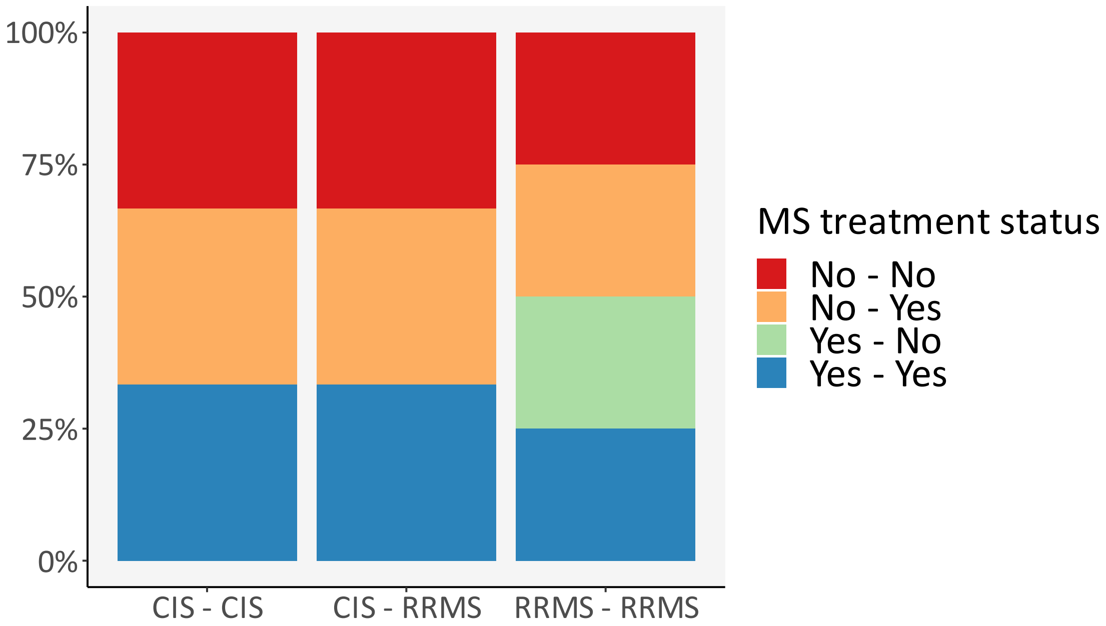

**Figure S19 — MS treatment status between VIS1 and VIS2, per diagnostic evolution**

Abbreviations: CIS — Clinically Isolated Syndrome; MS — Multiple Sclerosis; RRMS — Relapsing-Remitting Multiple Sclerosis; VIS1 — visit 1; VIS2 — visit 2.

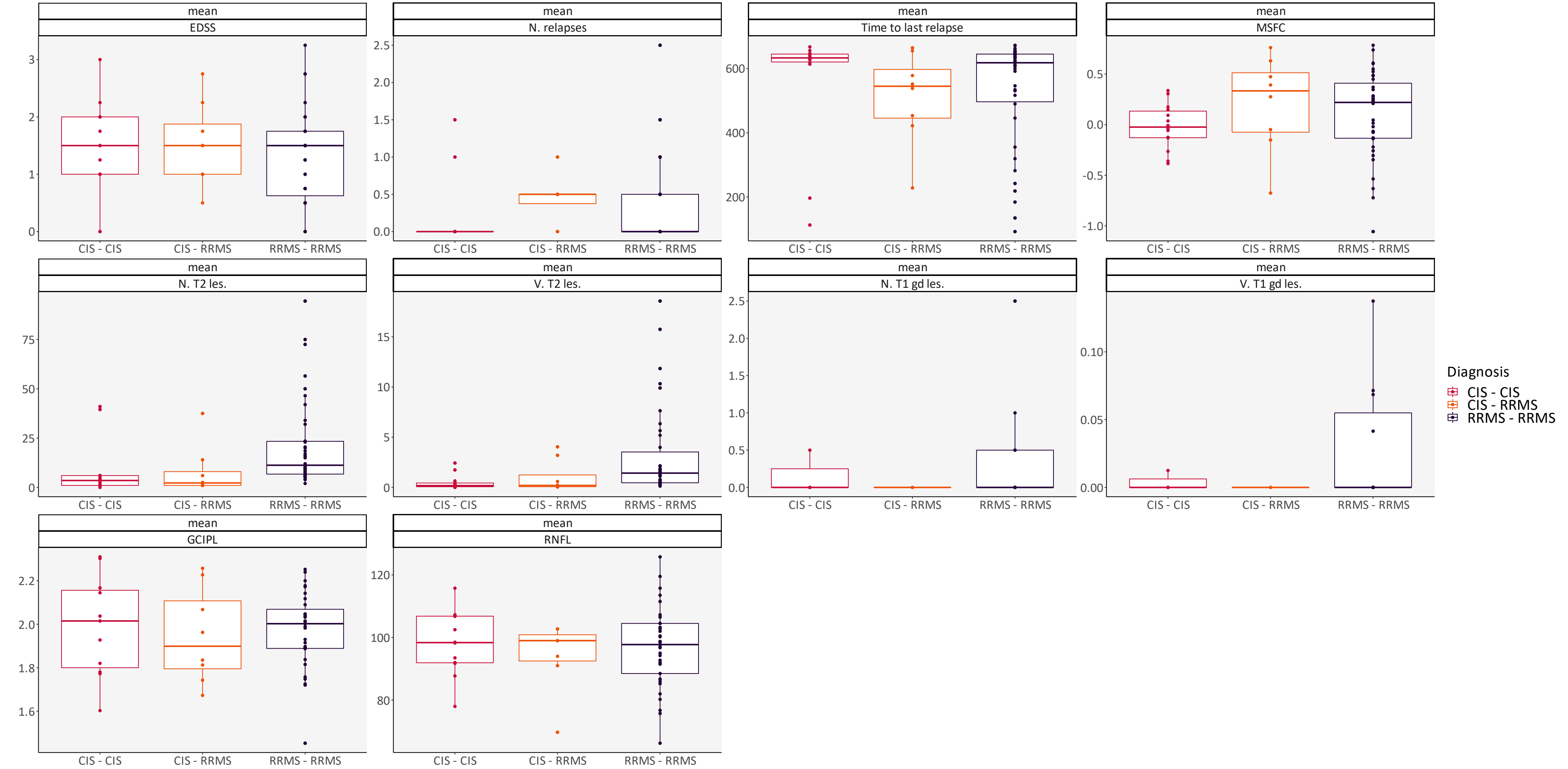

**Figure S20 — Clinical variables and diagnostic evolution: Mean values**

Distribution according to diagnostic evolution, from VIS1 to VIS2, regarding the mean value between VIS1 and VIS2 for EDSS, number of relapses, time to last relapse in days, MSFC, number of T2 hyperintense lesions, volume of T2 hyperintense lesions in mL, number of gadolinium-enhancing T1 lesions, volume of gadolinium-enhancing T1 lesions in mL, GCIPL volume in mm$^3$, RNFL thickness in µm. Abbreviations: CIS — Clinically Isolated Syndrome; EDSS — expanded disability status scale; GCIPL — ganglion cell-inner plexiform layer; gd — gadolinium; les. — lesions; MSFC — Multiple Sclerosis functional composite; N. — number; RNFL — retinal nerve fiber layer; RRMS — Relapsing-Remitting Multiple Sclerosis; V. — volume; VIS1 — visit 1; VIS2 — visit 2.

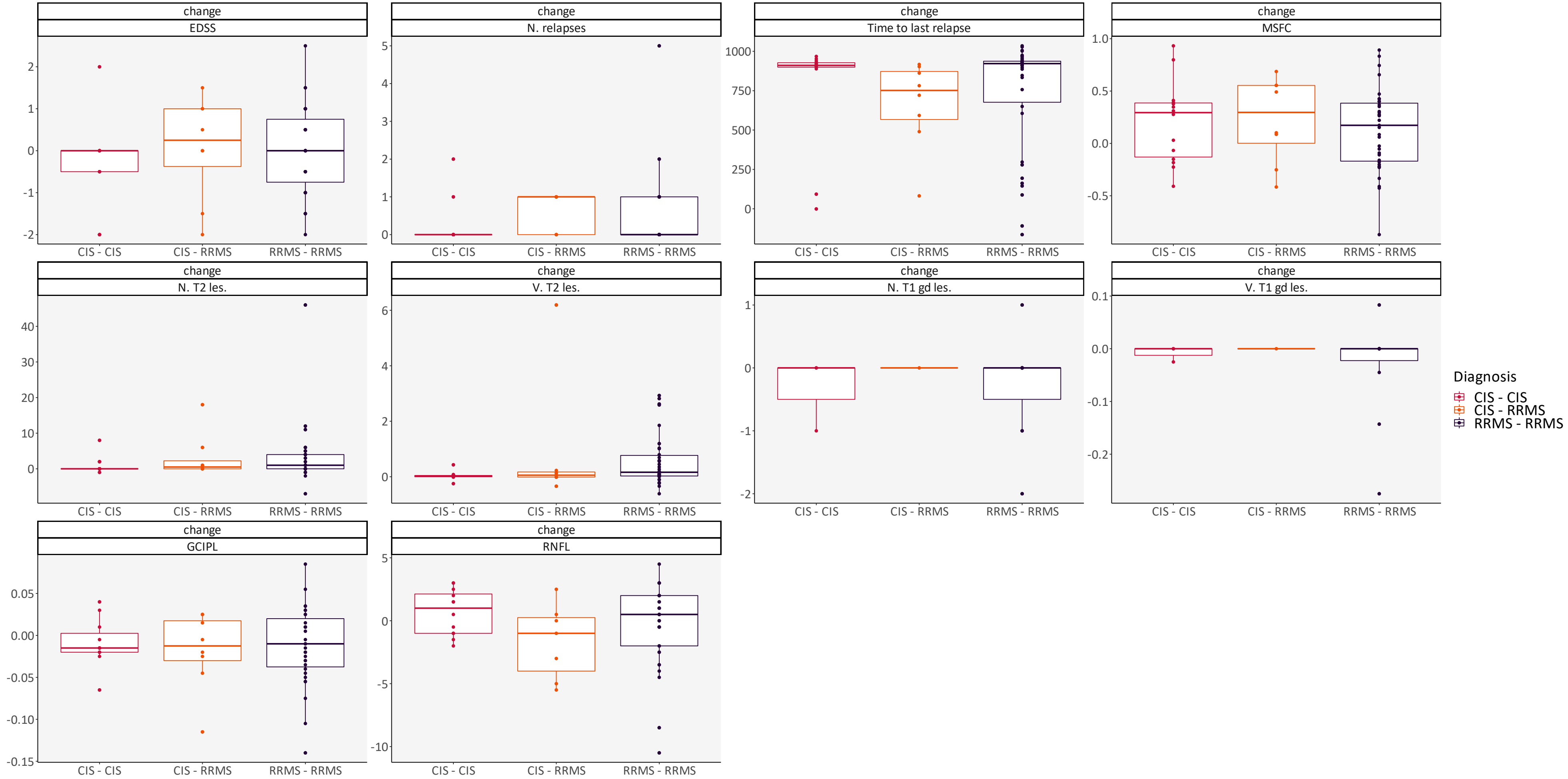

**Figure S21 — Clinical variables and diagnostic evolution: Difference**

Distribution according to diagnostic evolution, from VIS1 to VIS2, regarding the difference between VIS2 and VIS1 for EDSS, number of relapses, time to last relapse in days, MSFC, number of T2 hyperintense lesions, volume of T2 hyperintense lesions in mL, number of gadolinium-enhancing T1 lesions, volume of gadolinium-enhancing T1 lesions in mL, GCIPL volume in mm$^3$, RNFL thickness in µm. Abbreviations: CIS — Clinically Isolated Syndrome; EDSS — expanded disability status scale; GCIPL — ganglion cell-inner plexiform layer; gd — gadolinium; les. — lesions; MSFC — Multiple Sclerosis functional composite; N. — number; RNFL — retinal nerve fiber layer; RRMS — Relapsing-Remitting Multiple Sclerosis; V. — volume; VIS1 — visit 1; VIS2 — visit 2.

#### Table S15 — Clinical variables and diagnostic evolution: Mean values *vs.* Differences. Batch Kruskal-Wallis tests, with FDR

| Comparison | Mean adjusted *p*-value | Difference adjusted *p*-value |
|---|---|---|
| EDSS ~ Diagnosis | 8.68E-01 | 8.38E-01 |
| N. relapses ~ Diagnosis | 3.07E-01 | 3.70E-01 |
| Time to last relapse ~ Diagnosis | 8.67E-01 | 3.70E-01 |
| MSFC ~ Diagnosis | 6.05E-01 | 9.24E-01 |
| N. T2 les. ~ Diagnosis | 1.29E-03 | 3.70E-01 |
| V. T2 les. ~ Diagnosis | 1.29E-03 | 3.68E-01 |
| N. T1 gd les. ~ Diagnosis | 8.68E-01 | 9.24E-01 |
| V. T1 gd les. ~ Diagnosis | 8.68E-01 | 9.24E-01 |
| GCIPL ~ Diagnosis | 8.68E-01 | 9.24E-01 |
| RNFL ~ Diagnosis | 8.68E-01 | 5.13E-01 |

Significant p-values are highlighted. Abbreviations: EDSS — expanded disability status scale; FDR — false discovery rate; GCIPL — ganglion cell-inner plexiform layer; gd — gadolinium; les. — lesions; MSFC — Multiple Sclerosis functional composite; N. — number; RNFL — retinal nerve fiber layer; V. — volume.

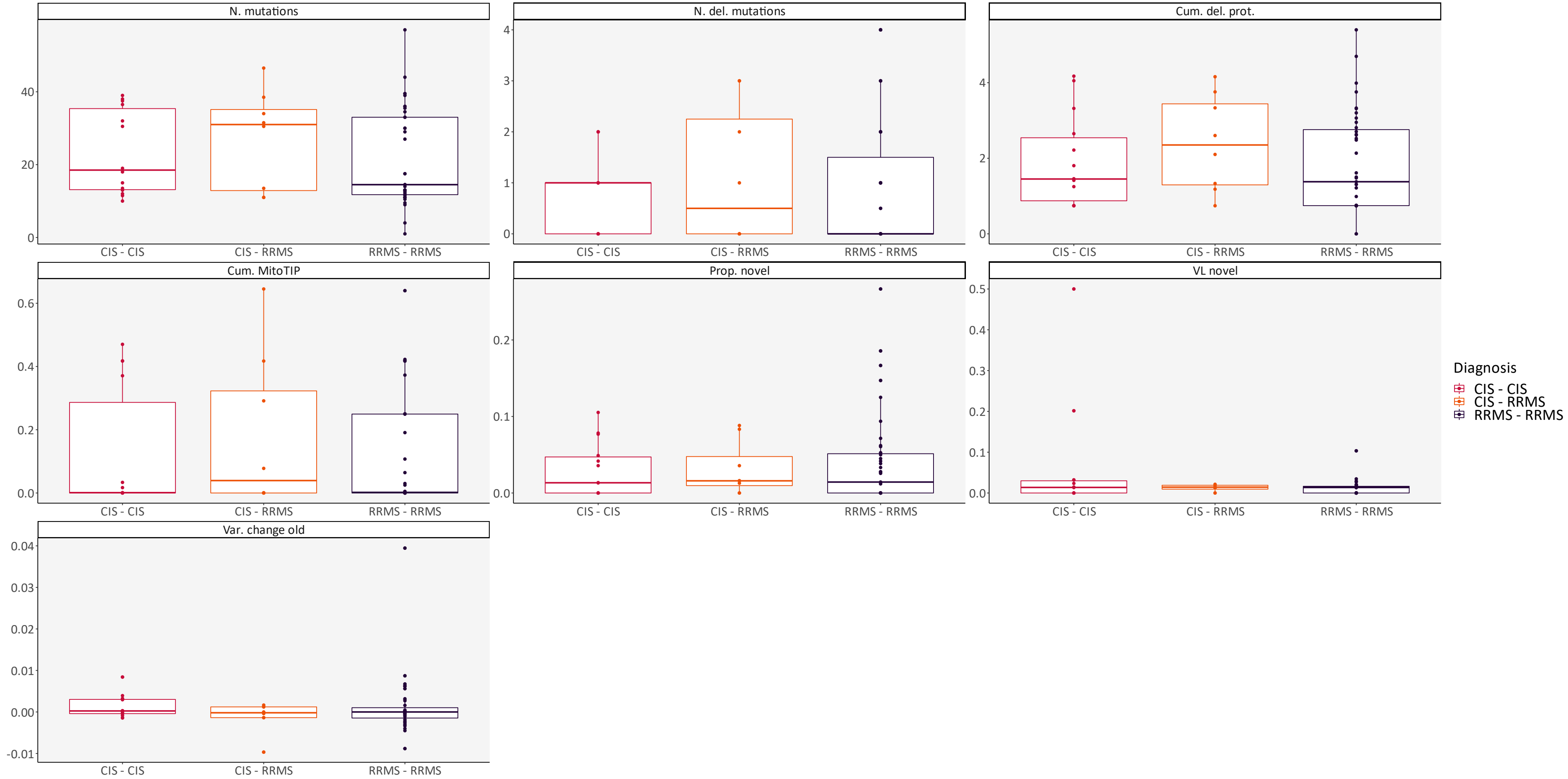

#### Figure S22 — WGS variables and diagnostic evolution

Distribution according to diagnostic evolution, from VIS1 to VIS2, regarding number of variants, number of deleterious variants, cumulative protein deleterious burden, cumulative MitoTIP, proportion of novel variants, mean VL of novel variants, and mean variant level change. Abbreviations: CIS — Clinically Isolated Syndrome; Cum. del. prot. — cumulative protein deleterious burden; Cum. MitoTIP — cumulative MitoTIP; del. — deleterious; N. — number; WGS — whole genome sequencing; Prop. — proportion; RRMS — Relapsing-Remitting Multiple Sclerosis; Var. — variant; VIS1 — visit 1; VIS2 — visit 2; VL — variant level.

**Table S16 — WGS variables and diagnostic evolution. Batch Kruskal-Wallis tests, with FDR**

| Comparison | Adjusted *p*-value |
|---|---|
| Cum. del. prot. ~ Diagnosis | 9.24E-01 |
| Cum. MitoTIP ~ Diagnosis | 9.24E-01 |
| N. del. mutations ~ Diagnosis | 9.24E-01 |
| N. mutations ~ Diagnosis | 9.24E-01 |
| Prop. novel ~ Diagnosis | 9.24E-01 |
| Var. change old ~ Diagnosis | 9.24E-01 |
| VL novel ~ Diagnosis | 9.24E-01 |

Abbreviations: Cum. del. prot. — cumulative protein deleterious burden; Cum. MitoTIP — cumulative MitoTIP; del. — deleterious; FDR — false discovery rate; N. — number; WGS — whole genome sequencing; Prop. — proportion; Var. — variant; VL — variant level.

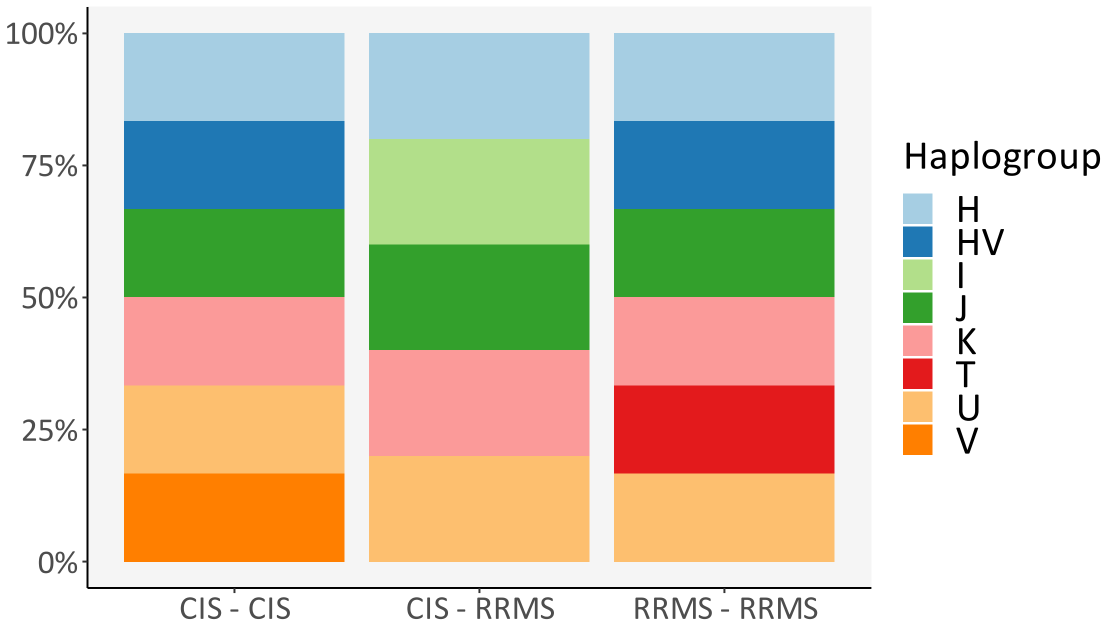

**Figure S23 — Haplogroup distribution per diagnostic evolution**

Abbreviations: CIS — Clinically Isolated Syndrome; RRMS — Relapsing-Remitting Multiple Sclerosis.

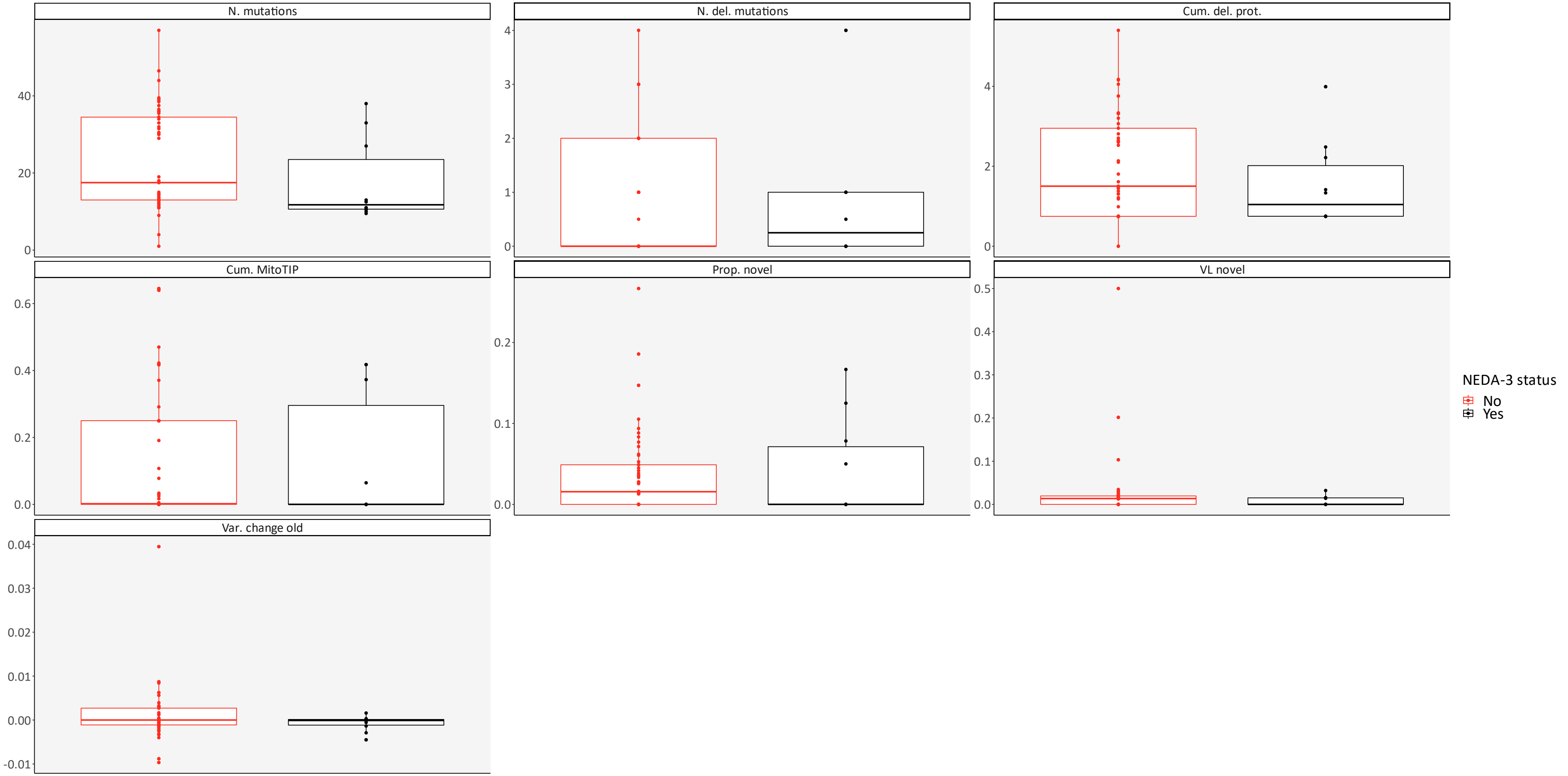

**Figure S24 — WGS variables and NEDA-3 status**

Distribution according to NEDA-3 status (Yes: *N*=9; Total: *N*=59), from VIS1 to VIS2, regarding number of variants, number of deleterious variants, cumulative protein deleterious burden, cumulative MitoTIP, proportion of novel variants, mean VL of novel variants, and mean variant level change. Abbreviations: Cum. del. prot. — cumulative protein deleterious burden; Cum. MitoTIP — cumulative MitoTIP; del. — deleterious; N. — number; NEDA — no evidence of disease activity; WGS — whole genome sequencing; Prop. — proportion; Var. — variant; VIS1 — visit 1; VIS2 — visit 2; VL — variant level.

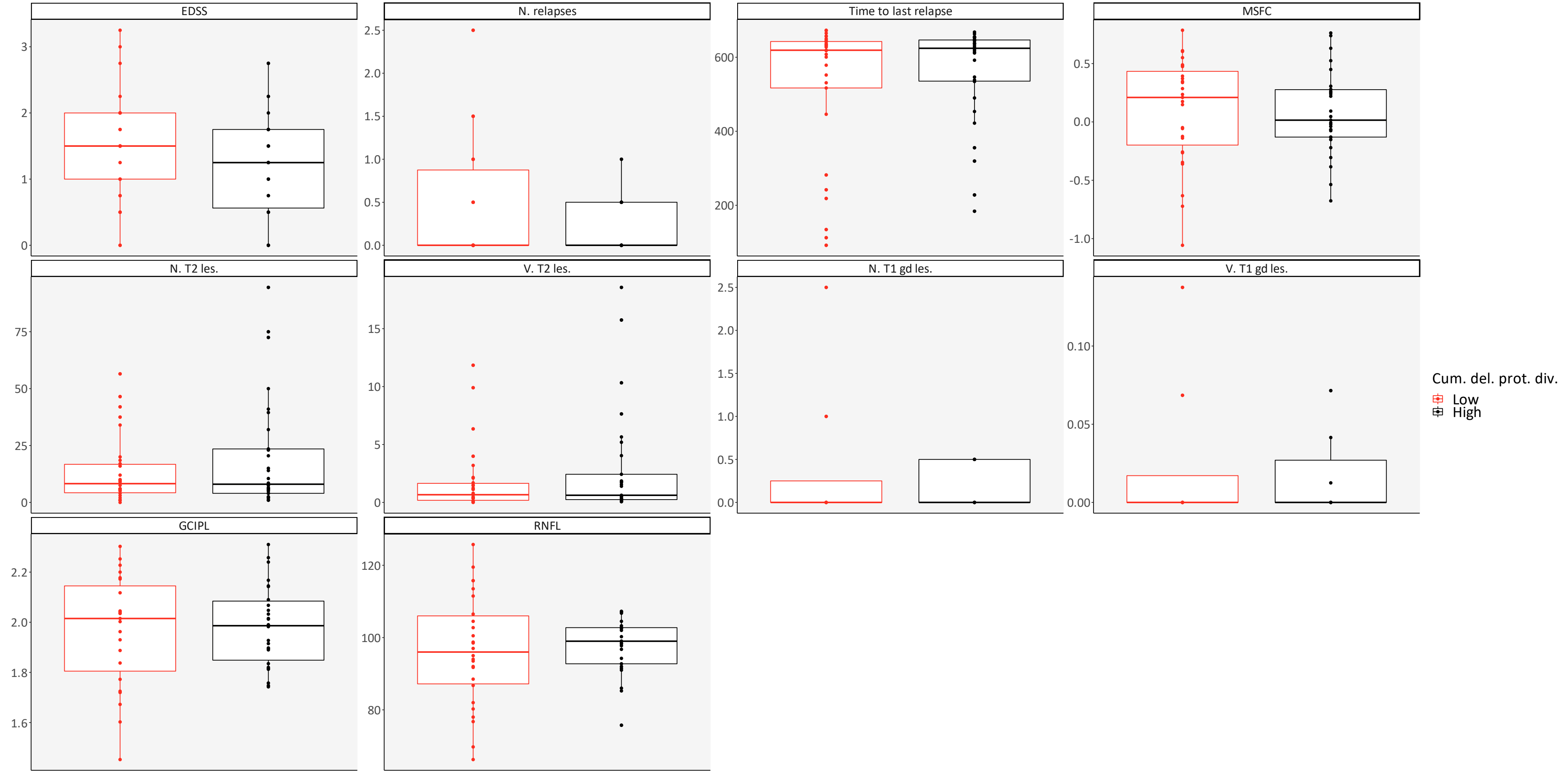

**Figure S25 — Clinical variables and cumulative protein deleterious burden**

Distribution according to cumulative protein deleterious burden (low — below median value and high — above median value; *N*=60), from VIS1 to VIS2, regarding the mean value between VIS1 and VIS2 for EDSS, number of relapses, time to last relapse in days, MSFC, number of T2 hyperintense lesions, volume of T2 hyperintense lesions in mL, number of gadolinium-enhancing T1 lesions, volume of gadolinium-enhancing T1 lesions in mL, GCIPL volume in mm$^3$, RNFL thickness in µm. Abbreviations: Cum. del. prot. div. — cumulative protein deleterious burden division; EDSS — expanded disability status scale; GCIPL — ganglion cell-inner plexiform layer; gd — gadolinium; les. — lesions; MSFC — Multiple Sclerosis functional composite; N. — number; RNFL — retinal nerve fiber layer; V. — volume; VIS1 — visit 1; VIS2 — visit 2.

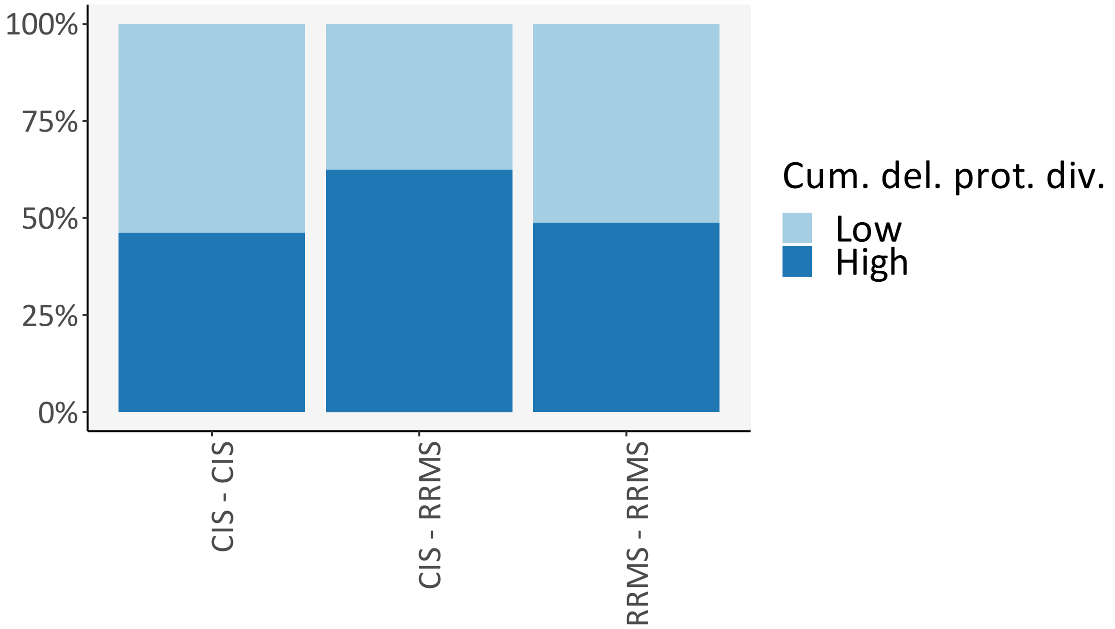

**Figure S26 — Cumulative protein deleterious burden per diagnostic evolution**

Abbreviations: CIS — Clinically Isolated Syndrome; Cum. del. prot. div. — cumulative protein deleterious burden division; RRMS — Relapsing-Remitting Multiple Sclerosis.
